# Two-phase sample selection strategies for design and analysis in post-genome wide association fine-mapping studies

**DOI:** 10.1101/2021.05.15.21257266

**Authors:** Osvaldo Espin-Garcia, Radu V. Craiu, Shelley B. Bull

## Abstract

Post-GWAS analysis, in many cases, focuses on fine-mapping targeted genetic regions discovered at GWAS-stage; that is, the aim is to pinpoint potential causal variants and susceptibility genes for complex traits and disease outcomes using next-generation sequencing (NGS) technologies. Large-scale GWAS cohorts are necessary to identify target regions given the typically modest genetic effect sizes. In this context, two-phase sampling design and analysis is a cost-reduction technique that utilizes data collected during phase 1 GWAS to select an informative subsample for phase 2 sequencing. The main goal is to make inference for genetic variants measured via NGS by efficiently combining data from phases 1 and 2. We propose two approaches for selecting a phase 2 design under a budget constraint. The first method identifies sampling fractions that select a phase 2 design yielding an asymptotic variance covariance matrix with certain optimal characteristics, e.g. smallest trace, via Lagrange multipliers (LM). The second relies on a genetic algorithm (GA) with a defined fitness function to identify exactly a phase 2 subsample. We perform comprehensive simulation studies to evaluate the empirical properties of the proposed designs for a genetic association study of a quantitative trait. We compare our methods against two ranked designs: residual-dependent sampling and a recently identified optimal design. Our findings demonstrate that the proposed designs, GA in particular, can render competitive power in combined phase 1 and 2 analysis compared to alternative designs while preserving type 1 error control. These results are especially apparent under the more practical scenario where design values need to be defined *a priori* and are subject to mispecification. We illustrate the proposed methods in a study of triglyceride levels in the North Finland Birth Cohort of 1966. R code to reproduce our results is available at github.com/egosv/TwoPhase_postGWAS.

## 1 Introduction

Genome-wide association studies (GWASs) have become well-established untargeted approaches for identifying genetic loci that influence the aetiology of complex diseases and traits. Single-nucleotide polymorphisms (SNPs) genotyped using GWAS arrays typically lack any known biological function. Consequently, in post-GWAS studies, identifying causal variants and susceptibility genes in GWAS-identified regions of association is the next important step for researchers. Identified variants and genes can become instrumental in personalized medicine from diagnosis and intervention to drug development and other forms of therapy.

Recent advances in next-generation sequencing (NGS) technologies allow investigators to sequence the entire human genome at the base-pair level, but, the costs of whole genome sequencing are relatively high in comparison to GWAS analysis. Targeted sequencing, which identifies all variants in a region with high-confidence, can be cost effective when fine mapping a genetic region identified at GWAS stage. Indeed, high-density sequence variants in the targeted region are typically in linkage disequilibrium (LD) with strongly associated SNPs from GWAS, making the latter good candidates as auxiliary covariates for subsample selection. Thus, two-phase sampling design and analysis^[1;2]^ emerges as a suitable cost-reduction technique in the post-GWAS context. The main goal of this strategy is to make inference on incompletely-observed sequencing data. At phase 1, GWAS data are collected for everyone in the study. At phase 2, sequencing data are collected only in a subsample of the phase 1 sample. The subsample is selected based on phase 1 information alone (outcome, auxiliary SNPs), making the sequence data missing-by-design in the non-selected individuals.

While the majority of the literature in two-phase sampling designs concentrates on effect estimation and hypothesis testing, relatively less attention has been paid to phase 2 sample selection. Specifically, most of the work examining optimal designs has focused on case-control studies^[3;4]^, in which for example, a *balanced* design (equal sample distribution across strata) has been recommended as near optimal^[5]^. Typically, in the design of case-control studies, optimization is performed to determine sampling fractions across predefined strata subject to a budget constraint on the phase 2 sample size^[6;7;8]^. Another approach, described in Zhao *et al*., seeks to optimize the sampling fraction (*ρ*) under simple random sampling considering asymptotic relative efficiency of the maximum likelihood estimators from the one-versus two-phase designs^[9]^. More recently, Tao *et al*. derived general optimal designs of two-phase studies paying special attention to continuous, binary, and time-to-event outcomes^[10]^. Specifically, Tao *et al*. demonstrate the relationship between their optimal design (hereafter referred to as TZL) and previously proposed (ranked) designs such as outcome-dependent and residual-dependent sampling (ODS and RDS, respectively).

In this report, we propose two approaches for two-phase sample selection in post-GWA fine-mapping studies. The resulting designs are valid and their implementation is available for all distributions in the exponential family. The first approach, LM, extends and adapts previous work primarily developed for case-control studies by solving a constrained optimization problem via Lagrange multipliers using numerical methods. The second approach, GA, exploits the advantages of genetic algorithms (GAs) for discrete optimization with fixed-subsets. To the best of our knowledge, this work introduces a novel usage of GAs in the context of selecting phase 2 designs.

In the next section we introduce a maximum likelihood framework for design and analysis of two-phase studies, and define the two approaches to select a phase 2 subsample. In addition, we contrast the proposed designs (LM and GA) with ranked designs (ODS, RDS, and TZL). In Section 3 we conduct simulation studies of a quantitative trait (QT) to evaluate the performance of the proposed designs against ranked designs under the ideal scenario in which all design quantities are known in advance. In Section 4, we assess a more practical scenario where the design values are misspecified using simulated data with realistic LD patterns from the 1000 Genomes Project. Our results show that the proposed designs, GA in particular, achieve competitive power against alternative designs under various scenarios. Additionally, in Section 5, we illustrate these methods in an application to the North Finland Birth Cohort of 1966. We conclude with a discussion of the advantages and challenges of the studied approaches as well as potential avenues of future research.

## 2 Phase 2 Sample Selection under Maximum Likelihood

### 2.1 Two-phase designs in fine-mapping studies

Let *Y* be the trait of interest and *G* be a (potentially causal) sequence variant located in a genomic region identified by GWAS test results. By design, variants in the region of interest are ascertained in only a fraction of individuals. Consequently, two-phase studies consist of a GWAS in phase 1 from which a subsample of individuals is selected; in phase 2, fine-mapping sequence data are collected for the subsample and combined analysis is performed using information from phases 1 & 2. In this post-GWAS setting, the trait data (*Y*) and the GWAS-SNP (*Z*), are observed for every subject in the study. The two-phase design aims to select a subset of informative subjects based on available data in the GWAS, namely (*Y, Z*). Of note, *Z* can be either an observed or imputed genotype, in the latter case the purpose might be to verify the association with sequencing data. Inference on the missing-by-design sequence variants is conducted using all available data. We define the missing indicator *R*_*i*_ = 𝟙{*i* ∈ *S*_2_}, *i* = 1, …, *N* where *N* is the number of individuals in the entire phase 1 cohort and *S*_2_ represents the set of 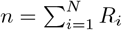 subjects selected into the phase 2 subsample. We let 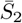 denote the set of (*N* − *n*) subjects in the GWAS study who were unselected for phase 2. We specify the selection model for the *i*th subject as *π*_*i*_(***ψ***) = *π*(*Y*_*i*_, *Z*_*i*_; ***ψ***) = Pr(*R*_*i*_ = 1|*Y*_*i*_, *Z*_*i*_; ***ψ***), where ***ψ*** is a vector that characterizes the distribution of the inclusion probabilities. To operationalize the selection, (*Y, Z*) can be stratified into *K* disjoint groups, {1}, …, {*K*}, such that *π*_*i*_(***ψ***) = *π*_*k*_(***ψ***) for all (*Y*_*i*_, *Z*_*i*_) ∈ {*k*}; that is, all subjects in the *k*th stratum have equal selection probabilities. *R*_*i*_ is designed to be conditionally independent of *G*_*i*_ given *Y*_*i*_ and *Z*_*i*_, i.e. the phase 2 selection mechanism dictated by *R*_*i*_ is completely determined by *Y*_*i*_ and *Z*_*i*_, making *G*_*i*_ missing at random^[11]^.

### 2.2 Maximum likelihood formulation

Let *f*_*β*_(*y*|*g, z*) be the parametric relationship between (*G, Z*) and *Y* indexed by ***β***. Here, *f*_*β*_(*y*|*g, z*) corresponds to a probability function in the exponential family with E[*Y* |*g, z*; ***β***] = *µ*(*g, z*; ***β***) = *h*^−1^(*β*_0_ +*β*_1_*g* +*β*_*z*_*z*), where *h*(*·*) denotes the link function. We denote 𝒢, *Ƶ* as the sets of uniquely observed values of *G* (in *S*_2_) and *Z* (in 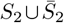). Let Pr(*G, Z*) be the joint probability function of *G* and *Z* given by the discrete probabilities *p*_*g,z*_, *g* ∈𝒢 and *z* ∈ *Ƶ*, which is left unspecified and define ***p*** = {*p*_*g,z*_}_*g*∈𝒢,*z*∈*Ƶ*_. We consider here the nonparametric estimation of the joint distribution of *G* and *Z*, with support on the Cartesian product between 𝒢 and *Ƶ*. Considering the above, we define the observed-data likelihood following previous literature^[12;13;14]^ as

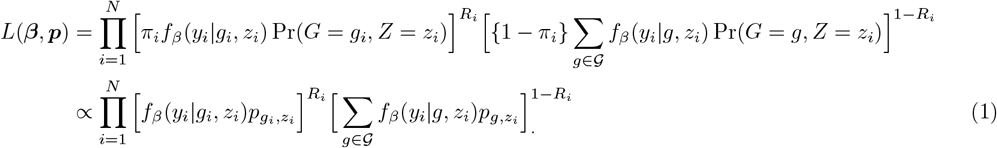

In (1), the proportionality arises since estimation of (***β, p***) does not involve *π*_*i*_’s. Thus the log-likelihood is

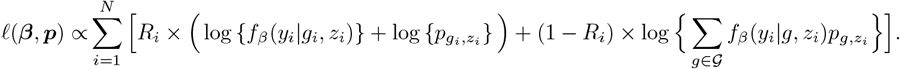

We note that additional phase 1 covariates ***X***, e.g. genomic principal components, can be introduced into the parametric model, i.e. *f*_*β*_(*y*_*i*_|*g*_*i*_, *z*_*i*_, ***x***_*i*_) with corresponding 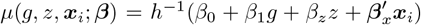, by assuming *G* and ***X*** are conditionally independent given *Z*. For simplicity, the formulation above considers *G* as a single variable, however, this can be extended to a vector with the respective considerations as illustrated in Section 4 below.

If we denote *θ* = (***β′***, ***p′***) ′, then, under regularity conditions, the limiting distribution of the maximum likelihood estimator 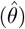 follows asymptotically 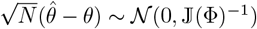 ^[15]^, where Φ = (***β′***, ***p′***, ***ψ′***) ′ and 𝕁 (Φ) is the expected information matrix, which is a function of the full parameter set Φ as the expectation is taken with respect to (*R, Y, G, Z*); note that *G* is observed in *S*_2_ and missing by design in 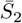 ^[8]^. The derivation of the expected information matrix is shown in Appendix A.

### 2.3 Post-GWAS analysis under maximum likelihood

Note that the likelihood in equation (1) is most useful at the design stage when no phase 2 subsample has been identified nor have any data been collected. However, once these items are available, the following re-expression is typically used:

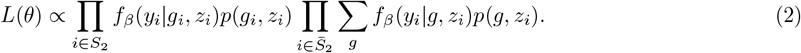

The formulation above has been amply studied^[13;14;16;17]^. Estimates can be obtained via the EM algorithm^[18;19;20]^ and the corresponding asymptotic variance covariance matrix is computed via the Louis’ method^[21]^. In fine-mapping, the aim is to identify and prioritize potential causal variants in a genetic region of interest to allow for follow-up replication and functional studies^[22]^. This can be achieved under the proposed maximum likelihood (ML) as follows: first, the genetic effect of each variant in the targeted region is estimated and tested individually (single-variant analysis); second, genetic effects are estimated and tested in multivariable models (conditional on strongest single-variant signals). Conditional analysis serves to identify independent signals in the region and to unmask associations that may have been missed in single-variant analysis. These steps are detailed in Section4.

### 2.4 Selecting phase 2 designs

In post-GWAS fine-mapping studies that target an identified genomic region, the costs of sequencing can make it unfeasible or inefficient to sequence all subjects available in phase 1, restricting the number of individuals in *S*_2_ (*n*). Here we propose two approaches to select a phase 2 design under a budget constraint and flexible optimality criteria using Lagrange multipliers or genetic algorithms. In addition, we discuss on the specification of such optimality criteria and compare the proposed methods against another class of widely used phase 2 sample selection strategies, the so-called ranked designs.

#### 2.4.1 Lagrange multipliers (LM)

Following previous ideas^[8;23;24]^, we first propose to obtain a phase 2 design for regional fine-mapping studies by minimizing the following expression

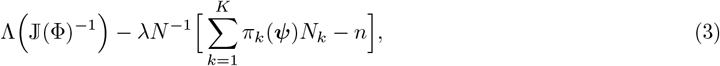

where Λ(*·*) is an optimality criterion, *λ* is a Lagrange multiplier accounting for the budget constraint and *N*_*k*_ is the number of subjects in phase 1 belonging to the *k*th stratum.

Here, we formulate the approach specifically for the ML framework described in Sections 2.2 and 2.3. For our purposes, ***β*** and ***p*** are design quantities, thus, they need to be specified *a priori* leaving the *π*s to be determined from phase 1 data alone. Note that this approach aims to find selection probabilities, 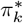, that minimize equation (3) for allocating the phase 2 sample across strata {*k*}, *k* = 1, …, *K*. The vector ***p*** can be interpreted in terms of the (joint) genotyping distribution between *G* and *Z*, which can be easily specified according to well-established genetic principles, e.g. Hardy-Weinberg equilibrium (HWE), or by external data such as the 1000 Genomes Project. Thus, the expected effect size of the sequence variant, *β*_1_, becomes the primary parameter to specify.

#### 2.4.2 Genetic algorithms (GA)

Genetic algorithms are designed to mimic nature’s evolutionary process, in which the fittest members of a population are selected to pass on their genetic information. GAs are powerful tools to optimize a fitness measure/objective function, Λ(*·*); overviews can be found in Holland^[25]^ and Whitley^[26]^. This optimization technique is suitable for a discrete solution space and is performed through a stochastic search by building an initial population of candidate solutions that evolves generationally through pairing, mating, recombining and mutating the candidate solutions. In our case, these candidate solutions correspond to vectors of the form ***R*** = (*R*_1_, …, *R*_*N*_) with 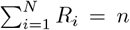 and *R*_*i*_ ∈ {0, 1} for all *i*, i.e. vectors of indicator variables denoting whether the *i*th subject is selected for phase 2. The reasoning behind GA implementation in the context of phase 2 sample selection is twofold: 1) it provides a suitable framework for discrete optimization, and 2) it has proven to be an efficient strategy to find a fittest member in large search spaces (2^*N*^ possibilities in this case)^[26]^. These appealing features of GAs come along with some challenges, namely that there are no clear convergence criteria, tuning parameters need to be specified, and they can be computationally expensive when the objective function is hard to calculate.

Nevertheless, the GA approach brings novelty to the field as it nullifies the uncertainty brought by the sampling variability introduced when utilizing stratum-specific selection probabilities. This is achieved by selecting a vector ***R***^∗^ that characterizes a unique phase 2 subsample and optimizes the fitness measure. Furthermore, GA can forgo strata definition since the search can be agnostic to specific strata configurations. In GA, the budget constraint can be introduced by a so-called cardinality constraint, which consists of selecting a subset of a required size (*n*)^[27]^. This constraint guarantees that the phase 2 sample is exactly of size *n* as opposed to methods that depend on selection probabilities, which introduce some variation into the achieved phase 2 sample size.

To date, there are several implementations for GAs available in the R statistical language^[28]^ namely packages *GA, genalg, kofnGA, mcga, mco*, and *NMOF*. Of these, only *kofnGA* is specifically designed for fixed-size subset selection with a flexible specification of the objective/fitness function, see Wolters for a detailed explanation of the package^[29]^. In any GA, it is important to consider the set of control parameters necessary to implement decision rules at each step. Specifically, *kofnGA* requires the user to specify the objective/fitness function (i.e. Λ(*·*)), the subset size (*n*), and the number of candidates (*N*) while additional control parameters related to algorithmic design have default (but adjustable) settings. These control parameters are: *population size* (ℳ), *number of generations* (ℋ), *size of selection tournament* (𝒯), *mutation rate* (𝓇), and *number of elites* (ℰ). A pseudo-algorithm relating standard GA terminology with the two-phase design along with a description of the steps where the control parameters are used is presented in Algorithm 1.

##### Algorithm 1

Pseudo-algorithm of the implemented genetic algorithm

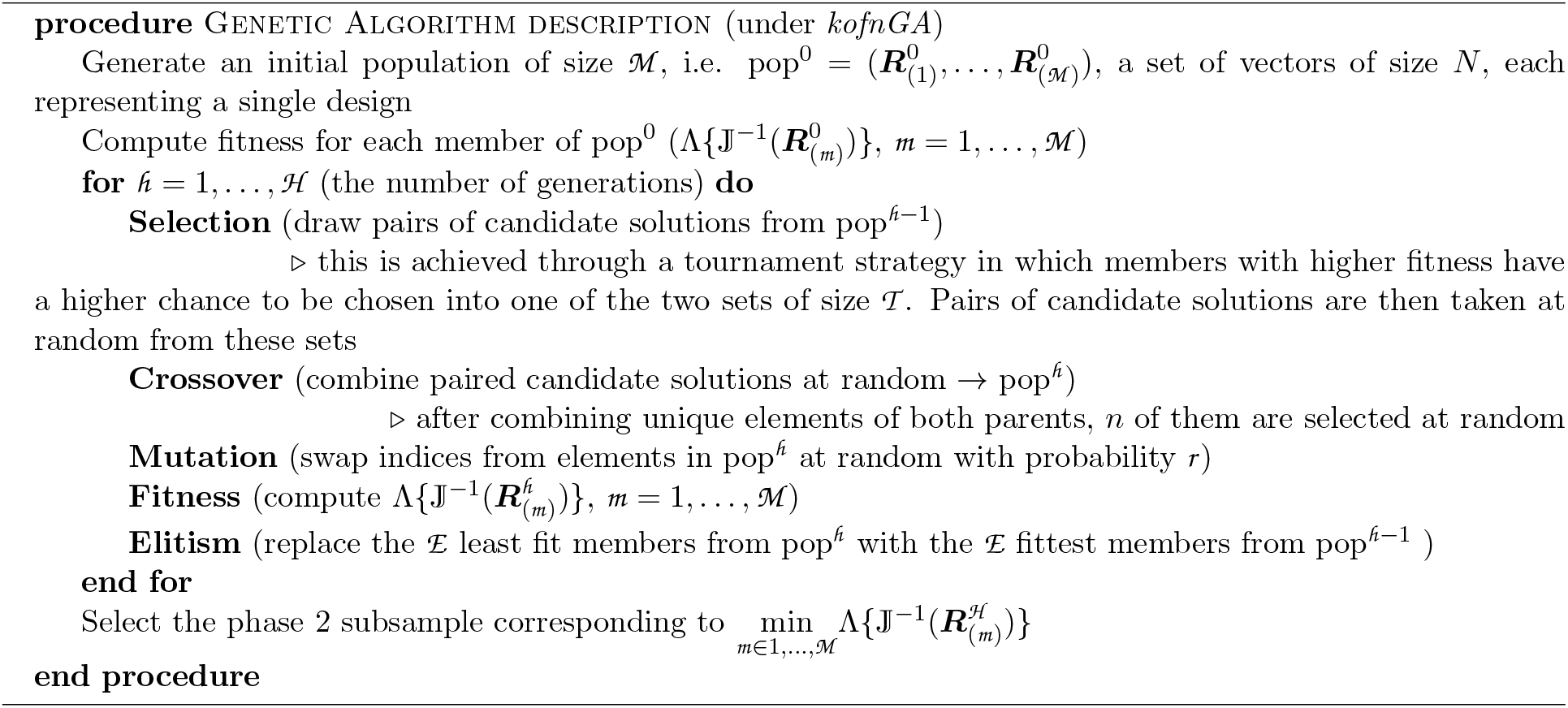

The population is the pool of candidate solutions at each iteration from which the fittest members, i.e. members with optimal Λ(*·*) values, will be ultimately selected. The number of generations denotes the number of iterations the algorithm will run for. The size of selection tournament determines the number of members of the population selected to produce the next generation. The mutation rate determines the probability at which random swaps in the indexes of the candidate solutions occur in the population. Lastly, elites are the fittest members in a given generation that get to be kept in the next generation.

The algorithm parameters can be tuned by the user in accordance with the problem at hand. For simplicity, parameters ℋ and ℰ can be reformatted as proportions of the population size (ℳ). Because *kofnGA* does not implement a stopping rule, the algorithm iterates for as many times as specified by the provided number of generations (ℋ). To accelerate convergence, we set 𝒯 and ℰ at high levels (= 0.90 × ℳ) as suggested by Walters^[29]^. This approach may diminish the search improvements derived from mutations, relying more heavily on the initial population and number of generations. Therefore, instead of setting a completely random initial population (pop^0^ in Algorithm 1), we initialize it with an equal number of samples with top 20 performers (based on Λ(*·*)) out of 100 draws of each of the balanced, combined, and LM sampling designs plus the RDS design. This strategy guarantees that GA has at least the same performance as the RDS design.

Additional considerations for the optimization strategies in LM and GA as well as further details on the balanced and combined designs can be found in Sections S2 and S3, respectively (Online Supplementary Material). It is also worth noting that the proposed approaches are feasible in any context where *Y* |*G, Z* can be modeled within the exponential family, and both *G* and *Z* are discrete (or easy to discretize) covariates.

#### 2.4.3 Specifying an optimality criterion

There are several ways to define a functional Λ(*·*) as the optimality criterion/fitness measure, mostly grounded in experimental design^[30]^. In this report, we explore three criteria: A-optimality, D-optimality and parameter-specific. Each criterion focuses on different features of the variance-covariance matrix (VCM), 𝕁 (Φ)^−1^ (Table 1). The parameter-specific criterion is optimal to identify designs with minimum variance when testing a single parameter. Similarly, A- and D-criteria would be optimal to identify designs with minimum average variance across all parameters and minimum volume of the confidence ellipsoid, respectively.

**Table 1:** Description of the three optimality criteria evaluated. 𝕁 (Φ)^−1^ denotes the variance-covariance matrix.

| $\Lambda(\cdot)$ | Formula | Description |
| --- | --- | --- |
| A-optimality | $\sum \text{diag}(\mathbb{J}(\Phi)^{-1})$ | minimizes the average variance of the parameter estimates |
| D-optimality | $\det(\mathbb{J}(\Phi)^{-1})$ | minimizes the product of the variances for diagonal matrices |
| Parameter-specific | $\mathbb{J}(\Phi)^{-1}_{[\beta_1, \beta_1]}$ | minimizes the variance of a particular entry in the VCM |

In the outlined post-GWAS setting, the focus lies on testing a single parameter, *β*_1_. Thus, the parameter specific criterion is the most natural choice. However, when multiple parameters are of interest, i.e. *β*_1_ is a vector, A-optimality may be preferred if the parameters of interest are loosely correlated, whereas D-optimality may be preferred when the parameters are strongly correlated. This makes intuitive sense when there are two (or more) estimators, for example, 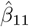 and 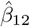 but 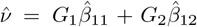 is of interest. Then 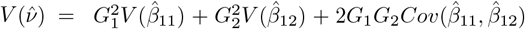, thus, involving off-diagonal elements of the VCM. We evaluate potential differences in the choice of optimality criterion in Section 4.

#### 2.4.4 Ranked designs

Recently, Tao *et al*. proposed general optimal designs for phase 2 studies^[10]^. In this section, we aim to describe their approach, summarize their findings, and draw comparisons with the proposed designs: LM and GA.

Despite their names, the outcome-dependent sampling (ODS) and residual-dependent sampling (RDS), as defined by Tao *et al*., are not sampling designs in the classical sense given the fact that their specification is independent of any sampling mechanism^[10]^. Indeed, this is also true for the TZL design. We refer to them as ranked designs because they are defined in terms of ordered quantities: outcome/residuals/scaled residuals for ODS, RDS, and TZL, respectively. Tao *et al*. show that the scaling factor in TZL is given by Var(*G*|*Z*)^1*/*2^, which is unknown at design stage and thus needs to be specified prior to phase 2. An intuition on why this scaling factor is important for the optimal design, provided by Tao *et al*., is that *G* is harder to be retrieved by *Z* when Var(*G*|*Z*) is large and thus these subjects need to be oversampled^[10]^.

The ranked designs achieve a given phase 2 sample size by selecting an equal number of subjects from each of the top and bottom rankings of the outcome/residuals/scaled residuals. This particularity makes them appealing for a few reasons: 1) the ranked designs are unique, 2) the selection is intuitive and can be performed quickly, and 3) for QTs, no stratification on the outcome is required. Tao *et al*. show that the TZL design reduces to the RDS design when *G* and *Z* are independent (and RDS reduces to ODS when *Y* and *Z* are independent); in Sections 4 and 5, we investigate the effect of misspecifying Var(*G*|*Z*).

There are five main underlying differences between the proposed designs (LM and GA) and the ranked designs, particularly TZL:

i. LM depends on the stratification strategy undertaken for the outcome while none of the ranked design depends directly on outcome stratification for QTs. On the other hand, GA can, in principle, be performed without defining any stratification, however, selection of initial values may depend on values drawn from LM or other designs to accelerate convergence, which could introduce some dependency on a chosen stratification.
ii. LM provides optimal sampling fractions and thus a sample must be drawn accordingly, subjecting this design to sampling variability. GA avoids this by selecting a unique solution, ***R***^∗^, with optimal Λ(*·*) value. However, given the stochastic nature of the genetic algorithm, this solution is approximate and varies at each run unless a random seed is specified. In contrast, the ranked designs are not subject to sampling variation.
iii. TZL and the proposed designs (under the parameter-specific criterion) seek to minimize the variance of *β*_1_. In the case of TZL, this is achieved by maximizing the inverse of the efficiency bound for estimating *β*_1_ with one observation in their Theorem 1. Note that the proof of this theorem relies on the assumption that *Y* and *G* are approximately independent given *Z*, which is justified when the effect of *G* on *Y* is small, i.e. *β*_1_ = *o*(1). LM and GA, on the other hand, do not depend on the small *β*_1_ assumption and can be thus implemented in more general settings.
iv. Related to the point above, LM and GA rely on an empirical approximation to the information matrix whereas the variance considered in TZL uses an exact expected information under the working assumption of *β*_1_ = *o*(1). This defines a trade-off between the generality of LM and GA and the increase in efficiency of TZL when the assumption is justified.
v. LM and GA can optimize general functions beyond 𝕁 (Φ)^−1^ through Λ(*·*), whereas the results in TZL are mostly concerned with 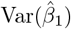 (or functions thereof).

It remains unclear what constitutes a good stratification for LM; intuitively, LM should approximate TZL as the number of strata approaches the phase 1 sample size. However, a theoretical proof is beyond the scope of this paper. To circumvent the sampling variability issue in LM, one could draw a predetermined number of subsamples and select the one with optimal Λ(*·*) value. Regarding whether restricted/unrestricted values of *θ* are preferred, Tao *et al*. show that TZL performs well for alternatives close to the null. Although these alternatives are typical for genetic studies, a more comprehensive comparison for alternatives farther away for the null is warranted. Lastly, LM and GA can, in fact, approximate the optimization strategy in TZL by utilizing 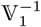 instead of 𝕁 (Φ)^−1^ in the objective function, where 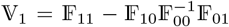 and 𝔽 = 𝕁 (Φ) is partitioned with respect to *β*_1_ as 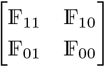. It is worth noting that the dimension of 𝕍_1_ corresponds to that of the subspace determined by the null hypothesis of interest. In the simplest case of *β*_1_ being a scalar, 𝕍_1_ is also a scalar. In LM and GA higher dimensions can be easily accommodated by specifying a different Λ(*·*) on 𝕍_1_, to obtain say A- or D-optimal designs.

## 3 Simulation Studies

In this section, we describe the data generation steps, analysis plan, and report the results of an initial set of simulations. The main objective is to compare the statistical power of the proposed phase 2 designs, LM and GA, in a post-GWAS fine-mapping scenario by testing for the effect of *G* (the missing-by-design variable), i.e. H_0_ : *β*_1_ = 0. In addition, we compare LM and GA against two ranked designs: TZL and RDS. Of note, we exclude ODS from these studies given the known indirect association between *Z* and *Y* at GWAS stage. Estimates and standard errors are constructed following equation (2) in Section 2.3. For comparability with RDS and TZL, we use 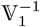 in the parameter-specific optimality criterion for LM and GA as described in Section 2.4.4 given that this variance estimate does not depend on the assumption of *β*_1_ = *o*(1). Additional numerical studies comparing LM and GA against alternative heuristic designs utilizing 𝕁 (Φ)^−1^ in the optimization are found in Section S3, Online Supplementary Material.

### 3.1 Data generation

We assume a data generating mechanism similar to Espin-Garcia *et al*. ^[17]^. Briefly, for a phase 1 sample size (*N*), and given values for minor allele frequencies (MAFs), *q*_*G*_ and *q*_*z*_ and the linkage disequilibrium (LD), quantified through the Pearson correlation coefficient, *r*, we simulate two variants on the same haplotype under Hardy-Weinberg equilibrium (HWE): *G*_1_ and *Z*. Here, *q*_*G*_ and *q*_*Z*_ are the frequencies of the less common allele in the population for *G*_1_ and *Z*, respectively, whereas LD is the level of correlation between them. Notably, since the actual allele frequencies cannot be negative and the additive linkage disequilibrium coefficient D is constrained, not all combinations of *r, q*_*G*_ and *q*_*Z*_ can occur.

The trait of interest is then generated as *Y* = *β*_0_ + *β*_1_*G*_1_ + *ε*, where *ε* ∼𝒩 (0, *σ*^2^). We note that in this setting, as opposed to Section 2.2, *Z* is assumed to be conditionally independent of *Y* given *G*. This simulation setup aims to resemble a more realistic scenario in which the GWAS-SNP, *Z*, is not causal itself but rather is in linkage disequilibrium with the causal variant, *G*.

To imitate the GWAS setting in each dataset, we test *γ*_*z*_, the genetic effect of *Z*, for association in the regression model *Y* = *γ*_0_ +*γ*_*z*_*Z* and only keep replicates that meet a suggestive genome-wide significance criterion of *p <* 1 × 10^−5^ for the hypothesis H_0_ : *γ*_*z*_ = 0. Lastly, to study type 1 error (T1E) under this data-generation mechanism, we simulate another SNP, *G*_0_ independently from *Z* and *G*_1_ with MAF *q*_*G*_.

Strata for *Y* are defined by discretizing the trait values into a three-category variable, *Y*_*st*_ = {*T*_1_, *T*_2_, *T*_3_}, according to fixed cut points (*C*1, *C*2) as the percentiles (40,60) of a normal distribution with mean *µ* = 2 and variance *σ*^2^ = 1, so that under the null, Pr(*Y < C*1) = Pr(*Y > C*2) = 0.4. Strata for the biallelic GWAS SNP, *Z*, are defined by considering *Z* as a three-category variable corresponding to genotypes, (*AA, Aa, aa*) and coded by the number of copies of the minor allele (*a*), i.e. *Z* = 0, 1, 2 (additive association).

Of note, stratification by *Y*_*st*_ and *Z* is only employed for optimization in LM and for visualization to compare the distribution of selected individuals under other designs.

### 3.2 Assessing the phase 2 designs

The first set of evaluations consists of the following. We specify a phase 1 sample size of *N* = 5000 and a phase 2 sample size of *n* = 540, 1500, 2500, i.e. 0.108, 0.25, and 0.50 of the phase 1 data, respectively. We draw 1250 replicates for each combination of simulation parameters *q*_*G*_ = 0.2, *q*_*Z*_ = 0.3, *r* = 0.75, *β*_0_ = 2, *σ*^2^ = 1, and *β*_1_ ∈ ⟨0.1 + 0.2*j*|*j* = 0, …, 3⟩. We evaluate the performance of the the proposed designs against two ranked designs, RDS and TZL, across three statistical tests (Wald, likelihood ratio (LR) and score). Since *β*_1_ is a scalar, the comparison against ranked designs only examines a parameter-specific criterion as a consequence from considering 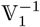 in the objective function, as mentioned in Section 2.4.4.

To compare power, we assess the ratio of the empirical power of each design over that of the complete data case (relative empirical power, rEP). In addition, estimation efficiency is compared via relative asymptotic and empirical standard error (rASE and rESE, respectively) of 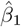 for each design over that of the complete data. We deem these measures provide a better reflection of the design performance compared to studies that benchmark against simple random sampling. Note that as these ratios become closer to 1 (100%), the better the studied designs are able to recover the performance of the complete data analysis.

The specification of ***β***_*des*_, the design regression parameters, corresponds to 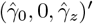. Here 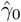 and 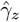 denote the maximum likelihood estimates (MLEs) from GWAS, i.e. the MLEs for *Y* = *γ*_0_ + *γ*_1_*Z*. Similarly, we specify ***p***_*des*_, the design haplotype distribution between *G* and *Z*, under HWE by estimating *q*_*Z*_ from the phase 1 sample and designating *q*_*G*_ and *r* to be the equal to their generating values. These design values are used to determine LM, GA, and TZL designs but not RDS, which is agnostic to these design quantities.

Although correct specification of the design quantities is hardly ever attainable, the settings above allow us to evaluate the true type 1 error/power of the studied designs. We discuss in the next section how to proceed in practice when the true design values are unavailable. Moreover, by specifying the regression parameters under the null hypothesis, i.e. 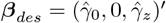, the design problem greatly simplifies to only specify values for ***p***_*des*_.

### 3.3 Results

The ranked designs can be specified without strata definitions, however, for visualization and comparison purposes, we plot the distribution of RDS and TZL according to the predetermined strata. When comparing LM and GA against the ranked designs for the smallest and largest studied genetic effects (*β*_1_ = 0.1 and *β*_1_ = 0.7), we observe: (1) LM, GA and TZL vary considerably across values of *β*_1_ and *n*, (2) LM displays more unstable strata distribution when compared against GA and ranked designs, especially for the smaller phase 2 sample sizes (*n* = 540, 1250), (3) GA follows closely the RDS design especially when *n* = 540, 1250, (4) LM and GA reach an approximately equal strata distribution when *n* = 2500, and (5) as expected, the strata distribution of the RDS design remains practically unchanged between genetic effect sizes and phase 2 sample sizes (Figure 1).

**Figure 1:**
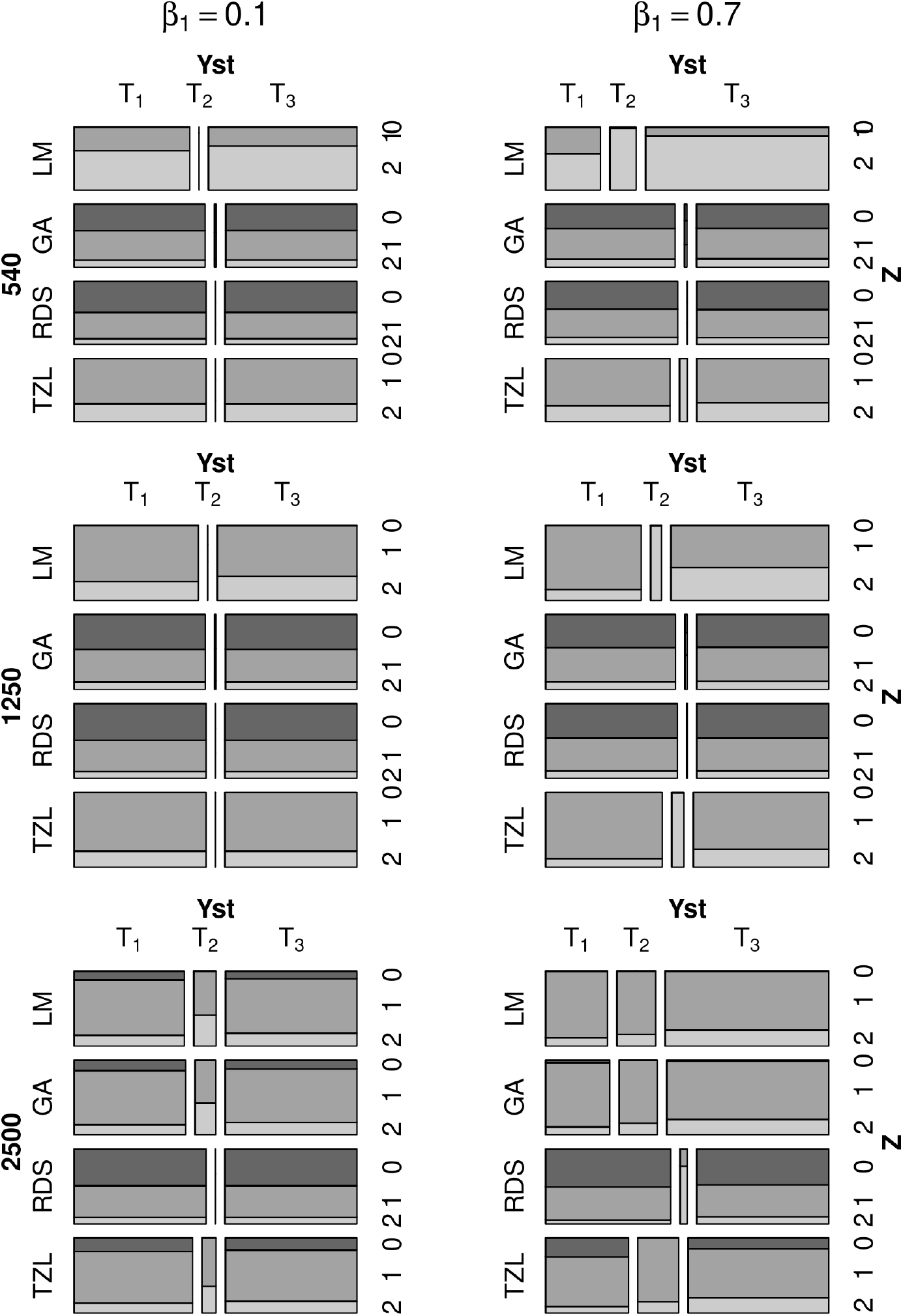
Mosaic plots with the average strata sizes across replicates for the proposed designs against ranked designs across phase 2 sample sizes, *n* = 540, 1250, 2500, under the parameter-specific criterion. Averages were taken from the resulting designs in the main simulation study for the two most extreme values of *β*_1_ (0.1,0.7).

At *α* =1%, type 1 error (T1E) rates demonstrate well controlled values across the three tests in most cases. For LM, slightly anti-conservative T1E rates under the Wald test and conservative rates under the score test are observed when *n* = 540 and approximate the nominal rate as *n* increases(Table 2). Closer inspection of the *p*-value distribution under LR displays no gross departure from the expected uniform distribution (Figure S1). Overall, we observed the LR statistics showed better behavior compared to score and Wald statistics even under small sample sizes specially under LM design (Table 2). Empirical bias 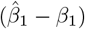 is well centered around zero overall and decreases as *n* increases for all designs when the true value for *β*_1_ is small (*<* 0.3) (Figure 2). However, for larger values of *β*_1_ (≥ 0.5), LM and TZL show biased estimates when *n* = 540, 1250 and deteriorate as *β*_1_ increases (Figure 2). All designs show relatively close agreement between (r)ASE and (r)ESE across values of *β*_1_ and *n* (Tables S1-S2). TZL shows values of rASE and rESE closer to 1, with GA second, RDS third, and LM coming last. GA, RDS designs exhibit adequate coverage while the coverage for LM and TZL worsens as *β*_1_ increases for *n* = 540, 1250 (Table S3).

**Table 2:** Type 1 error (T1E) (*α* = 1%) across studied designs, phase 2 sample sizes (*n* = 540, 1250, 2500) and statistical tests under a parameter-specific criterion. Each entry represents 11250 replicates pooled across empirical null scenarios. The rest of the simulation parameters correspond to *q*_*G*_ = 0.2, *q*_*Z*_ = 0.3, *r* = 0.75, *β*_0_ = 2, *σ*^2^ = 1, *N* = 5000. The complete data T1E is 1.16 for Wald/LR tests and 1.14 for the score test. To further evaluate test validity under the studied sample sizes, we plot histograms of the observed LR test *p*-values in Figure S1.

| $n$ | Test | LM | GA | RDS | TZL |
| --- | --- | --- | --- | --- | --- |
| 540 | Wald | 1.45 | 1.17 | 1.10 | 1.01 |
|  | LR | 1.02 | 1.14 | 1.10 | 0.97 |
|  | Score | 0.94 | 1.10 | 1.07 | 0.95 |
| 1250 | Wald | 1.06 | 1.15 | 1.12 | 1.02 |
|  | LR | 0.94 | 1.14 | 1.12 | 0.99 |
|  | Score | 0.90 | 1.13 | 1.10 | 0.96 |
| 2500 | Wald | 1.00 | 1.08 | 1.08 | 1.09 |
|  | LR | 0.97 | 1.06 | 1.07 | 1.07 |
|  | Score | 0.96 | 1.05 | 1.07 | 1.06 |

**Figure 2:**
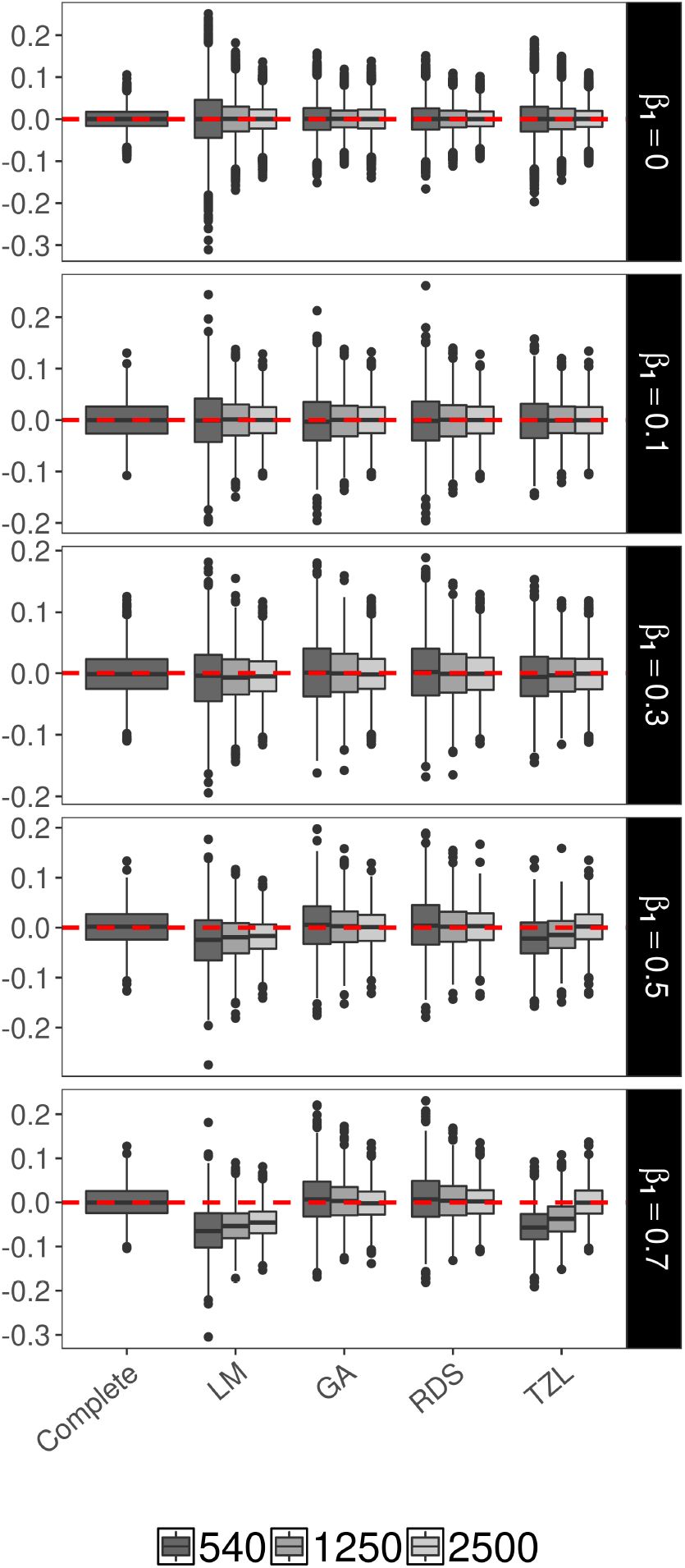
Boxplots for the distribution of the bias across genetic effect estimates 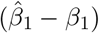 in the studied designs under a parameter-specific criterion. Row facets denote different true *β*_1_ values (0, 0.1, 0.3, 0.5, 0.7).

Power curves under the LR test at *α* = 1 × 10^−8^ level, show that TZL consistently demonstrates the highest power across values of *n* with GA second, RDS in the third place and LM having the lowest power. Notably, all designs reach similar power when *n* = 2500 (Figure S2). Interestingly, not all methods show power increases at the same rate due to the differences in efficiency across designs. Additional simulations for larger phase 1 sample size (*N* = 10000) and similar selection fractions (*n/N* = 0.10, 0.25, 0.50) result in analogous type 1 error and power results among designs, suggesting that testing performance is contingent upon sampling fraction and not phase 2 sample size (Section S4.1, Online Supplementary Material). Under the LR test, the rEP is highest for the TZL across values of *β*_1_ and *n*. GA shows higher power than RDS across virtually all scenarios while LM comes last when *n* = 540, 1250 but reaches similar power to GA when *n* = 2500 (Table 3).

**Table 3:** Relative empirical power (rEP), calculated as the ratio of the empirical power of each studied design over that of the complete data, across studied designs, phase 2 sample sizes, and effect sizes under the LR test (*α* = 1 × 10^−8^). Phase 1 sample size is *N* = 5000 whereas phase 2 sample size is *n* = 540, 1250, 2500. These results exclude *β*_1_ *>* 0.5 since power had already reached 100%. The rest of the simulation parameters correspond to *q*_*G*_ = 0.2, *q*_*Z*_ = 0.3, *r* = 0.75, *β*_0_ = 2, and *σ*^2^ = 1.

| $n$ | $\beta_1$ | LM | GA | RDS | TZL |
| --- | --- | --- | --- | --- | --- |
| 540 | 0.100 | 0.0 | 0.0 | 100.0 | 0.0 |
|  | 0.125 | 12.5* | 12.5* | 12.5* | 0.0 |
|  | 0.150 | 0.0 | 4.2 | 6.2 | 12.5 |
|  | 0.175 | 1.1 | 5.6 | 3.9 | 19.7 |
|  | 0.200 | 2.2 | 3.8 | 3.6 | 23.3 |
|  | 0.225 | 2.9 | 9.2 | 6.7 | 32.5 |
|  | 0.250 | 4.5 | 14.8 | 11.7 | 40.4 |
|  | 0.300 | 15.0 | 38.7 | 35.5 | 74.2 |
|  | 0.400 | 69.2 | 94.2 | 91.8 | 99.6 |
|  | 0.500 | 97.1 | 100.0 | 99.9 | 100.0 |
| 1250 | 0.100 | 0.0 | 0.0 | 0.0 | 0.0 |
|  | 0.125 | 25.0* | 37.5* | 37.5* | 87.5* |
|  | 0.150 | 14.6 | 18.8 | 18.8 | 54.2 |
|  | 0.175 | 19.7 | 30.9 | 27.0 | 71.3 |
|  | 0.200 | 23.8 | 32.5 | 29.4 | 67.5 |
|  | 0.225 | 36.0 | 44.1 | 45.5 | 82.1 |
|  | 0.250 | 44.4 | 57.5 | 56.0 | 87.4 |
|  | 0.300 | 77.0 | 85.7 | 84.9 | 97.0 |
|  | 0.400 | 99.6 | 99.9 | 99.9 | 100.0 |
|  | 0.500 | 100.0 | 100.0 | 100.0 | 100.0 |
| 2500 | 0.100 | 100.0* | 100.0* | 100.0* | 100.0* |
|  | 0.125 | 137.5* | 112.5* | 75.0* | 100.0* |
|  | 0.150 | 72.9 | 72.9 | 70.8 | 91.7 |
|  | 0.175 | 94.9 | 94.9 | 75.3 | 95.5 |
|  | 0.200 | 89.0 | 89.0 | 72.4 | 91.3 |
|  | 0.225 | 96.0 | 96.1 | 86.4 | 95.9 |
|  | 0.250 | 96.8 | 97.1 | 90.8 | 98.6 |
|  | 0.300 | 99.3 | 99.2 | 98.2 | 99.6 |
|  | 0.400 | 100.0 | 100.0 | 100.0 | 100.0 |
|  | 0.500 | 100.0 | 100.0 | 100.0 | 100.0 |

\* Non-consistent improvement due to low power in complete data.

Besides the additional simulations on different phase 1 and 2 sample sizes, we also studied the influence of different specifications of the joint distribution of *G* and *Z*. In summary, these results are analogous to what was reported above with GA showing competitive power when compared against alternative designs (Section S4.2, Online Supplementary Material).

## 4 Two-phase Study Design in Practice

For LM, GA and TZL designs, specifying different design quantities, 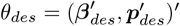, will lead to different phase 2 subsamples. Little attention has been paid to the practical considerations entailed in choosing a study design. One practical strategy is to make an educated guess for the design quantities; another is to consider a range of plausible values. Though adaptive/sequential designs may be feasible in some circumstances^[31]^, in the post-GWAS setting processing data by batch may be operationally inefficient. In addition, although the sequential strategy will provide more precise design parameters, it will not necessarily aid in solving the withstanding issue of selecting a unique phase 2 sample given that multiple more precise estimates will be potentially identified. Therefore, we propose a strategy that relies only on phase 1 data to select a unique phase 2 sample when a range of effect sizes, allele frequencies, and LD values can be considered at design stage.

### 4.1 A grid search procedure to select a unique phase 2 subsample

It is likely that there will be uncertainty about the specification of the effect size (***β***_*des*_) and haplotype distribution (***p***_*des*_) at design stage, so we must consider a range of probable values and define a grid of intermediate points inside this range. Let {*θ*_*h*_}, *h* = 1, …, *H* be the set of probable values or design quantities of interest. Each design quantity *θ*_*h*_ will yield an optimal phase 2 subsample, P2S^(*h*)^, for the second stage. Thus, to select an unique design under the set {*θ*_*h*_}_*h*=1,…,*H*_ we propose the following procedure, which is motivated by robustness considerations.

1. Given an optimality criterion, Λ(*·*), for each *h*
  - obtain a phase 2 subsample, namely P2S^(*h*)^, via LM/GA or otherwise by optimizing Λ(*θ*_*h*_).
  - given P2S^(*h*)^, calculate 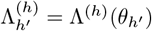 for *h′* = 1, …, *H*
  - compute 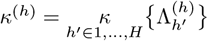, where *κ* is a summary function, e.g. mean or median
2. select the P2S^(*h*)^ with minimum *κ*^(*h*)^

This procedure will identify a unique design from the ones generated using alternative specifications {*θ*_*h*_}. To better understand the proposed procedure, let us assume that we are interested in comparing two designs, PS2^(1)^ and PS2^(2)^ which are optimal when the design values are *θ*_1_ or *θ*_2_, respectively. In order to select the best design, we adopt a criterion based on robustness. In other words, we are interested in determining which one of these two designs exhibits an overall superior performance when *θ* differs from its generating design value. To this end, we compute the fitness function for each design and each *θ*_*h′*_ with *h′* ≠ *h* and select the design that achieves the best average (or median) performance. The formal description simply extends this principle to comparing *H* designs and selecting the most robust one. Ultimately, regional sequencing data will be collected for the subsample from the resulting design alone. Once data are collected, statistical fine-mapping can be conducted following the analytic strategy described in Section 2.3.

### 4.2 Simulation under realistic LD patterns

The purpose of this simulation study is to evaluate the studied designs when the values of *θ* = (***β′, p′***) ′ are unknown and a range of values for *θ*_*des*_ is considered instead. In this study, we generate data under a scenario where multiple “causal variants” and a realistic LD structure from a targeted region were considered. Specifically, we select four loci in chromosome 16 as causal variants, ***G*** = (*G*_1_, …, *G*_4_) with corresponding effect sizes 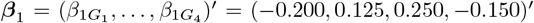 in hg19 positions 56989830, 56993324, 56994990, 56995236, and designate rs247617 (pos. 56990716) as the GWAS SNP (hereafter all positions are truncated to the last 5 digits). We then generate a QT, *Y*, across 500 replicates following 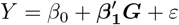, where *β*_0_ = 2, and *ε ∼*𝒩 (0, *σ*^2^ = 1). Details of the data generation are provided in Section S5, Online Supplementary Material.

### 4.3 Selecting phase 2 samples under prespecified sets of design quantities

Since multiple causal variants are assumed in this section, we ascertain the performance of the studied designs under alternative optimization criteria in addition to the parameter-specific criterion, specifically under A- and D-optimality. This allows *β*_1_ to be treated as a vector at design stage, that is 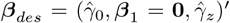, with **0** being a zero vector. As before, 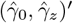 correspond to the MLEs of the regression model *Y* = *γ*_0_ + *γ*_*z*_*Z*, i.e. GWAS MLEs. In the simulated data, 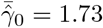 range (1.68 − 1.78), 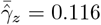 range (0.097 − 0.176) across the 500 replicates.

As mentioned in Section 2.4.4, it is straightforward to modify the optimality criteria for LM and GA. For TZL, no specific results were provided under alternative optimality criteria. However, Tao *et al*. discussed that as Var(***G***|*Z*) becomes a matrix when ***β***_1_ becomes a vector, it was sufficient to replace the scaling factor Var(*G*|*Z*)^1*/*2^ (when *G* is a scalar) with Λ[Var(***G***|*Z*)]^[10]^.

Additionally, for LM and GA, the optimization is performed using 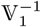, as this approach showed best performance in the first simulation study when the true values are close to the null hypothesis (Section 3.2). A unique design is then selected for LM, GA and TZL considering multiple (mispecified) values of ***p***_*des*_ following 4.1. We also considered RDS in this simulation study, however, since RDS does not depend on *θ*_*des*_ in any way, it was not determined using the outlined procedure 4.1. For each replicate, we select a phase 2 data of size *n* = 1250, 2500. The specification of ***p***_*des*_ under parameter-specific, A-, and D-optimality criteria is described below.

#### 4.3.1 Parameter-specific criterion

Under this criterion, *β*_1_ is a scalar. Thus, we specify ***p***_*des*_ = *P* (*G, Z*) = *p*_*gz*_ assuming each of the resulting combinations between the following:

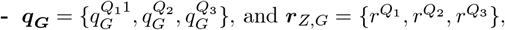

where *Q*_1_, *Q*_2_, *Q*_3_ denote the first, second, and third quartiles of *q*_*G*_ (MAF) or *r*_*Z,G*_ (LD between *Z* and *G*) across the 29 sequence variants in the region, e.g. 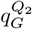 is the median MAF value across seq-SNPs in the fine-mapped region while 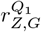 denotes the 25th percentile across correlation values between the GWAS-SNP, *Z*, and the seq-SNPs, *G*.

#### 4.3.2 A- and D- optimality criteria

Under these criteria, ***β***_1_ is assumed to be a vector of size 2. Thus, we specify 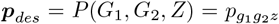 assuming each of the resulting combinations between the following:

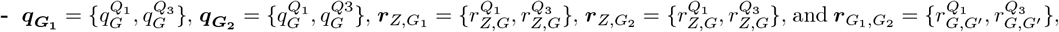

as before, *Q*_1_ and *Q*_3_ denote the first and third quartiles of *q*_*G*_ (MAF), *r*_*Z,G*_ (LD between *Z* and *G*) or *r*_*G,G′*_ (LD between *G* and *G′*) across the 29 sequence variants in the region.

In the simulated data, 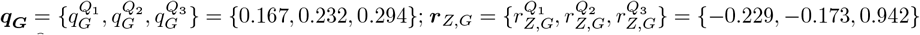; and 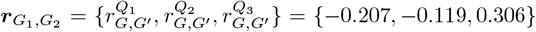. The average sample distribution of the resulting phase 2 designs across the 500 replicates is portrayed via mosaic plots (Figures S4 and S5). GA, RDS, and TZL designs show rather similar distributions across optimality criteria especially when *n* = 2500. LM, on the other hand, selects only from the extremes of the distribution for common heterozygous (*Z* = 0) for *n* = 1250. In addition, the intersection of the subsamples taken across each design/optimality criterion combination is presented via upset plots for a single replicate (Figures S3 and 3). These plots show that almost a third of the phase 2 subsamples are common among GA, RDS and TZL designs and optimality criteria when *n* = 1250. Notably, the number of common subsamples jumps to about a half among GA, RDS and TZL and optimality criteria when *n* = 2500.

**Figure 3:**
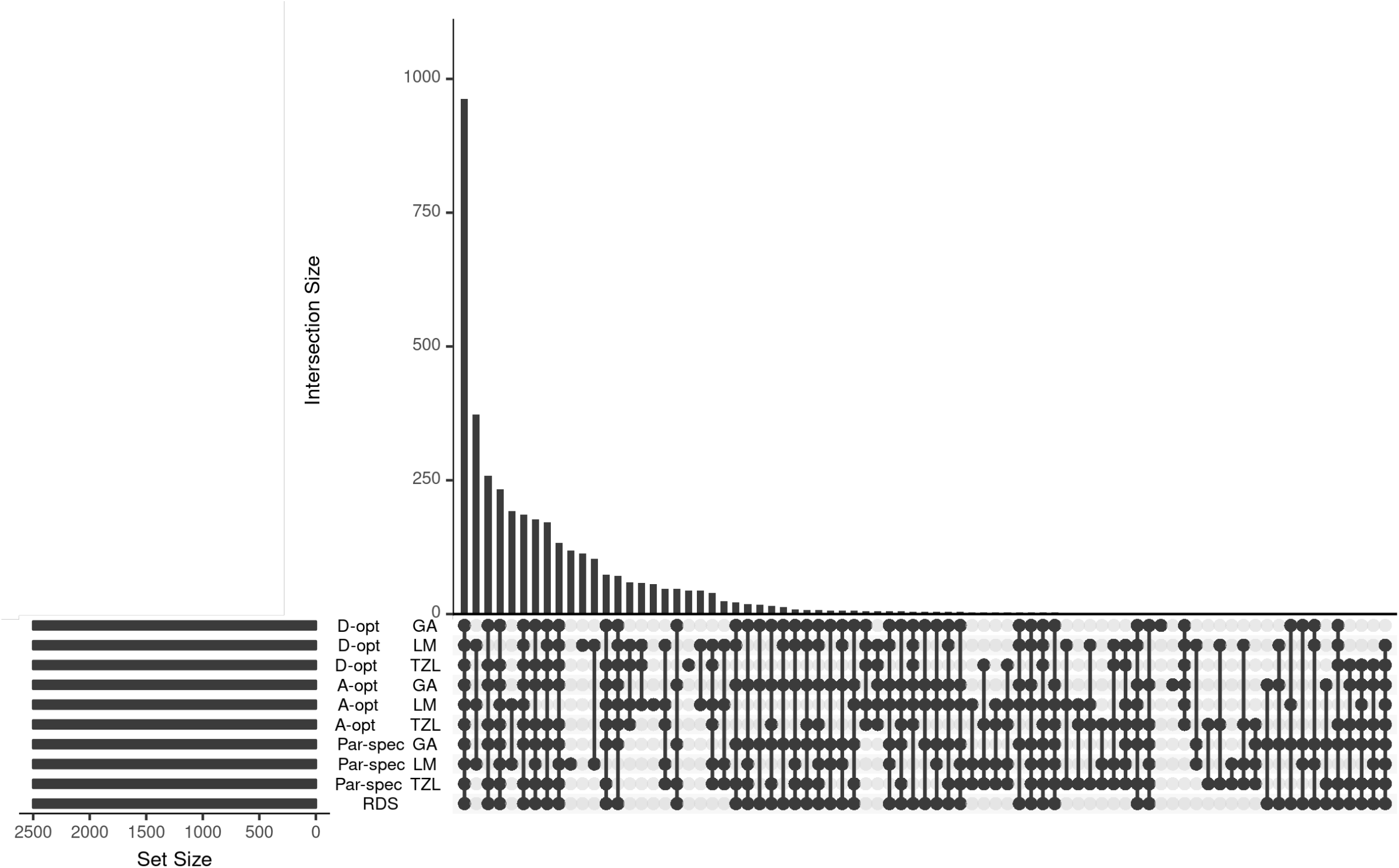
Upset plot for a single replicate in the realistic simulation to quantify the intersection sizes across studied designs and optimality criteria when *n* = 2500. Each bar denotes the size of a given intersection highlighted in the x-axis, i.e. the number of subjects common among designs. The matrix in the x-axis corresponds to each optimality criterion (parameter-specific/A-/D-optimality) and design (LM/GA/TZL) combination as well as RDS.

### 4.4 Single-variant fine-mapping analysis

Once the phase 2 sample is selected in a given replicate, we perform a region scan using the 29 variants, i.e. we test for association one variant at a time, across each phase 2 sample size (*n* = 1250, 2500). To decrease the collinearity between *G* and *Z* in the model, we treat the GWAS SNP, *Z* (rs247617), as a (3-level) categorical covariate at design and analysis stages. We summarize the point estimates, asymptotic standard errors, and empirical power rates (under the LR test) for the region scans across replicates for each design and optimality criteria (Tables S4-4).

**Table 4:**
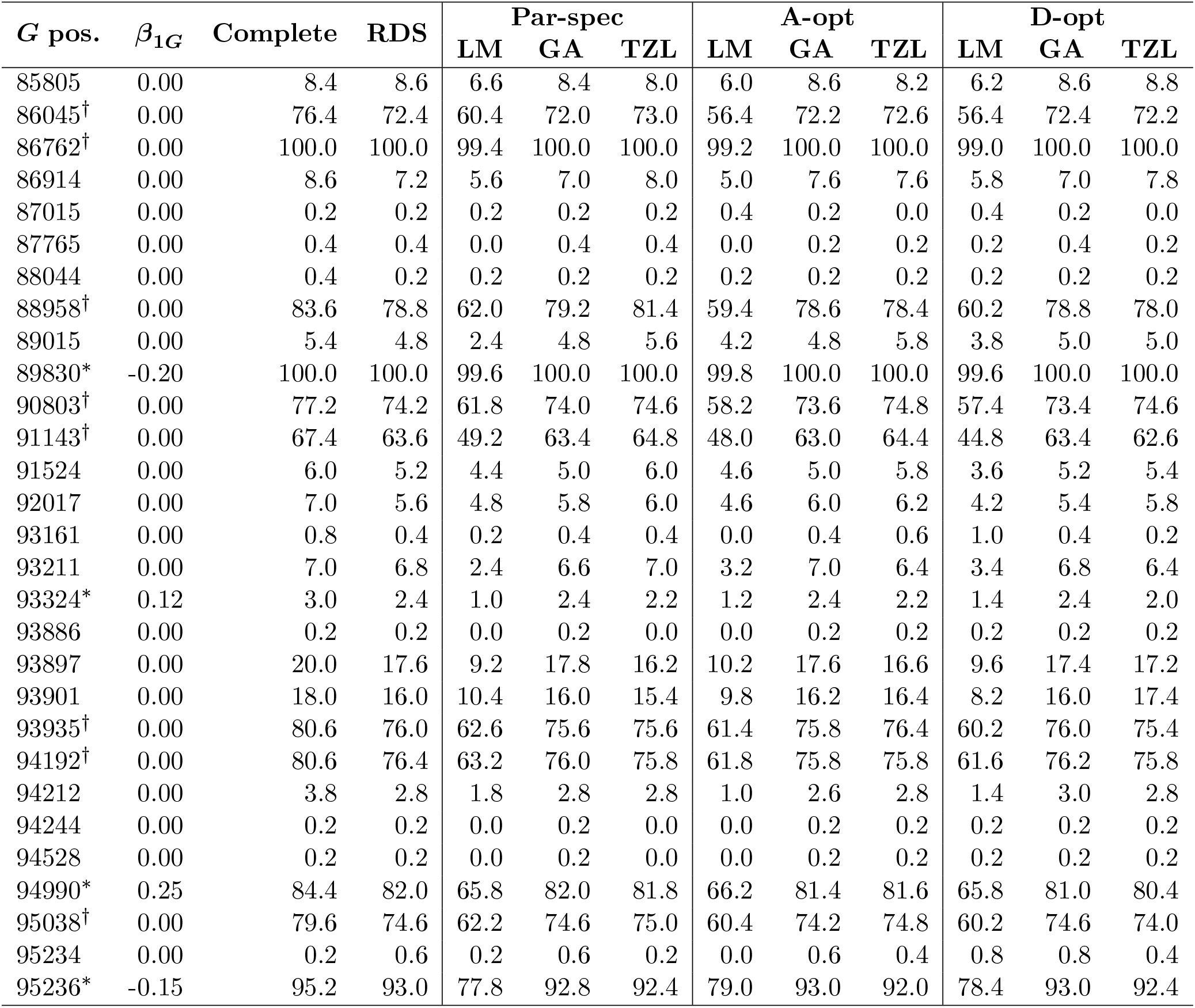
Empirical power rates at significance level *α* = 0.05*/*29 for causal variants only and *n* = 2500 across 500 replicates in realistic fine-mapping simulation single-variant analysis. Base pair positions (pos.) marked with ∗ denote causal variants whereas the ones marked with *†* denote hitchhikers. The remaining ones are non-causal. Positions are truncated to the last 5 digits.

| *G* pos. | $\beta_{1G}$ | Complete | RDS | Par-spec | | | A-opt | | | D-opt | | |
| --- | --- | --- | --- | --- | --- | --- | --- | --- | --- | --- | --- | --- |
|  |  |  |  | LM | GA | TZL | LM | GA | TZL | LM | GA | TZL |
| 85805 | 0.00 | 8.4 | 8.6 | 6.6 | 8.4 | 8.0 | 6.0 | 8.6 | 8.2 | 6.2 | 8.6 | 8.8 |
| 86045† | 0.00 | 76.4 | 72.4 | 60.4 | 72.0 | 73.0 | 56.4 | 72.2 | 72.6 | 56.4 | 72.4 | 72.2 |
| 86762† | 0.00 | 100.0 | 100.0 | 99.4 | 100.0 | 100.0 | 99.2 | 100.0 | 100.0 | 99.0 | 100.0 | 100.0 |
| 86914 | 0.00 | 8.6 | 7.2 | 5.6 | 7.0 | 8.0 | 5.0 | 7.6 | 7.6 | 5.8 | 7.0 | 7.8 |
| 87015 | 0.00 | 0.2 | 0.2 | 0.2 | 0.2 | 0.2 | 0.4 | 0.2 | 0.0 | 0.4 | 0.2 | 0.0 |
| 87765 | 0.00 | 0.4 | 0.4 | 0.0 | 0.4 | 0.4 | 0.0 | 0.2 | 0.2 | 0.2 | 0.4 | 0.2 |
| 88044 | 0.00 | 0.4 | 0.2 | 0.2 | 0.2 | 0.2 | 0.2 | 0.2 | 0.2 | 0.2 | 0.2 | 0.2 |
| 88958† | 0.00 | 83.6 | 78.8 | 62.0 | 79.2 | 81.4 | 59.4 | 78.6 | 78.4 | 60.2 | 78.8 | 78.0 |
| 89015 | 0.00 | 5.4 | 4.8 | 2.4 | 4.8 | 5.6 | 4.2 | 4.8 | 5.8 | 3.8 | 5.0 | 5.0 |
| 89830* | -0.20 | 100.0 | 100.0 | 99.6 | 100.0 | 100.0 | 99.8 | 100.0 | 100.0 | 99.6 | 100.0 | 100.0 |
| 90803† | 0.00 | 77.2 | 74.2 | 61.8 | 74.0 | 74.6 | 58.2 | 73.6 | 74.8 | 57.4 | 73.4 | 74.6 |
| 91143† | 0.00 | 67.4 | 63.6 | 49.2 | 63.4 | 64.8 | 48.0 | 63.0 | 64.4 | 44.8 | 63.4 | 62.6 |
| 91524 | 0.00 | 6.0 | 5.2 | 4.4 | 5.0 | 6.0 | 4.6 | 5.0 | 5.8 | 3.6 | 5.2 | 5.4 |
| 92017 | 0.00 | 7.0 | 5.6 | 4.8 | 5.8 | 6.0 | 4.6 | 6.0 | 6.2 | 4.2 | 5.4 | 5.8 |
| 93161 | 0.00 | 0.8 | 0.4 | 0.2 | 0.4 | 0.4 | 0.0 | 0.4 | 0.6 | 1.0 | 0.4 | 0.2 |
| 93211 | 0.00 | 7.0 | 6.8 | 2.4 | 6.6 | 7.0 | 3.2 | 7.0 | 6.4 | 3.4 | 6.8 | 6.4 |
| 93324* | 0.12 | 3.0 | 2.4 | 1.0 | 2.4 | 2.2 | 1.2 | 2.4 | 2.2 | 1.4 | 2.4 | 2.0 |
| 93886 | 0.00 | 0.2 | 0.2 | 0.0 | 0.2 | 0.0 | 0.0 | 0.2 | 0.2 | 0.2 | 0.2 | 0.2 |
| 93897 | 0.00 | 20.0 | 17.6 | 9.2 | 17.8 | 16.2 | 10.2 | 17.6 | 16.6 | 9.6 | 17.4 | 17.2 |
| 93901 | 0.00 | 18.0 | 16.0 | 10.4 | 16.0 | 15.4 | 9.8 | 16.2 | 16.4 | 8.2 | 16.0 | 17.4 |
| 93935† | 0.00 | 80.6 | 76.0 | 62.6 | 75.6 | 75.6 | 61.4 | 75.8 | 76.4 | 60.2 | 76.0 | 75.4 |
| 94192† | 0.00 | 80.6 | 76.4 | 63.2 | 76.0 | 75.8 | 61.8 | 75.8 | 75.8 | 61.6 | 76.2 | 75.8 |
| 94212 | 0.00 | 3.8 | 2.8 | 1.8 | 2.8 | 2.8 | 1.0 | 2.6 | 2.8 | 1.4 | 3.0 | 2.8 |
| 94244 | 0.00 | 0.2 | 0.2 | 0.0 | 0.2 | 0.0 | 0.0 | 0.2 | 0.2 | 0.2 | 0.2 | 0.2 |
| 94528 | 0.00 | 0.2 | 0.2 | 0.0 | 0.2 | 0.0 | 0.0 | 0.2 | 0.2 | 0.2 | 0.2 | 0.2 |
| 94990* | 0.25 | 84.4 | 82.0 | 65.8 | 82.0 | 81.8 | 66.2 | 81.4 | 81.6 | 65.8 | 81.0 | 80.4 |
| 95038† | 0.00 | 79.6 | 74.6 | 62.2 | 74.6 | 75.0 | 60.4 | 74.2 | 74.8 | 60.2 | 74.6 | 74.0 |
| 95234 | 0.00 | 0.2 | 0.6 | 0.2 | 0.6 | 0.2 | 0.0 | 0.6 | 0.4 | 0.8 | 0.8 | 0.4 |
| 95236* | -0.15 | 95.2 | 93.0 | 77.8 | 92.8 | 92.4 | 79.0 | 93.0 | 92.0 | 78.4 | 93.0 | 92.4 |

GA, RDS and TZL show similar results in terms of estimation and power across different values of *n* and optimality criteria (for GA and TZL) whereas LM exhibits considerably lower power (Tables S6-4). We observe similar distributions of the LR test *p*-value (in − log_10_ scale) across replicates for the studied designs with the exception of some outliers; LM shows the smallest (− log_10_) *p*-values compared to the other designs (Figures S6 and 4). Lastly, no optimality criterion shows consistently best estimation nor power, suggesting that no specific criteria substantially improves overall performance when the design values, *θ*_*des*_, are misspecified.

**Figure 4:**
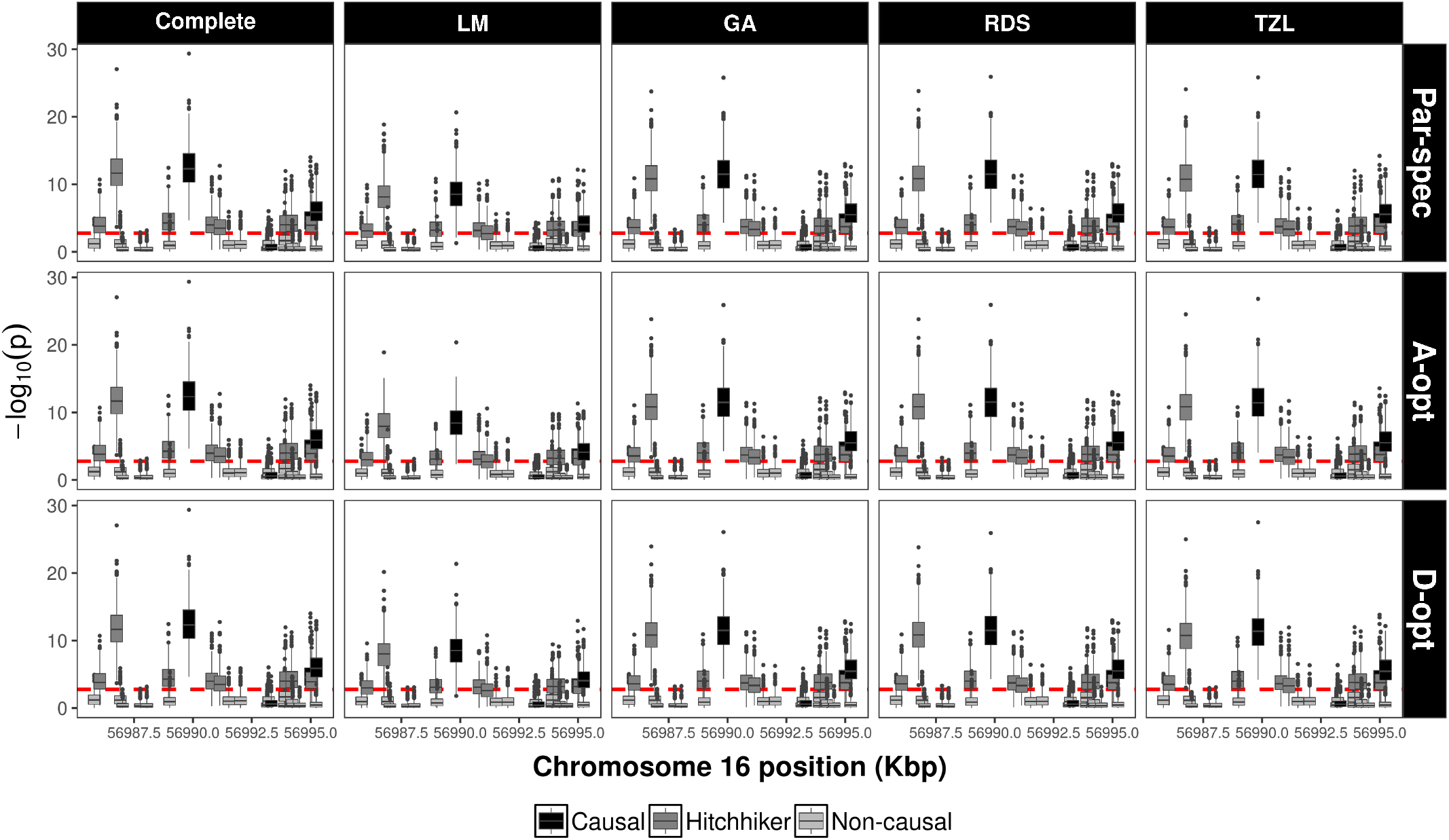
Boxplots of the (− log_10_) *p*-values across 500 replicates in the fine-mapping simulation single-variant analyses for a phase 2 sample size of *n* = 2500 across optimality criteria (for LM, GA, and TZL only): parameter-specific, A- and D-optimality in each row facet. Each column facet corresponds to the complete data analysis and studied designs respectively. The dashed line corresponds to a Bonferroni-corrected significance threshold of *α* = 0.05*/*29.

It is obvious that in most cases the mean of the estimate for causal sequence variants does not correspond with its true value being both over- and under- estimated (Tables S4-S5). In fact, this discrepancy occurs even for the complete data case, which is unsurprising considering the unaccounted variation resulting from the single-variant analysis.

The power to detect association (at *α* = 0.05*/*29) is above 80% in the complete data for 3 out of 4 causal variants: 89830, 94990, 95236. The power for the remaining causal variant (93324) is almost zero (Table S6). This decrease in power is likely due to the high LD between this variant and the GWAS SNP, *Z* (*r* = 0.94 and D′ = 0.95) which is already included in the regression, thus, diluting its signal. Moreover, there are additional non-causal variants that display power above 80% in the complete data analysis (Table S6). These so-called “hitchhiker variants” achieve significant association as a consequence of their LD with causal variants. The performance of the studied designs for the hitchhiker variants resembles the complete data analysis and its ranking is similar to the one shown with the causal variants. These results indicate that single-variant analysis does not distinguish well between causal and hitchhiker SNPs in complete nor two-phase analysis.

A common strategy to identify potential causal variants from hitchhikers consists of adjusting for the most significant variant (or variants) in the region and performing a new -conditional-scan (i.e. one variant at a time) fitting the following model: 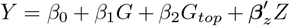, where *G* denotes a variant in the region, *G*_*top*_ is the most significant locus from the single-variant analysis, and *Z* is the GWAS SNP treated as a (3-level) categorical variable. This approach aims to discover independent signals in the region. Results for this conditional analysis can be found on Section S6, Online Supplementary Material.

## 5 Application in the Northern Finland Birth Cohort of 1966

We illustrate the methods outlined in Sections 2.4 and 4 using the Northern Finland Birth Cohort of 1966 (NFBC1966), which is a longitudinal, prospective birth cohort constituted by women and their offspring from the two northernmost provinces in Finland: Oulu and Lapland. Comprehensive phenotypic, lifestyle and demographic data were collected after birth via questionnaires and clinical evaluations on the offspring at years 1, 7, 14-16 and 31. The NFBC1966 aims to study genetic, biological, social or behavioural risk factors associated with the onset of different diseases as well as morbidity and mortality derived from adverse events such as pre-term birth and intrauterine growth retardation^[32;33]^. In particular, as part of an NHLBI-sponsored project designed to characterize the genetic determinants of metabolic and cardiovascular diseases, special attention was paid to a selected list of heritable quantitative traits related to cardiovascular diseases or type 2 diabetes. These traits are body mass index (BMI), high density lipoproteins (HDL), low density lipoproteins (LDL), triglycerides (TG), glucose (GLU), insulin (INS), C-reactive protein (CRP), systolic blood pressure (SBP) and diastolic blood pressure (DBP).

We focus on 5402 subjects for which genotype information was collected using the Illumina Infinium platform, which is comprised by 346,590 SNPs (after standard quality control). In addition to the genotype information, custom targeted sequencing (CTS) was collected for 4511 of them (83.5%) as part of a series of re-sequencing studies to deepen the understanding of genotypic variation on metabolic traits^[34]^. The CTS data contain the coding sequence and 5’ and 3’ untranslated regions of 78 genes, which were selected based on previous GWAS meta-analyses of cardiovascular diseases. Details of these regions can be found in Service *et al*. ^[34]^. The purpose of this illustration is to optimally select a subsample of subjects for targeted sequencing study (phase 2) and fine-mapping to locate potential causal variants using the methods outlined in the previous sections.

### 5.1 Phase 2 subsample selection

We first identify GWAS-SNPs by performing genome-wide associations on the available quantitative traits. Although genome-wide scans on these very same metabolic traits for the NFBC1966 have been previously carried out in Sabatti *et al*., our analyses differ in a couple of aspects: (1) the sample size we utilized is slightly larger because additional subjects were genotyped at a later time and (2) we perform multiple linear regression adjusting by the SexOCPG covariate described in Sabatti *et al*., which is a composed categorical variable determined by sex, oral contraceptive use and pregnancy status^[33]^.

We center attention on one trait, log-transformed TG (*Y*), as its GWAS has few peaks that identify only two genetic regions for further study: GCKR in chromosome 2 and LPL in chromosome 8 (Figure S7). We comment on the challenges of more complex GWAS scenarios in the discussion. We locate one SNPs in each of these regions that meet the usual genome-wide significance threshold (5 × 10^−8^): rs1260326 (chr2:27730940, 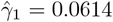, s.e. 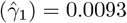, *p* = 5.67 × 10^−11^, MAF= 35.7%) and rs10096633 (chr8:19830921, 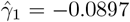, s.e. 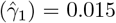, *p* = 3.24 × 10^−9^, MAF= 9.7%). Due to missing data on the TG values, the available subjects for the genome scan was 5300. Of these, the number of subjects with both GWAS and CTS data is *N* = 4493, which is the phase 1 sample size considered for phase 2 analyses. In addition to the GCKR and LPL regions used in the phase 2 selection, we consider another region for analysis: APOA5, to illustrate the correspondance of the two-phase design and analysis with the complete data approach pursued in Service *et al*. ^[34]^.

Using the two identified GWAS-SNPs, we select phase 2 subsamples under three of the previously described designs: GA, RDS and TZL. We drop LM as it showed the worst performance in simulations. The phase 2 sample size is specified to be approximately 25%, or 50% of the phase 1 sample size *(n* = 1123, 2246). To define *Z*, we use all allele combinations of the GWAS-SNPs rs1260326 and rs10096633, which results in a nine-category variable. Considering that no optimality criterion performed best in Section 4, we deem it appropriate to assume *β*_1_ is a scalar and use a parameter-specific criterion, which greatly simplifies the specification of the design quantities, particularly ***p***_*des*_. Phase 2 subsample selection is performed separately per each phase 2 sample size. In each case, a set of design quantities is defined as follows: First, 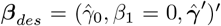, where 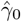 and 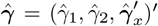 correspond to the MLEs from the following regression model based on phase 1 data: 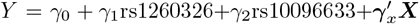, where ***X*** is a vector of additional covariates including SexOCPG and the first four genetic principal components (PC1-4). Second, for ***p***_*des*_ and given that MAF and LD values are unavailable *a priori*, we postulate the following ranges for these design quantities: *q*_*G*_ ∈ ⟨0.05 + 0.05*j*|*j* = 0, 1, …, 6⟩ and ***r*** = {0.0, *±*0.17, *±*0.33, *±*0.50}.

For visualization purposes, we categorize TG (*Y*_*st*_) into 3 groups corresponding to commonly used blood test ranges, i.e. normal (*<* 150 mg/dL), borderline high (150 − 199 mg/dL) and high (≥ 200 mg/dL). Notably, the groups in *Y*_*st*_ are asymmetrical with respect the middle *Y*_*st*_ stratum. On the other hand, the distribution of the nine-category variable determined by the two GWAS-SNPs (rs1260326 and rs10096633) has a small number of subjects for some categories due to the relatively low MAFs of SNPs rs10096633, which differs largely from the simulations (Figure S8). Consequently, the phase 2 subsample distributions tend to not select subjects from those categories. Notably, GA, RDS, and TZL show similar category distribution across phase 2 sample sizes (Figure S9). The proportion of subjects that are common among designs in the phase 2 subsamples is above 95% between pairs of GA, RDS, and TZL across phase 2 sample sizes (Figure S10).

### 5.2 Fine-mapping analysis

CTS data for genes GCKR, LPL and APOA5 were downloaded from the NCBI’s dbGAP repository according to their GRCh37.p13 location *±*5kbps. Since aligned reads were available, we performed variant calling using the GotCloud pipeline developed by the Center for Statistical Genetics at the University of Michigan^[35]^. Sequence data were analyzed in two ways. First, we used linear regression with complete data, i.e. subjects with both genotyping and CTS data (*N* = 4493), and second via the ML approach described above for each studied design: GA, RDS, and TZL (*n* = 1123, 2246). For the ML analysis, the nine-category variable defined by the GWAS SNPs was used as auxiliary variable *Z*. All analyses were adjusted by the GWAS-SNPs (rs1260326 and rs10096633), SexOCPG and PC1-PC4 as covariates.

Our main interest lies in gauging the performance of the two-phase designs with respect to the complete data analysis, evaluating both estimation and hypothesis testing. For estimation, we focus on three sequence variants reported in Table 2 of Service *et al*. ^[34]^: rs268, rs2266788, and rs3135506 (Table 5). For these variants, GA, RDS and TZL show similar results with improving performance as *n* increases. Comparisons of association estimates in Beta-Beta plots show similar spread estimates across designs (Figure S11). Similarly, region plots of association signals for complete data and ML analyses across the studied designs indicate that all designs tend to display results closer to those of the complete data case as *n* increases with no overwhelmingly better design (Figure 5). In addition to the region scans performed for analyzing common variants, we demonstrate that rare variants can be investigated under a two-phase design via burden tests in Section S7, Online Supplementary Material.

**Table 5:**
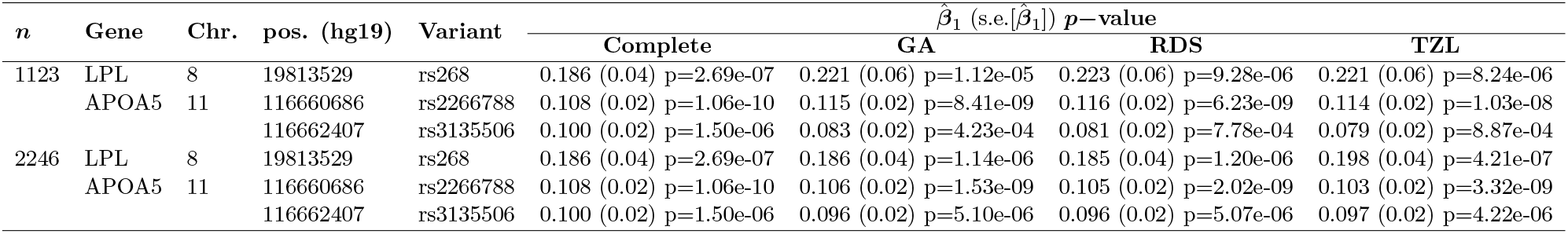
Estimation and testing results for analyzing (log-transformed) triglyceride levels across 3 sequence SNPs from the fine-mapping analysis in the NFBC66.

**Figure 5:**
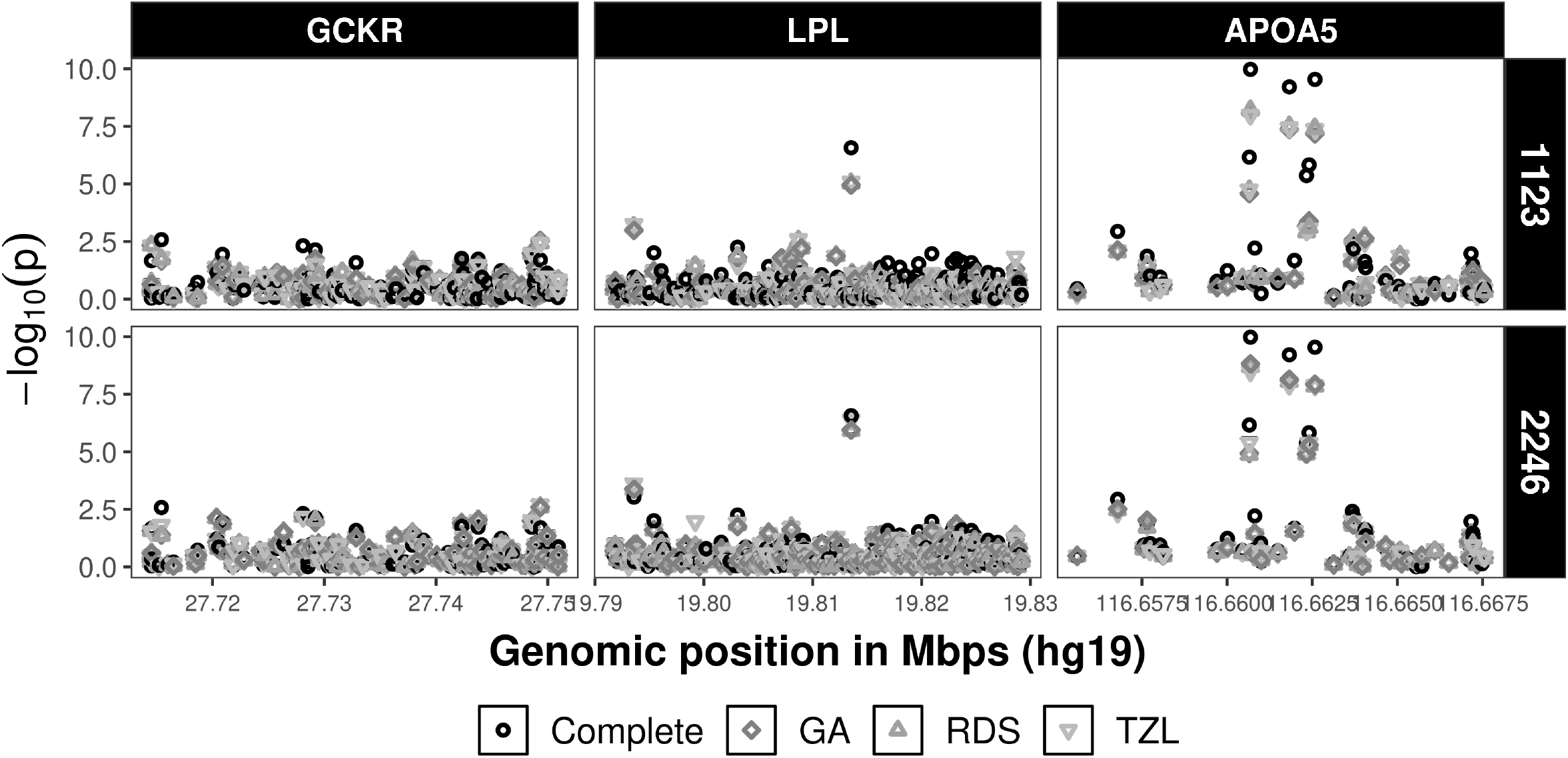
Region plots of the NFCBB66 CTS data for the ML analyses under the studied designs compared to the complete data analysis across different phase 2 sample sizes, *n* = 1123, 2246. Column facets show the region plot for each loci of interest. Row facets show different phase 2 sample sizes.

## 6 Discussion

In this report, we propose and evaluate state-of-the-art sample selection strategies for two-phase designs in the context of post-GWAS fine-mapping studies. We pay special attention in the comparison against a recently proposed optimal design, TZL. Our first set of simulations, considering a parameter-specific criterion, shows a clear advantage of TZL under the strong assumption of correctly specified design values (***p***_*des*_ or Var(*G*|*Z*)). On the other hand, TZL demonstrates biased estimation when the phase 2 sample size/non-missing fraction is small (*n/N* = 0.10, 0.25) and the effect sizes are farther from the null *β*_1_ ≥ 0.5, with improvements noticed when *n* increases. These results are aligned with those obtained under LM, which reinforces our initial belief that LM is a crude approximation to TZL. Moreover, LM is only competitive when *n* = 2500, suggesting that the sampling variability introduced by this method can only be overcome under larger non-missing fractions. In contrast, GA demonstrates unbiased estimation and competitive power (often larger than RDS) across all simulation settings. The appeal of GA lies in its generality as it can be extended to other settings beyond linear, logistic and Cox models while also avoiding uncertainty associated with sampling. Thus, GA provides an alternative approach to obtain efficient and robust two-phase designs across a wider range of settings, including large effect sizes.

Additionally, we investigate the use of different variances considered in the optimization: 𝕁^−1^, and 𝕍^−1^, which are, respectively, the inverse of the Fisher information matrix and the variance-covariance matrix of the most powerful test under the null hypothesis (Sections 3.2 and S3, Online Supplementary Material). The results support the use of 𝕍^−1^ for the parameter-specific criterion when the effect size is close to the null, as expected.

Correct design values specification is never attainable in practice. Hence, our second set of simulations evaluates a more realistic setting for which we propose a grid search approach to select a unique phase 2 design assuming a set of (mispecified) design values is available. In this scenario, GA, RDS and TZL have comparable performance with LM falling behind. GA is advantageous as it can be easily applied to general Λ(*·*) functions. Thus, apart from the parameter-specific criterion, we examine two additional criteria to select a phase 2 design: A- and D-optimality. Notably, based on our simulations we found no evidence in favour of any particular optimality criterion. However, the parameter-specific criterion may be preferred as designating its design values requires fewer assumptions. A- and D-optimality criteria were chosen because they have been amply explored in the literature, possess solid roots in experimental designs, and have natural connection with hypothesis testing. Nonetheless, other criteria may be better suited for optimizing power, for instance, maximizing the non-centrality parameter of the likelihood ratio or score test *χ*^2^ statistic may improve the power performance of the phase 2 designs.

An important observation from the simulations is that although the best performing designs have higher relative efficiency compared to less favorable designs (e.g. combined or simple random sampling) across all phase 2 sample sizes, this improvement does not automatically translate to a closer agreement with the complete data analysis. That is, power performance is contingent upon non-missing fraction and not necessarily phase 2 sample size itself. This finding is consistent throughout our investigations, where both parameter estimates and *p*−values achieve similar values as in the complete data analysis only when the phase 2 sample size is half of the phase 1 sample size (*n/N* = 0.5). Thus, careful evaluation of the statistical power of the phase 2 design needs to be considered in advance.

The competitive performance of GA notwithstanding, there are other considerations in implementing this algorithm. For instance, we provide in all simulations an ad-hoc approach to initialize the population of possible solutions to accelerate convergence. In general, giving a particular initialization is not necessary, however, a completely random initial population may need a larger number of generations (ℋ) to achieve good performance. In addition, given the stochastic nature of the search, there are few guarantees that the final solution has indeed reached a global as opposed to a local optimum. It is also worth mentioning that the tuning parameter settings for the proposed GA are intended as a guideline only and do not replace a more careful evaluation in specific problems. Walters suggests iterative calls, which consist of running the GA multiple times, so that the final population in each run serves as the initial population in subsequent runs^[29]^.

The budgetary constraint implicitly assumes that the cost of sequencing samples is the same for all study samples. This assumption may be relaxed as sequencing costs may vary due to location, tissue availability or number of samples. Thus, extensions to consider differential costs are yet to be considered; one example of such approach under tracing study designs can be found in Moon *et al*. ^[36]^. Another important issue that deserves further investigation in terms of budget constraints (or otherwise) is use of differential sequencing depths across samples.

Further investigation regarding selection of optimal phase 2 subsample under a set of loosely defined design values, *θ* = (***β′***, ***p′***) ′ ∈ Θ, is warranted. Indeed, beyond the proposed grid search approach, alternative means to select a phase 2 subsample across ranges of *θ* are possible, e.g. via min-max approaches^[37]^. However, this selection problem may also be addressed under a Bayesian framework for which a prior (joint) distribution for ***β*** and ***p*** needs to be specified^[3;38;39;40]^. The appeal of this approach is that it may better incorporate the uncertainty in the design values for selecting an phase 2 subsample although at the expense of computational complexity.

Beyond the feasibility and applicability of the proposed methodologies in practice, the illustration on the amply studied NFBC66 raises some additional questions on considerations posed in the design and analysis of two-phase post-GWAS fine-mapping studies. First, our rare-variant analysis shows no association in either of the studied regions. This result is not that surprising for a couple of reasons: the limited sample size in the NFBC66 and the low correlation between the GWAS SNPs and the computed genetic score. Additionally, the burden test assumes the same direction across all variants, which is a limitation in various settings. Thus, further investigation is required for variance component rare-variant tests, as the properties of these test statistics have not been thoroughly studied under the proposed missing by design scenario. Second, current practice in the field involves the imputation of GWAS data using high-quality reference panels such as the TOPMed Imputation Panel^[41]^. In principle, one can use the imputed variants to construct a suitable auxiliary variable, *Z*, to select a phase 2 subsample using the proposed framework. Alternative methods that accommodate differences between genotyped and imputed data for subjects not selected for phase 2 sequencing have been discussed^[42;43]^. Hence, comparisons between these methods and the approach undertaken in this article can be also evaluated. Lastly, in a similar vein, methodological extensions for situations when multiple loci are pinpointed by GWAS and/or multiple traits of equal interest are collected remain as topics of future work. A starting point in this direction may involve the calculation and application of polygenic risk scores to inform the phase 2 subsampling.

Another issue deserving further investigation is the influence of the phase 2 sample selection in the variant calling pipeline. For simplicity, in this application, variant calling was performed per each loci on all available CTS samples. However, even though genotype likelihoods are typically inferred by sample, population-specific filters such as MAF may change with the design. Thus, sensitivity analyses of these filters can be additionally explored.

We emphasize that although this report focuses on a normally distributed continuous trait, all the derivations apply in the context of generalized linear models within the exponential family. Furthermore, we are engaged in the development of an R package for this general case. Results of this research and the accompanying software aim to support investigators decision-making pertaining to study design, evaluation and analysis of two-phase studies. These tools can serve to make more efficient use of limited budgetary resources for data acquisition and analysis.

Two-phase study designs can be sought in other contexts. In particular, their use in a variety of ‘omics problems is broadly relevant as new and more costly technologies continue to arise. Beyond the case of fine-mapping where causal variants from GWAS-identified regions can be pinpointed at a fraction of the cost, this approach can be extended for phase 2 variables that are not categorical, for example, methylation, gene expression or other ‘omics measurements. Additionally, methods that introduce functional knowledge to further inform the inference (possibly through Bayesian methods) deserves further investigation.

## Data Availability

The data/analyses presented in the current publication are based on the use of study data downloaded from the dbGaP web site, under study id phs000276.v2.p1.

https://www.ncbi.nlm.nih.gov/projects/gap/cgi-bin/study.cgi?study_id=phs000276.v2.p1

## Acknowledgments

The NFBC1966 Study is conducted and supported by the National Heart, Lung, and Blood Institute (NHLBI) in collaboration with the Broad Institute, UCLA, University of Oulu, and the National Institute for Health and Welfare in Finland. This manuscript was not prepared in collaboration with investigators of the NFBC1966 Study and does not necessarily reflect the opinions or views of the NFBC1966 Study Investigators, Broad Institute, UCLA, University of Oulu, National Institute for Health and Welfare in Finland and the NHLBI.

Computations were performed on the Niagara supercomputer at the SciNet HPC Consortium and Galen, the HPC facility at the Lunenfed-Tanenbaum Research Institute (LTRI). SciNet is funded by: the Canada Foundation for Innovation under the auspices of Compute Canada; the Government of Ontario; Ontario Research Fund - Research Excellence; and the University of Toronto. The LTRI HPC facility is supported by the Canada Foundation for Innovation. OE-G would like to thank Prof. Olli Saarela and Prof. Richard J. Cook for constructive discussions and feedback. Authors thank the anonymous reviewers who helped enrich this work.

## Author contributions

OE-G wrote a first draft of this manuscript and subsequent revisions, performed simulations and analyzed the NFBC66 data. RVC and SBB provided statistical guidance and methodological support. All authors contributed in the design of the numerical experiments, overall analysis plan, as well as reviewing and approving the final version of the manuscript.

## Financial disclosure

This research is supported by funding from the Canadian Institutes of Health Research: CIHR Operating Grant MOP-84287 (RVC, SBB), CIHR Training Grant GET-101831 (OE-G); and the Ontario Institute for Cancer Research (OICR) through funding provided by the Government of Ontario (OE-G). OE-G has been fellow trainee of OICR Biostatistics Training Initiative and CIHR STAGE (Strategic Training for Advanced Genetic Epidemiology) - CIHR Training Grant in Genetic Epidemiology and Statistical Genetics.

## Data Availability

The data/analyses presented in the current publication are based on the use of study data downloaded from the dbGaP web site, under phs000276.v2.p1.

## Conflict of interest

The authors declare no potential conflict of interests.

## Supporting information

The supporting information file contains 7 additional sections, 24 tables, and 27 figures.

## A Derivation of the information matrix

This section derives the observed and expected information matrices, which allow us to compute the optimality criterion. The expected information matrix in particular is called repeatedly during the optimization performed in both LM and GA approaches.

### A.1 Log-likelihood

Considering the observed-data likelihood in (1), the log-likelihood takes the form

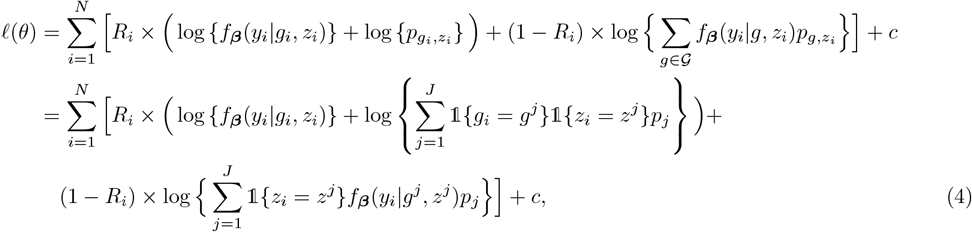

where ***p*** = (*p*_1_, …, *p*_*J*_) is the vector of probabilities corresponding to the *J* unique pairs (*g*^*j*^, *z*^*j*^) in 𝒢 × *Ƶ*.

### A.2 Score equations

Taking the first derivative of (4) with respect to ***β*** and ***p*** for a single observation, we have

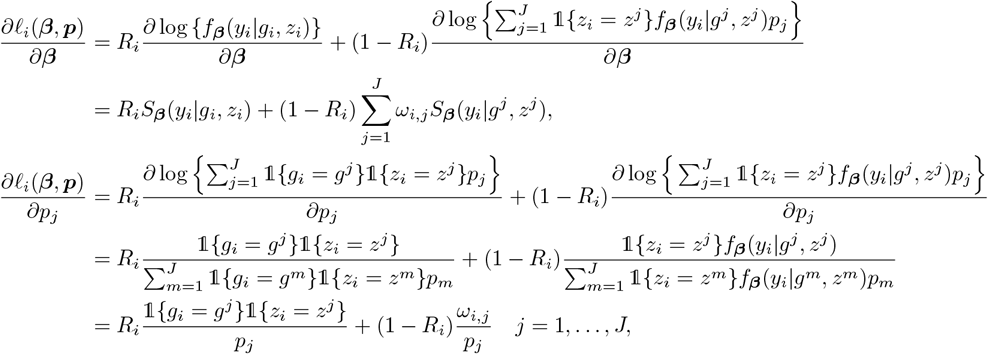

for the exponential family case, 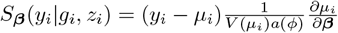, where *V* (*µ*_*i*_) is the variance function, *a*(*ϕ*) is the dispersion parameter and 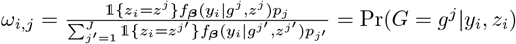 is the profile weight.

### A.3 Fisher Information Matrix

By definition, the Fisher information matrix (FIM) is given by 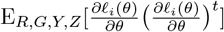.

Note that

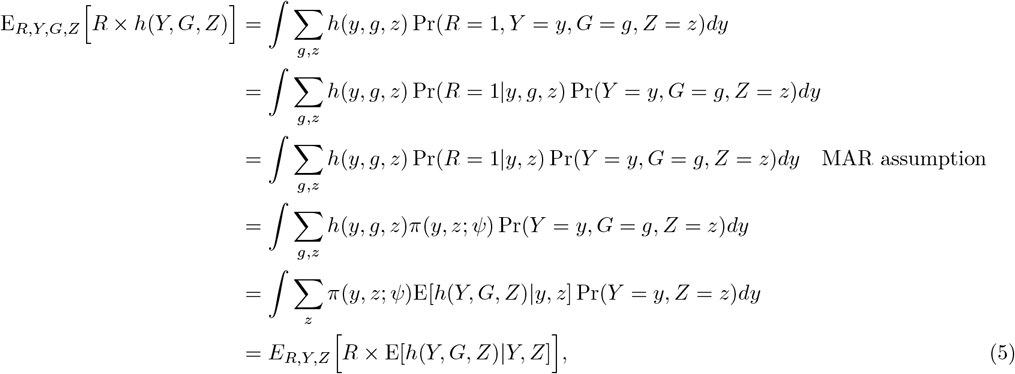

where the sums are taken with respect to all observed values of (*g, z*) and Pr(*Y* = *y, G* = *g, Z* = *z*) = Pr(*y, g, z*) = *f*_*β*_(*y*|*g, z*)*p*_*g,z*_.

Thus, considering that the score equations in subsection A.2 can be rewritten as

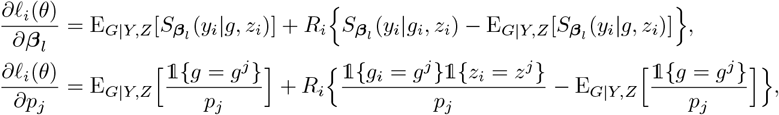

the cross-products are defined as follows

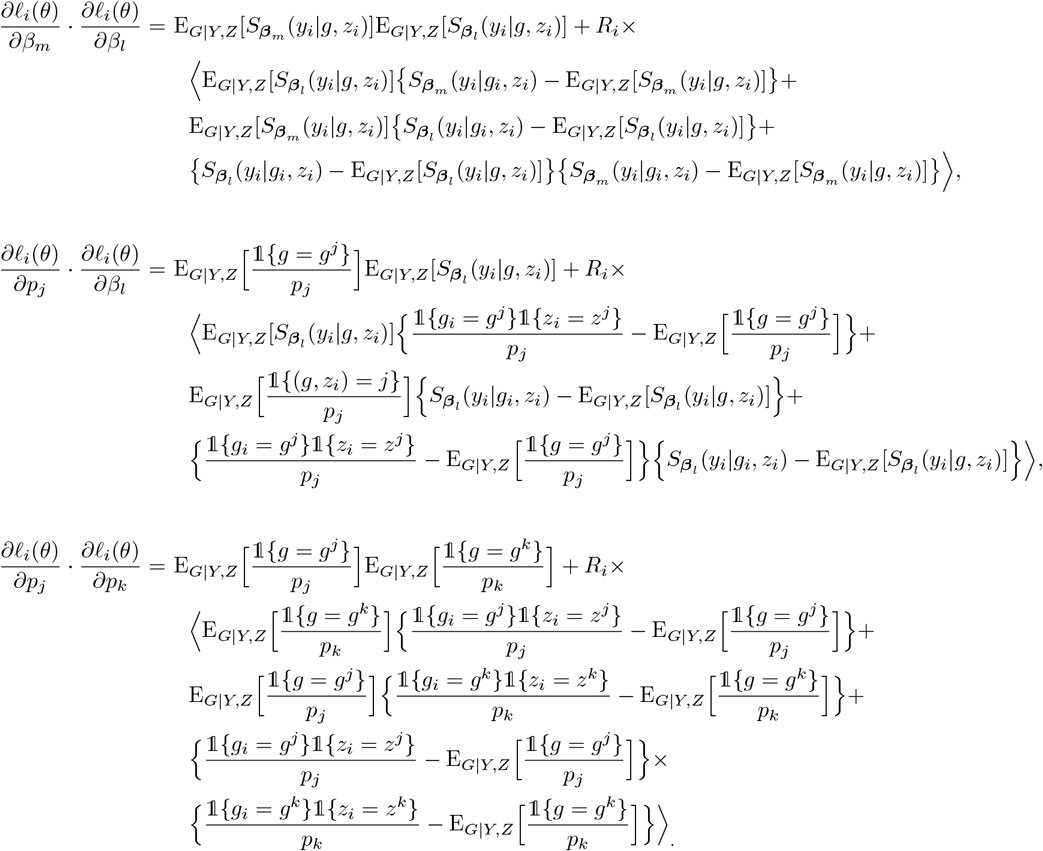

After taking expected values on both sides and by equation (5), we have

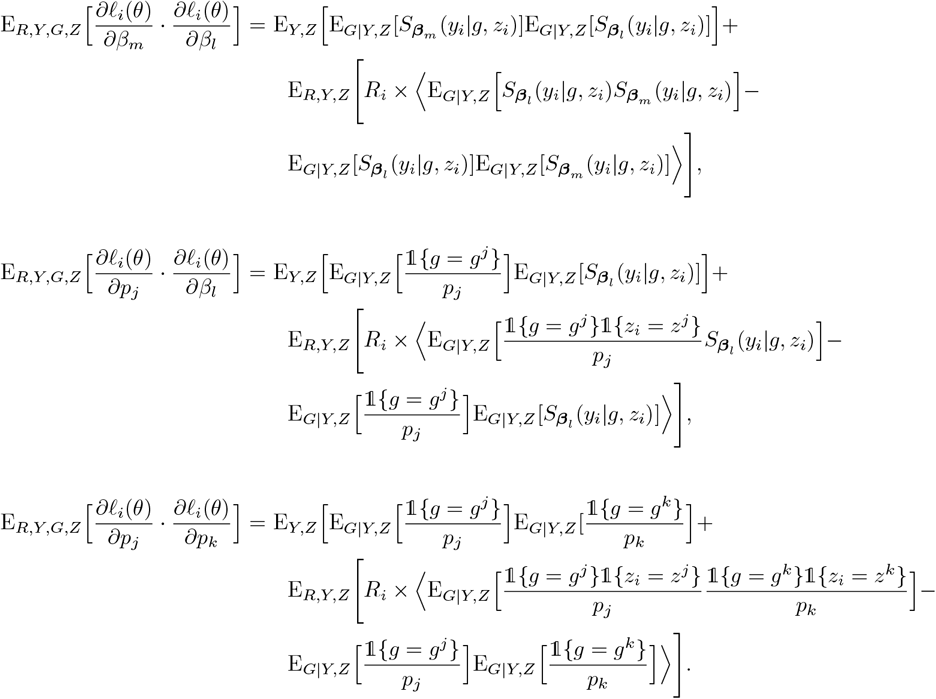

The expression above can be approximated empirically especially when *G* and *Z* are discrete. However, under the null hypothesis of interest, i.e. *β*_1_ = 0, *Y* and *G* are conditionally independent given *Z*. Thus, letting 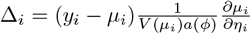, the expressions above simplify to:

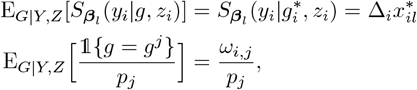

where 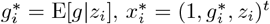, and ω_*i,j*_ = *Pr*(*G* = *g*^*j*^ | *z*_*i*_ = *z*^*j*^). Additionally,

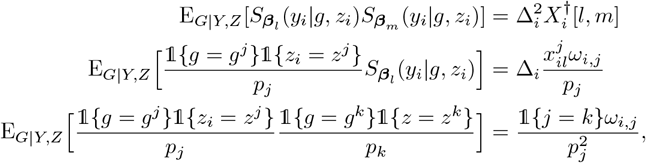

here, 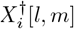,is the *l;m* entry of the outer product 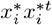 where 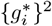 is replaced by E[*g*^2^ |*z*_*i*_] and 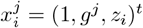.

Therefore, under the null hypothesis of interest, the Fisher information matrix can be expressed as

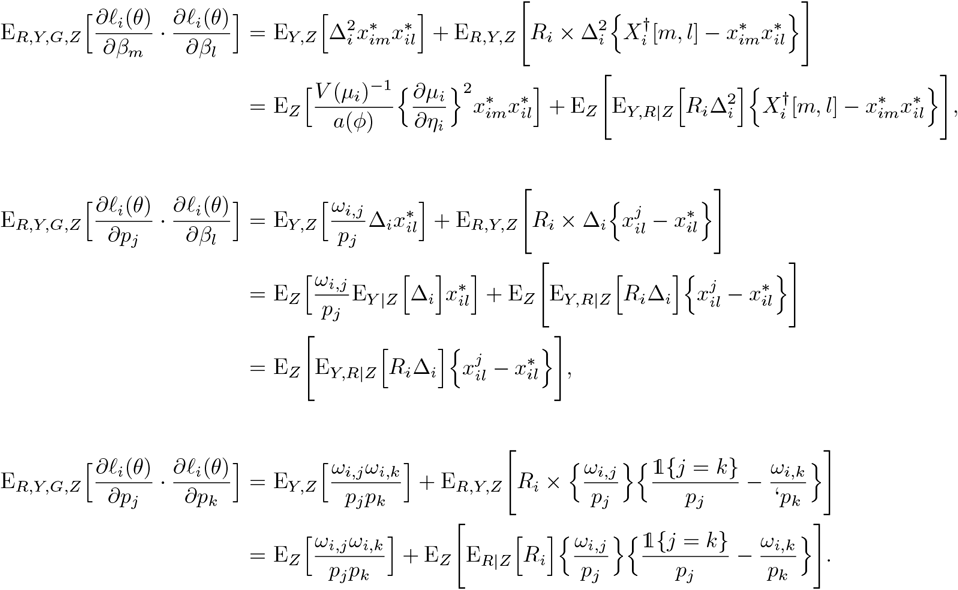

## Notes

### Competing Interest Statement

The authors have declared no competing interest.

