## Supplemental information for "Two-phase sample selection strategies for design and analysis in post-genome wide association fine-mapping studies"

This supplementary material contains 7 sections, including 24 tables and 27 figures. The content of each supplementary section is outlined below.

### S1 Complementary results from main text

#### S1.1 Tables

Table S1: Asymptotic/Empirical standard errors (ASE and ESE, respectively) comparing studied designs across effect sizes ($\beta_1$), and phase 2 sample sizes. Phase 1 sample size is $N = 5000$ whereas phase 2 sample size is $n = 540, 1250, 2500$.

| $n$ | $\beta_1$ | ASE | | | | | ESE | | | | |
|---|---|---|---|---|---|---|---|---|---|---|---|
| | | **Complete** | **LM** | **GA** | **RDS** | **TZL** | **Complete** | **LM** | **GA** | **RDS** | **TZL** |
| 540 | 0.0 | 0.025 | 0.066 | 0.038 | 0.037 | 0.043 | 0.025 | 0.068 | 0.039 | 0.038 | 0.044 |
| | 0.1 | 0.038 | 0.061 | 0.055 | 0.056 | 0.046 | 0.038 | 0.063 | 0.056 | 0.057 | 0.047 |
| | 0.3 | 0.038 | 0.056 | 0.054 | 0.055 | 0.044 | 0.038 | 0.058 | 0.056 | 0.057 | 0.046 |
| | 0.5 | 0.038 | 0.050 | 0.053 | 0.054 | 0.042 | 0.037 | 0.059 | 0.055 | 0.057 | 0.045 |
| | 0.7 | 0.038 | 0.045 | 0.053 | 0.053 | 0.041 | 0.037 | 0.057 | 0.059 | 0.060 | 0.044 |
| 1250 | 0.0 | 0.025 | 0.044 | 0.030 | 0.030 | 0.036 | 0.025 | 0.044 | 0.030 | 0.030 | 0.037 |
| | 0.1 | 0.038 | 0.046 | 0.044 | 0.045 | 0.040 | 0.038 | 0.045 | 0.045 | 0.045 | 0.039 |
| | 0.3 | 0.038 | 0.043 | 0.044 | 0.045 | 0.039 | 0.038 | 0.044 | 0.046 | 0.046 | 0.040 |
| | 0.5 | 0.038 | 0.040 | 0.045 | 0.045 | 0.037 | 0.037 | 0.045 | 0.045 | 0.046 | 0.039 |
| | 0.7 | 0.038 | 0.038 | 0.045 | 0.045 | 0.036 | 0.037 | 0.041 | 0.047 | 0.048 | 0.042 |
| 2500 | 0.0 | 0.025 | 0.034 | 0.034 | 0.026 | 0.028 | 0.025 | 0.034 | 0.034 | 0.026 | 0.028 |
| | 0.1 | 0.038 | 0.038 | 0.038 | 0.039 | 0.038 | 0.038 | 0.038 | 0.038 | 0.039 | 0.038 |
| | 0.3 | 0.038 | 0.037 | 0.038 | 0.039 | 0.038 | 0.038 | 0.037 | 0.039 | 0.040 | 0.038 |
| | 0.5 | 0.038 | 0.035 | 0.038 | 0.040 | 0.038 | 0.037 | 0.036 | 0.038 | 0.039 | 0.038 |
| | 0.7 | 0.038 | 0.033 | 0.038 | 0.040 | 0.038 | 0.037 | 0.036 | 0.040 | 0.040 | 0.038 |

Table S2: Relative asymptotic/empirical standard error (rASE/rESE, respectively) comparing studied designs across effect sizes ($\beta_1$) and phase 2 sample sizes. The ASE/ESE of each method (LM, GA, RDS, TZL) was calculated against the ASE/ESE of the complete data. Phase 1 sample size is $N = 5000$ whereas phase 2 sample size is $n = 540, 1250, 2500$.

| $n$ | $\beta_1$ | rASE | | | | rESE | | | |
|---|---|---|---|---|---|---|---|---|---|
| | | **LM** | **GA** | **RDS** | **TZL** | **LM** | **GA** | **RDS** | **TZL** |
| 540 | 0.0 | 2.635 | 1.524 | 1.479 | 1.720 | 2.682 | 1.543 | 1.499 | 1.728 |
| | 0.1 | 1.605 | 1.448 | 1.478 | 1.221 | 1.670 | 1.482 | 1.518 | 1.258 |
| | 0.3 | 1.469 | 1.435 | 1.462 | 1.173 | 1.540 | 1.485 | 1.506 | 1.223 |
| | 0.5 | 1.311 | 1.413 | 1.436 | 1.113 | 1.590 | 1.489 | 1.537 | 1.211 |
| | 0.7 | 1.179 | 1.390 | 1.406 | 1.072 | 1.548 | 1.579 | 1.628 | 1.179 |
| 1250 | 0.0 | 1.742 | 1.193 | 1.177 | 1.448 | 1.744 | 1.194 | 1.180 | 1.450 |
| | 0.1 | 1.210 | 1.172 | 1.178 | 1.049 | 1.192 | 1.201 | 1.212 | 1.038 |
| | 0.3 | 1.149 | 1.174 | 1.180 | 1.025 | 1.164 | 1.212 | 1.215 | 1.058 |
| | 0.5 | 1.067 | 1.180 | 1.185 | 0.991 | 1.212 | 1.223 | 1.234 | 1.047 |
| | 0.7 | 0.996 | 1.186 | 1.192 | 0.961 | 1.104 | 1.271 | 1.287 | 1.123 |
| 2500 | 0.0 | 1.348 | 1.336 | 1.038 | 1.125 | 1.343 | 1.337 | 1.040 | 1.126 |
| | 0.1 | 1.011 | 1.013 | 1.039 | 1.007 | 0.999 | 1.004 | 1.037 | 1.003 |
| | 0.3 | 0.976 | 1.010 | 1.043 | 1.008 | 0.991 | 1.022 | 1.063 | 1.014 |
| | 0.5 | 0.920 | 1.004 | 1.052 | 1.011 | 0.965 | 1.021 | 1.062 | 1.013 |
| | 0.7 | 0.874 | 1.007 | 1.063 | 1.015 | 0.960 | 1.070 | 1.064 | 1.027 |

Table S3: 95% coverage probabilities for $\beta_1$ across effect sizes and phase 2 sample sizes. Phase 1 sample size is $N = 5000$ whereas phase 2 sample size is $n = 540, 1250, 2500$.

| $\boldsymbol{n}$ | $\boldsymbol{\beta_1}$ | **LM** | **GA** | **RDS** | **TZL** |
|---|---|---|---|---|---|
| 540 | 0.0 | 94.1 | 94.6 | 94.6 | 95.1 |
| | 0.1 | 94.1 | 94.6 | 95.2 | 94.6 |
| | 0.3 | 93.2 | 94.2 | 94.9 | 94.1 |
| | 0.5 | 87.7 | 93.9 | 94.3 | 91.0 |
| | 0.7 | 65.3 | 92.5 | 91.8 | 71.7 |
| 1250 | 0.0 | 94.8 | 94.9 | 94.9 | 94.9 |
| | 0.1 | 95.0 | 94.6 | 94.2 | 95.0 |
| | 0.3 | 94.5 | 93.9 | 94.0 | 93.4 |
| | 0.5 | 89.5 | 94.9 | 94.3 | 93.8 |
| | 0.7 | 70.1 | 93.4 | 93.4 | 76.7 |
| 2500 | 0.0 | 95.0 | 94.9 | 95.0 | 94.7 |
| | 0.1 | 95.1 | 95.2 | 94.6 | 95.0 |
| | 0.3 | 93.4 | 94.0 | 94.1 | 94.1 |
| | 0.5 | 91.8 | 94.9 | 95.3 | 95.8 |
| | 0.7 | 69.8 | 93.7 | 95.4 | 95.2 |

Table S4: Estimates and asymptotic standard errors in fine-mapping simulation single-variant analysis for $n = 1250$ across optimality criteria (for LM, GA and TZL). The analysis summarizes 500 replicates. Base pair positions (pos.) marked with $*$ denote causal variants whereas the ones marked with † denote hitchhikers. The remaining ones are non-causal. Positions are truncated to the last 5 digits. LD measures (r, $r^2$, D′) are calculated with respect to the GWAS SNP, $Z$, rs247617 in 56990716 (hg19).

| $G$ pos. | $q_G$(%) | $r_{G,Z}$ | $r^2_{G,Z}$ | $D'_{G,Z}$ | $\beta_{1G}$ | Complete | RDS | Par-spec | | | A-opt | | | D-opt | | |
|---|---|---|---|---|---|---|---|---|---|---|---|---|---|---|---|---|
| | | | | | | | | LM | GA | TZL | LM | GA | TZL | LM | GA | TZL |
| 85805 | 15.7 | -0.23 | 0.05 | -0.83 | 0.00 | -0.052 (0.03) | -0.051 (0.03) | -0.056 (0.04) | -0.051 (0.03) | -0.051 (0.03) | -0.062 (0.05) | -0.052 (0.03) | -0.052 (0.03) | -0.061 (0.05) | -0.052 (0.03) | -0.054 (0.03) |
| 86045† | 23.2 | -0.21 | 0.04 | -0.59 | 0.00 | 0.094 (0.02) | 0.095 (0.03) | 0.101 (0.04) | 0.095 (0.03) | 0.095 (0.03) | 0.100 (0.04) | 0.095 (0.03) | 0.096 (0.03) | 0.099 (0.04) | 0.095 (0.03) | 0.098 (0.03) |
| 86762† | 47.9 | -0.40 | 0.16 | -0.65 | 0.00 | -0.153 (0.02) | -0.154 (0.03) | -0.152 (0.03) | -0.154 (0.03) | -0.154 (0.03) | -0.149 (0.04) | -0.154 (0.03) | -0.154 (0.03) | -0.150 (0.04) | -0.154 (0.03) | -0.153 (0.03) |
| 86914 | 16.4 | -0.25 | 0.06 | -0.87 | 0.00 | -0.052 (0.03) | -0.051 (0.03) | -0.056 (0.04) | -0.051 (0.03) | -0.051 (0.03) | -0.059 (0.05) | -0.051 (0.03) | -0.052 (0.03) | -0.059 (0.05) | -0.051 (0.03) | -0.053 (0.03) |
| 87015 | 29.4 | 0.98 | 0.96 | 0.98 | 0.00 | -0.032 (0.11) | -0.035 (0.14) | -0.064 (0.23) | -0.035 (0.14) | -0.036 (0.14) | -0.059 (0.15) | -0.037 (0.14) | -0.042 (0.14) | -0.061 (0.15) | -0.036 (0.14) | -0.048 (0.15) |
| 87765 | 28.7 | 0.96 | 0.93 | 0.98 | 0.00 | 0.023 (0.09) | 0.024 (0.10) | -0.005 (0.24) | 0.024 (0.10) | 0.025 (0.11) | -0.008 (0.13) | 0.022 (0.10) | 0.017 (0.11) | -0.006 (0.13) | 0.022 (0.10) | 0.012 (0.11) |
| 88044 | 29.9 | 0.95 | 0.90 | 0.96 | 0.00 | 0.029 (0.07) | 0.028 (0.08) | 0.019 (0.13) | 0.027 (0.08) | 0.026 (0.09) | 0.033 (0.11) | 0.028 (0.08) | 0.029 (0.08) | 0.036 (0.11) | 0.028 (0.08) | 0.027 (0.09) |
| 88958† | 43.8 | 0.26 | 0.07 | 0.36 | 0.00 | -0.087 (0.02) | -0.087 (0.02) | -0.096 (0.04) | -0.087 (0.02) | -0.088 (0.03) | -0.090 (0.04) | -0.087 (0.02) | -0.087 (0.03) | -0.090 (0.04) | -0.086 (0.02) | -0.089 (0.03) |
| 89015 | 2.3 | -0.06 | 0.00 | -0.61 | 0.00 | 0.106 (0.07) | 0.107 (0.08) | 0.118 (0.10) | 0.106 (0.08) | 0.107 (0.08) | 0.122 (0.12) | 0.106 (0.08) | 0.108 (0.08) | 0.116 (0.12) | 0.106 (0.08) | 0.111 (0.08) |
| 89830* | 47.8 | -0.41 | 0.17 | -0.67 | -0.20 | -0.159 (0.02) | -0.159 (0.03) | -0.156 (0.03) | -0.159 (0.03) | -0.159 (0.03) | -0.155 (0.04) | -0.159 (0.03) | -0.160 (0.03) | -0.157 (0.04) | -0.159 (0.03) | -0.158 (0.03) |
| 90803† | 22.6 | -0.19 | 0.04 | -0.55 | 0.00 | 0.095 (0.02) | 0.096 (0.03) | 0.105 (0.04) | 0.096 (0.03) | 0.097 (0.03) | 0.103 (0.04) | 0.096 (0.03) | 0.098 (0.03) | 0.102 (0.04) | 0.096 (0.03) | 0.100 (0.03) |
| 91143† | 11.0 | -0.15 | 0.02 | -0.67 | 0.00 | 0.118 (0.03) | 0.118 (0.04) | 0.120 (0.05) | 0.118 (0.04) | 0.119 (0.04) | 0.118 (0.05) | 0.118 (0.04) | 0.118 (0.04) | 0.114 (0.05) | 0.119 (0.04) | 0.118 (0.04) |
| 91524 | 16.3 | -0.26 | 0.07 | -0.91 | 0.00 | -0.046 (0.03) | -0.045 (0.03) | -0.051 (0.04) | -0.045 (0.03) | -0.046 (0.03) | -0.054 (0.05) | -0.046 (0.03) | -0.047 (0.03) | -0.055 (0.05) | -0.045 (0.03) | -0.048 (0.03) |
| 92017 | 16.7 | -0.26 | 0.07 | -0.90 | 0.00 | -0.047 (0.03) | -0.046 (0.03) | -0.051 (0.04) | -0.046 (0.03) | -0.046 (0.03) | -0.053 (0.05) | -0.046 (0.03) | -0.047 (0.03) | -0.055 (0.05) | -0.046 (0.03) | -0.049 (0.03) |
| 93161 | 28.8 | 0.95 | 0.91 | 0.96 | 0.00 | 0.047 (0.07) | 0.050 (0.09) | 0.038 (0.18) | 0.051 (0.09) | 0.051 (0.09) | 0.026 (0.12) | 0.050 (0.09) | 0.048 (0.09) | 0.028 (0.12) | 0.050 (0.09) | 0.044 (0.10) |
| 93211 | 18.5 | -0.19 | 0.03 | -0.61 | 0.00 | -0.045 (0.03) | -0.044 (0.03) | -0.038 (0.04) | -0.044 (0.03) | -0.043 (0.03) | -0.038 (0.05) | -0.045 (0.03) | -0.044 (0.03) | -0.043 (0.05) | -0.045 (0.03) | -0.043 (0.03) |
| 93324* | 29.4 | 0.94 | 0.89 | 0.95 | 0.12 | 0.074 (0.07) | 0.076 (0.08) | 0.070 (0.13) | 0.076 (0.08) | 0.076 (0.08) | 0.056 (0.11) | 0.075 (0.08) | 0.075 (0.08) | 0.056 (0.11) | 0.076 (0.08) | 0.071 (0.08) |
| 93886 | 29.2 | 0.95 | 0.90 | 0.95 | 0.00 | 0.040 (0.07) | 0.041 (0.08) | 0.032 (0.15) | 0.041 (0.08) | 0.041 (0.09) | 0.027 (0.12) | 0.041 (0.08) | 0.041 (0.09) | 0.028 (0.12) | 0.041 (0.08) | 0.039 (0.09) |
| 93897 | 26.0 | -0.30 | 0.09 | -0.78 | 0.00 | -0.056 (0.02) | -0.056 (0.03) | -0.054 (0.04) | -0.056 (0.03) | -0.055 (0.03) | -0.059 (0.04) | -0.057 (0.03) | -0.057 (0.03) | -0.060 (0.04) | -0.056 (0.03) | -0.056 (0.03) |
| 93901 | 18.1 | -0.19 | 0.04 | -0.63 | 0.00 | -0.059 (0.03) | -0.059 (0.03) | -0.053 (0.04) | -0.059 (0.03) | -0.057 (0.03) | -0.055 (0.05) | -0.059 (0.03) | -0.058 (0.03) | -0.059 (0.05) | -0.059 (0.03) | -0.058 (0.03) |
| 93935† | 22.9 | -0.17 | 0.03 | -0.49 | 0.00 | 0.097 (0.02) | 0.098 (0.03) | 0.107 (0.04) | 0.098 (0.03) | 0.098 (0.03) | 0.104 (0.04) | 0.098 (0.03) | 0.099 (0.03) | 0.104 (0.04) | 0.098 (0.03) | 0.101 (0.03) |
| 94192† | 22.9 | -0.17 | 0.03 | -0.49 | 0.00 | 0.097 (0.02) | 0.098 (0.03) | 0.107 (0.04) | 0.098 (0.03) | 0.098 (0.03) | 0.104 (0.04) | 0.098 (0.03) | 0.099 (0.03) | 0.103 (0.04) | 0.098 (0.03) | 0.101 (0.03) |
| 94212 | 31.4 | -0.25 | 0.06 | -0.57 | 0.00 | -0.028 (0.02) | -0.029 (0.03) | -0.023 (0.04) | -0.029 (0.03) | -0.028 (0.03) | -0.024 (0.04) | -0.029 (0.03) | -0.028 (0.03) | -0.026 (0.04) | -0.029 (0.03) | -0.027 (0.03) |
| 94244 | 29.2 | 0.95 | 0.90 | 0.95 | 0.00 | 0.040 (0.07) | 0.041 (0.08) | 0.032 (0.15) | 0.041 (0.08) | 0.041 (0.09) | 0.027 (0.12) | 0.041 (0.08) | 0.041 (0.09) | 0.028 (0.12) | 0.041 (0.08) | 0.039 (0.09) |
| 94528 | 29.2 | 0.95 | 0.90 | 0.95 | 0.00 | 0.040 (0.07) | 0.041 (0.08) | 0.032 (0.15) | 0.041 (0.08) | 0.041 (0.09) | 0.027 (0.12) | 0.041 (0.08) | 0.041 (0.09) | 0.028 (0.12) | 0.041 (0.08) | 0.039 (0.09) |
| 94990* | 12.6 | -0.15 | 0.02 | -0.60 | 0.25 | 0.129 (0.03) | 0.129 (0.04) | 0.130 (0.05) | 0.129 (0.04) | 0.130 (0.04) | 0.129 (0.05) | 0.129 (0.04) | 0.129 (0.04) | 0.125 (0.05) | 0.130 (0.04) | 0.130 (0.04) |
| 95038† | 22.7 | -0.18 | 0.03 | -0.51 | 0.00 | 0.096 (0.02) | 0.097 (0.03) | 0.107 (0.04) | 0.097 (0.03) | 0.098 (0.03) | 0.104 (0.04) | 0.097 (0.03) | 0.099 (0.03) | 0.103 (0.04) | 0.097 (0.03) | 0.101 (0.03) |
| 95234 | 6.5 | -0.17 | 0.03 | -1.00 | 0.00 | -0.034 (0.04) | -0.033 (0.05) | -0.040 (0.06) | -0.033 (0.05) | -0.034 (0.05) | -0.042 (0.07) | -0.033 (0.05) | -0.035 (0.05) | -0.042 (0.07) | -0.033 (0.05) | -0.035 (0.05) |
| 95236* | 48.6 | 0.54 | 0.29 | 0.82 | -0.15 | -0.117 (0.02) | -0.119 (0.03) | -0.108 (0.04) | -0.119 (0.03) | -0.117 (0.03) | -0.117 (0.04) | -0.119 (0.03) | -0.118 (0.03) | -0.114 (0.04) | -0.119 (0.03) | -0.117 (0.03) |

Table S5: Estimates and asymptotic standard errors in fine-mapping simulation single-variant analysis for $n = 2500$ across optimality criteria (for LM, GA and TZL). The analysis summarizes 500 replicates. Base pair positions (pos.) marked with $*$ denote causal variants whereas the ones marked with † denote hitchhikers. The remaining ones are non-causal. Positions are truncated to the last 5 digits. LD measures (r, $r^2$, $D'$) are calculated with respect to the GWAS SNP, $Z$, rs247617 in 56990716 (hg19).

| $G$ pos. | $q_G$(%) | $r_{G,Z}$ | $r^2_{G,Z}$ | $D'_{G,Z}$ | $\beta_{1G}$ | Complete | RDS | Par-spec | | | A-opt | | | D-opt | | |
|---|---|---|---|---|---|---|---|---|---|---|---|---|---|---|---|---|
| | | | | | | | | LM | GA | TZL | LM | GA | TZL | LM | GA | TZL |
| 85805 | 15.7 | -0.23 | 0.05 | -0.83 | 0.00 | -0.052 (0.03) | -0.052 (0.03) | -0.054 (0.03) | -0.052 (0.03) | -0.052 (0.03) | -0.055 (0.03) | -0.052 (0.03) | -0.052 (0.03) | -0.055 (0.03) | -0.052 (0.03) | -0.052 (0.03) |
| 86045† | 23.2 | -0.21 | 0.04 | -0.59 | 0.00 | 0.094 (0.02) | 0.095 (0.03) | 0.098 (0.03) | 0.095 (0.03) | 0.095 (0.03) | 0.099 (0.03) | 0.095 (0.03) | 0.095 (0.03) | 0.098 (0.03) | 0.095 (0.03) | 0.095 (0.03) |
| 86762† | 47.9 | -0.40 | 0.16 | -0.65 | 0.00 | -0.153 (0.02) | -0.153 (0.02) | -0.153 (0.03) | -0.153 (0.02) | -0.153 (0.02) | -0.151 (0.03) | -0.153 (0.02) | -0.153 (0.02) | -0.151 (0.03) | -0.153 (0.02) | -0.153 (0.02) |
| 86914 | 16.4 | -0.25 | 0.06 | -0.87 | 0.00 | -0.052 (0.03) | -0.052 (0.03) | -0.054 (0.03) | -0.051 (0.03) | -0.052 (0.03) | -0.054 (0.03) | -0.052 (0.03) | -0.052 (0.03) | -0.055 (0.03) | -0.051 (0.03) | -0.051 (0.03) |
| 87015 | 29.4 | 0.98 | 0.96 | 0.98 | 0.00 | -0.032 (0.11) | -0.034 (0.12) | -0.041 (0.15) | -0.034 (0.12) | -0.032 (0.12) | -0.060 (0.15) | -0.033 (0.12) | -0.035 (0.12) | -0.051 (0.15) | -0.033 (0.12) | -0.034 (0.12) |
| 87765 | 28.7 | 0.96 | 0.93 | 0.98 | 0.00 | 0.023 (0.09) | 0.023 (0.09) | 0.013 (0.13) | 0.023 (0.09) | 0.023 (0.09) | 0.002 (0.12) | 0.022 (0.09) | 0.021 (0.09) | 0.008 (0.11) | 0.023 (0.09) | 0.022 (0.09) |
| 88044 | 29.9 | 0.95 | 0.90 | 0.96 | 0.00 | 0.029 (0.07) | 0.029 (0.07) | 0.017 (0.09) | 0.029 (0.07) | 0.028 (0.07) | 0.028 (0.09) | 0.029 (0.07) | 0.029 (0.07) | 0.029 (0.09) | 0.029 (0.07) | 0.030 (0.07) |
| 88958† | 43.8 | 0.26 | 0.07 | 0.36 | 0.00 | -0.087 (0.02) | -0.087 (0.02) | -0.092 (0.03) | -0.087 (0.02) | -0.087 (0.02) | -0.090 (0.03) | -0.087 (0.02) | -0.087 (0.02) | -0.090 (0.03) | -0.087 (0.02) | -0.087 (0.02) |
| 89015 | 2.3 | -0.06 | 0.00 | -0.61 | 0.00 | 0.106 (0.07) | 0.106 (0.07) | 0.110 (0.08) | 0.106 (0.07) | 0.107 (0.07) | 0.113 (0.08) | 0.106 (0.07) | 0.108 (0.07) | 0.111 (0.08) | 0.106 (0.07) | 0.108 (0.07) |
| 89830* | 47.8 | -0.41 | 0.17 | -0.67 | -0.20 | -0.159 (0.02) | -0.158 (0.02) | -0.157 (0.03) | -0.158 (0.02) | -0.159 (0.02) | -0.157 (0.03) | -0.158 (0.02) | -0.158 (0.02) | -0.157 (0.03) | -0.158 (0.02) | -0.158 (0.02) |
| 90803† | 22.6 | -0.19 | 0.04 | -0.55 | 0.00 | 0.095 (0.02) | 0.096 (0.03) | 0.100 (0.03) | 0.096 (0.03) | 0.096 (0.03) | 0.101 (0.03) | 0.096 (0.03) | 0.096 (0.03) | 0.100 (0.03) | 0.096 (0.03) | 0.096 (0.03) |
| 91143† | 11.0 | -0.15 | 0.02 | -0.67 | 0.00 | 0.118 (0.03) | 0.119 (0.03) | 0.117 (0.04) | 0.119 (0.03) | 0.119 (0.03) | 0.119 (0.04) | 0.118 (0.03) | 0.119 (0.03) | 0.118 (0.04) | 0.119 (0.03) | 0.119 (0.03) |
| 91524 | 16.3 | -0.26 | 0.07 | -0.91 | 0.00 | -0.046 (0.03) | -0.046 (0.03) | -0.048 (0.03) | -0.046 (0.03) | -0.046 (0.03) | -0.050 (0.03) | -0.046 (0.03) | -0.046 (0.03) | -0.050 (0.03) | -0.046 (0.03) | -0.046 (0.03) |
| 92017 | 16.7 | -0.26 | 0.07 | -0.90 | 0.00 | -0.047 (0.03) | -0.047 (0.03) | -0.049 (0.03) | -0.047 (0.03) | -0.047 (0.03) | -0.049 (0.03) | -0.047 (0.03) | -0.047 (0.03) | -0.050 (0.03) | -0.047 (0.03) | -0.047 (0.03) |
| 93161 | 28.8 | 0.95 | 0.91 | 0.96 | 0.00 | 0.047 (0.07) | 0.047 (0.08) | 0.043 (0.11) | 0.047 (0.08) | 0.048 (0.08) | 0.033 (0.10) | 0.047 (0.08) | 0.047 (0.08) | 0.039 (0.10) | 0.047 (0.08) | 0.047 (0.08) |
| 93211 | 18.5 | -0.19 | 0.03 | -0.61 | 0.00 | -0.045 (0.03) | -0.045 (0.03) | -0.041 (0.03) | -0.045 (0.03) | -0.044 (0.03) | -0.040 (0.03) | -0.045 (0.03) | -0.044 (0.03) | -0.042 (0.03) | -0.045 (0.03) | -0.044 (0.03) |
| 93324* | 29.4 | 0.94 | 0.89 | 0.95 | 0.12 | 0.074 (0.07) | 0.074 (0.07) | 0.071 (0.09) | 0.074 (0.07) | 0.074 (0.07) | 0.063 (0.09) | 0.074 (0.07) | 0.073 (0.07) | 0.068 (0.09) | 0.074 (0.07) | 0.074 (0.07) |
| 93886 | 29.2 | 0.95 | 0.90 | 0.95 | 0.00 | 0.040 (0.07) | 0.040 (0.07) | 0.033 (0.10) | 0.040 (0.07) | 0.040 (0.07) | 0.031 (0.10) | 0.040 (0.07) | 0.041 (0.07) | 0.036 (0.09) | 0.040 (0.07) | 0.041 (0.08) |
| 93897 | 26.0 | -0.30 | 0.09 | -0.78 | 0.00 | -0.056 (0.02) | -0.056 (0.02) | -0.054 (0.03) | -0.056 (0.02) | -0.056 (0.02) | -0.056 (0.03) | -0.056 (0.02) | -0.056 (0.02) | -0.057 (0.03) | -0.056 (0.02) | -0.056 (0.03) |
| 93901 | 18.1 | -0.19 | 0.04 | -0.63 | 0.00 | -0.059 (0.03) | -0.059 (0.03) | -0.055 (0.03) | -0.059 (0.03) | -0.058 (0.03) | -0.056 (0.03) | -0.059 (0.03) | -0.058 (0.03) | -0.058 (0.03) | -0.059 (0.03) | -0.058 (0.03) |
| 93935† | 22.9 | -0.17 | 0.03 | -0.49 | 0.00 | 0.097 (0.02) | 0.097 (0.03) | 0.102 (0.03) | 0.097 (0.03) | 0.098 (0.03) | 0.103 (0.03) | 0.097 (0.03) | 0.098 (0.03) | 0.102 (0.03) | 0.097 (0.03) | 0.098 (0.03) |
| 94192† | 22.9 | -0.17 | 0.03 | -0.49 | 0.00 | 0.097 (0.02) | 0.097 (0.03) | 0.102 (0.03) | 0.097 (0.03) | 0.098 (0.03) | 0.103 (0.03) | 0.097 (0.03) | 0.098 (0.03) | 0.102 (0.03) | 0.097 (0.03) | 0.098 (0.03) |
| 94212 | 31.4 | -0.25 | 0.06 | -0.57 | 0.00 | -0.028 (0.02) | -0.028 (0.02) | -0.027 (0.03) | -0.028 (0.02) | -0.028 (0.02) | -0.025 (0.03) | -0.028 (0.02) | -0.028 (0.02) | -0.027 (0.03) | -0.028 (0.02) | -0.028 (0.02) |
| 94244 | 29.2 | 0.95 | 0.90 | 0.95 | 0.00 | 0.040 (0.07) | 0.040 (0.07) | 0.033 (0.10) | 0.040 (0.07) | 0.040 (0.07) | 0.031 (0.10) | 0.040 (0.07) | 0.041 (0.07) | 0.036 (0.09) | 0.040 (0.07) | 0.041 (0.08) |
| 94528 | 29.2 | 0.95 | 0.90 | 0.95 | 0.00 | 0.040 (0.07) | 0.040 (0.07) | 0.033 (0.10) | 0.040 (0.07) | 0.040 (0.07) | 0.031 (0.10) | 0.040 (0.07) | 0.041 (0.07) | 0.036 (0.09) | 0.040 (0.07) | 0.041 (0.08) |
| 94990* | 12.6 | -0.15 | 0.02 | -0.60 | 0.25 | 0.129 (0.03) | 0.130 (0.03) | 0.128 (0.04) | 0.130 (0.03) | 0.130 (0.03) | 0.131 (0.04) | 0.130 (0.03) | 0.130 (0.03) | 0.129 (0.04) | 0.130 (0.03) | 0.130 (0.03) |
| 95038† | 22.7 | -0.18 | 0.03 | -0.51 | 0.00 | 0.096 (0.02) | 0.097 (0.03) | 0.102 (0.03) | 0.097 (0.03) | 0.097 (0.03) | 0.103 (0.03) | 0.097 (0.03) | 0.097 (0.03) | 0.101 (0.03) | 0.097 (0.03) | 0.097 (0.03) |
| 95234 | 6.5 | -0.17 | 0.03 | -1.00 | 0.00 | -0.034 (0.04) | -0.033 (0.04) | -0.037 (0.05) | -0.033 (0.04) | -0.034 (0.04) | -0.038 (0.05) | -0.033 (0.04) | -0.034 (0.04) | -0.038 (0.05) | -0.033 (0.04) | -0.034 (0.04) |
| 95236* | 48.6 | 0.54 | 0.29 | 0.82 | -0.15 | -0.117 (0.02) | -0.117 (0.03) | -0.113 (0.03) | -0.117 (0.03) | -0.117 (0.02) | -0.115 (0.03) | -0.117 (0.03) | -0.117 (0.03) | -0.115 (0.03) | -0.117 (0.03) | -0.117 (0.03) |

Table S6: Empirical power rates at significance level $\alpha = 0.05/29$ and $n = 1250$ across 500 replicates in realistic fine-mapping simulation single-variant analysis. Positions marked with $*$ denote causal variants whereas the ones marked with † denote hitchhikers. The remaining ones are non-causal. Positions are truncated to the last 5 digits.

| **$G$ pos.** | **$\beta_{1G}$** | **Complete** | **RDS** | **Par-spec** | | | **A-opt** | | | **D-opt** | | |
|---|---|---|---|---|---|---|---|---|---|---|---|---|
| | | | | **LM** | **GA** | **TZL** | **LM** | **GA** | **TZL** | **LM** | **GA** | **TZL** |
| 85805 | 0.00 | 8.4 | 6.0 | 2.6 | 6.0 | 5.8 | 3.6 | 6.0 | 6.0 | 2.8 | 6.2 | 6.8 |
| 86045† | 0.00 | 76.4 | 56.4 | 30.4 | 56.4 | 57.4 | 21.6 | 55.4 | 54.2 | 20.2 | 55.2 | 56.4 |
| 86762† | 0.00 | 100.0 | 99.8 | 89.2 | 99.8 | 99.8 | 82.8 | 99.8 | 99.8 | 83.6 | 99.8 | 99.8 |
| 86914 | 0.00 | 8.6 | 4.4 | 3.6 | 4.2 | 4.4 | 3.0 | 5.4 | 5.0 | 3.8 | 4.4 | 6.0 |
| 87015 | 0.00 | 0.2 | 0.0 | 0.2 | 0.0 | 0.0 | 0.0 | 0.0 | 0.0 | 0.0 | 0.0 | 0.4 |
| 87765 | 0.00 | 0.4 | 0.2 | 0.2 | 0.2 | 0.0 | 0.0 | 0.4 | 0.0 | 0.0 | 0.4 | 0.2 |
| 88044 | 0.00 | 0.4 | 0.0 | 0.2 | 0.0 | 0.0 | 0.4 | 0.0 | 0.2 | 0.0 | 0.0 | 0.2 |
| 88958† | 0.00 | 83.6 | 64.4 | 27.4 | 62.8 | 65.0 | 24.6 | 64.4 | 63.4 | 21.4 | 64.2 | 62.6 |
| 89015 | 0.00 | 5.4 | 3.2 | 1.2 | 3.2 | 3.2 | 1.0 | 3.4 | 3.2 | 1.2 | 3.4 | 4.2 |
| 89830* | -0.20 | 100.0 | 99.8 | 91.4 | 99.8 | 100.0 | 86.4 | 99.8 | 99.8 | 87.2 | 99.8 | 99.8 |
| 90803† | 0.00 | 77.2 | 56.0 | 34.2 | 57.0 | 58.8 | 23.6 | 56.2 | 55.6 | 22.4 | 58.0 | 57.4 |
| 91143† | 0.00 | 67.4 | 46.4 | 22.2 | 46.6 | 48.0 | 16.4 | 44.8 | 45.4 | 18.0 | 46.8 | 48.4 |
| 91524 | 0.00 | 6.0 | 3.6 | 2.0 | 3.6 | 3.2 | 1.8 | 4.0 | 4.2 | 1.8 | 3.4 | 5.0 |
| 92017 | 0.00 | 7.0 | 4.0 | 2.2 | 4.0 | 3.6 | 3.0 | 3.6 | 3.6 | 2.8 | 4.4 | 4.0 |
| 93161 | 0.00 | 0.8 | 0.6 | 0.0 | 0.6 | 0.8 | 0.2 | 0.8 | 0.2 | 0.4 | 1.0 | 0.8 |
| 93211 | 0.00 | 7.0 | 5.6 | 1.4 | 5.6 | 5.0 | 1.4 | 5.4 | 5.6 | 1.0 | 5.6 | 4.8 |
| 93324* | 0.12 | 3.0 | 1.4 | 0.4 | 1.4 | 1.4 | 0.6 | 1.6 | 1.2 | 1.2 | 1.6 | 0.8 |
| 93886 | 0.00 | 0.2 | 0.4 | 0.0 | 0.4 | 0.2 | 0.2 | 0.4 | 0.2 | 0.2 | 0.4 | 0.0 |
| 93897 | 0.00 | 20.0 | 11.0 | 4.2 | 11.0 | 10.2 | 4.4 | 11.0 | 9.6 | 4.2 | 11.6 | 9.2 |
| 93901 | 0.00 | 18.0 | 11.6 | 3.0 | 11.8 | 11.2 | 3.0 | 11.4 | 10.4 | 4.2 | 11.2 | 11.6 |
| 93935† | 0.00 | 80.6 | 58.0 | 37.2 | 59.4 | 60.0 | 24.6 | 57.4 | 56.8 | 24.4 | 58.6 | 59.6 |
| 94192† | 0.00 | 80.6 | 59.0 | 38.8 | 59.2 | 61.2 | 24.0 | 56.2 | 58.6 | 25.8 | 57.2 | 62.0 |
| 94212 | 0.00 | 3.8 | 1.6 | 0.4 | 1.6 | 1.2 | 0.8 | 1.8 | 2.4 | 0.2 | 1.8 | 1.0 |
| 94244 | 0.00 | 0.2 | 0.4 | 0.0 | 0.4 | 0.2 | 0.2 | 0.4 | 0.2 | 0.2 | 0.4 | 0.0 |
| 94528 | 0.00 | 0.2 | 0.4 | 0.0 | 0.4 | 0.2 | 0.2 | 0.4 | 0.2 | 0.2 | 0.4 | 0.0 |
| 94990* | 0.25 | 84.4 | 65.0 | 34.8 | 65.2 | 65.8 | 26.2 | 63.0 | 64.6 | 26.4 | 66.2 | 64.0 |
| 95038† | 0.00 | 79.6 | 58.0 | 36.2 | 57.0 | 60.2 | 24.4 | 57.0 | 56.8 | 24.6 | 57.8 | 59.0 |
| 95234 | 0.00 | 0.2 | 0.2 | 0.6 | 0.4 | 0.0 | 1.2 | 0.2 | 0.6 | 0.2 | 0.0 | 0.4 |
| 95236* | -0.15 | 95.2 | 86.6 | 40.2 | 86.8 | 84.0 | 40.6 | 86.4 | 84.6 | 36.8 | 86.4 | 81.8 |

##### S1.2 Figures

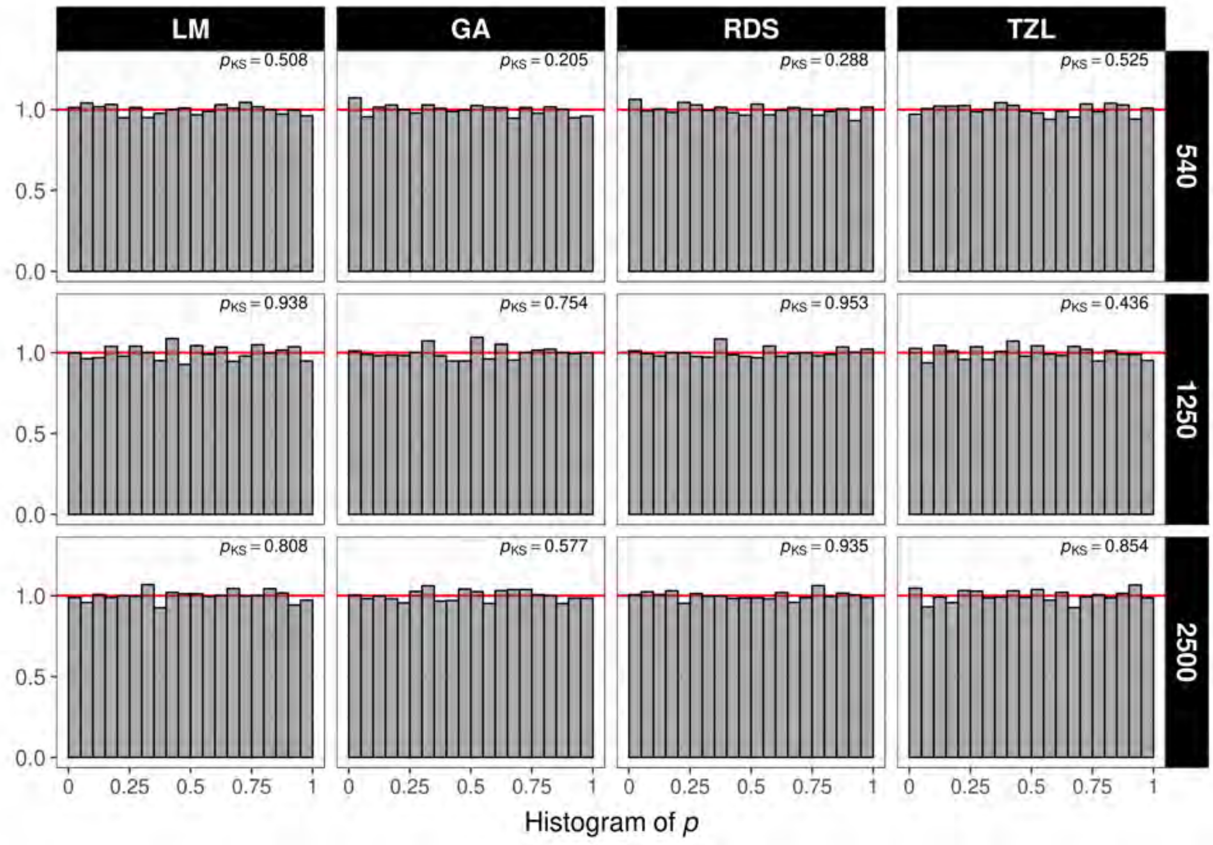

Figure S1: $P$-value histograms under the null hypothesis and LR test in the comparison against ranked designs across optimality criteria and phase 2 sample sizes ($n = 540, 1250, 2500$). Column facets correspond to each of the studied designs. Row facets correspond to each phase 2 sample size. The top right values in each facet denote the $p$-value of the Kolmogorov-Sminrnov test that the observed $p$-values follow the expected uniform distribution.

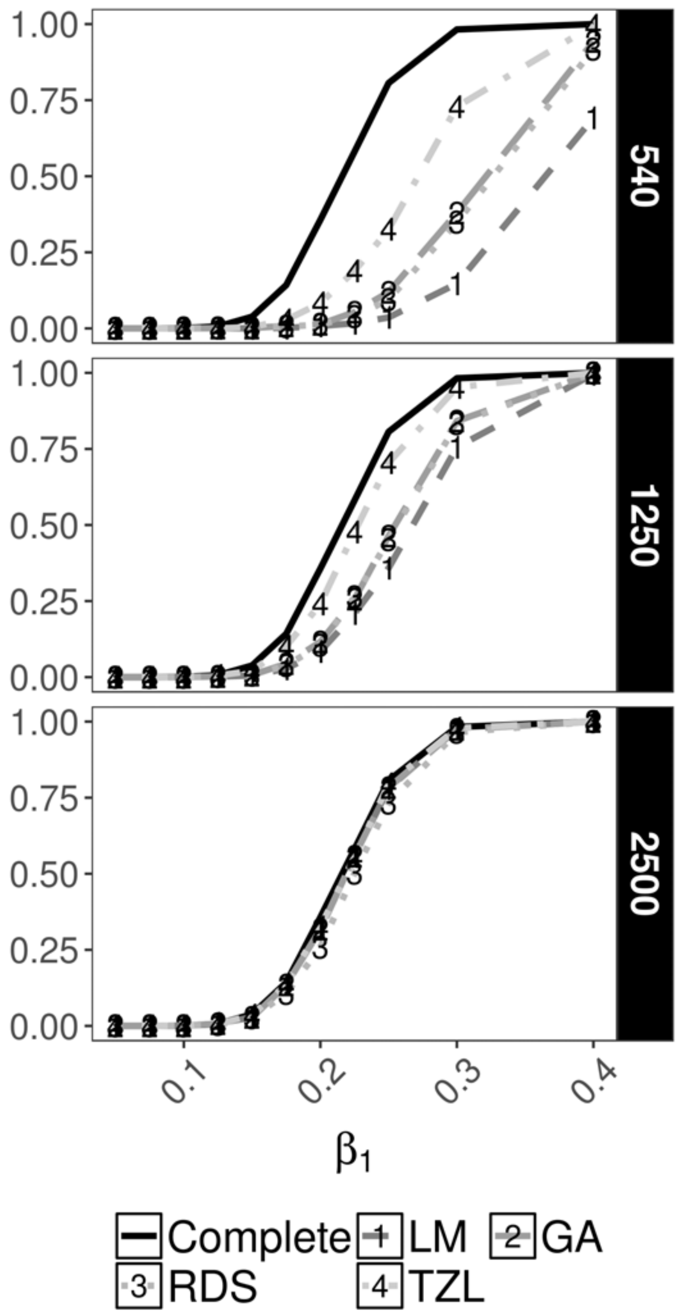

Figure S2: Power curves under the alternative $\beta_1 \neq 0$ at $\alpha = 1 \times 10^{-8}$ for testing $\mathrm{H}_0 : \beta_1 = 0$ under the LR test and parameter-specific criterion comparing designs across phase 2 sample sizes. Row facets denote different phase 2 sample sizes $n = 540, 1250, 2500$.

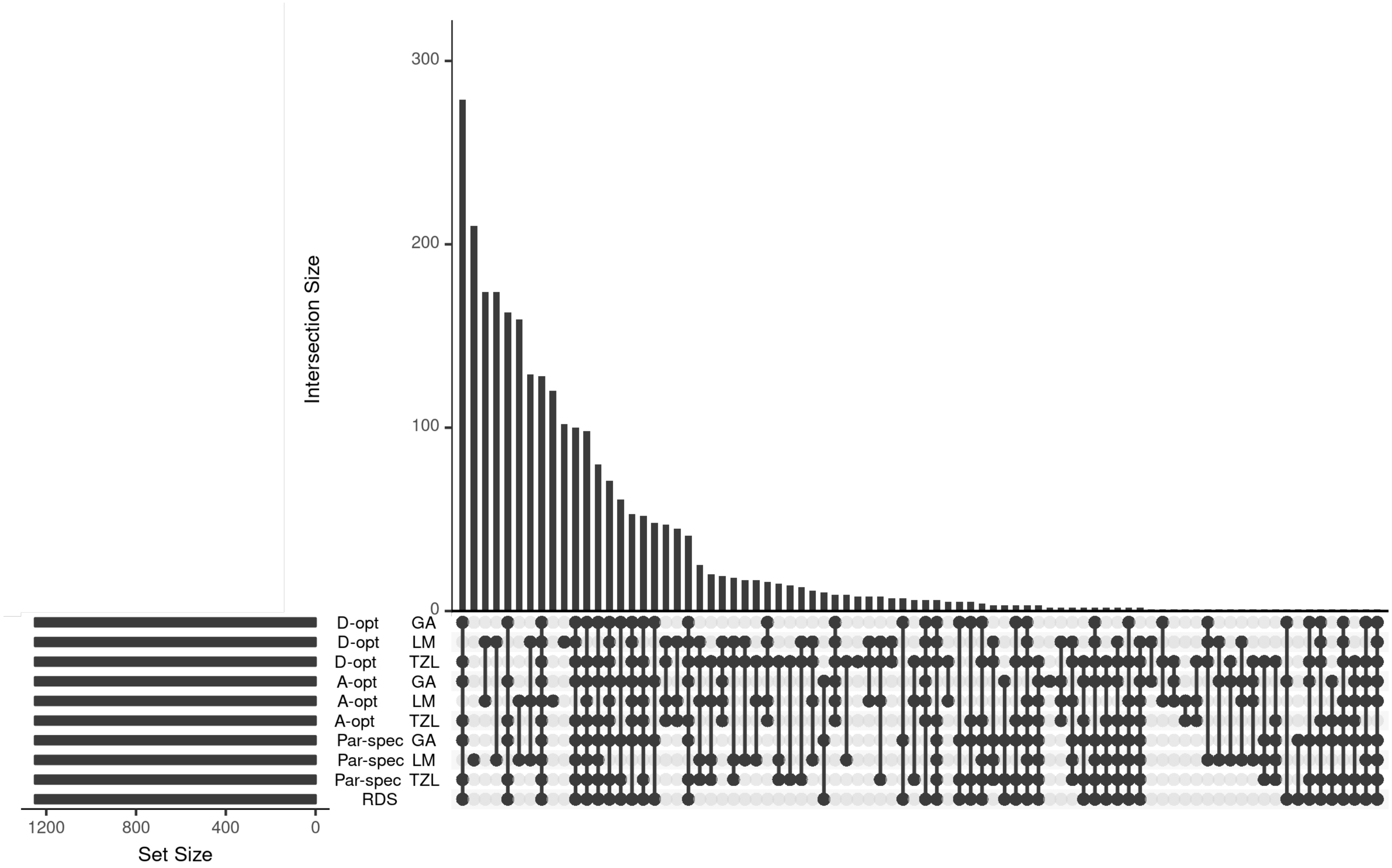

Figure S3: Upset plots for a single replicate in the realistic simulation to quantify the intersection sizes across studied designs and optimality criteria when $n = 1250$. Each bar denotes the size of a given intersection highlighted in the x-axis.

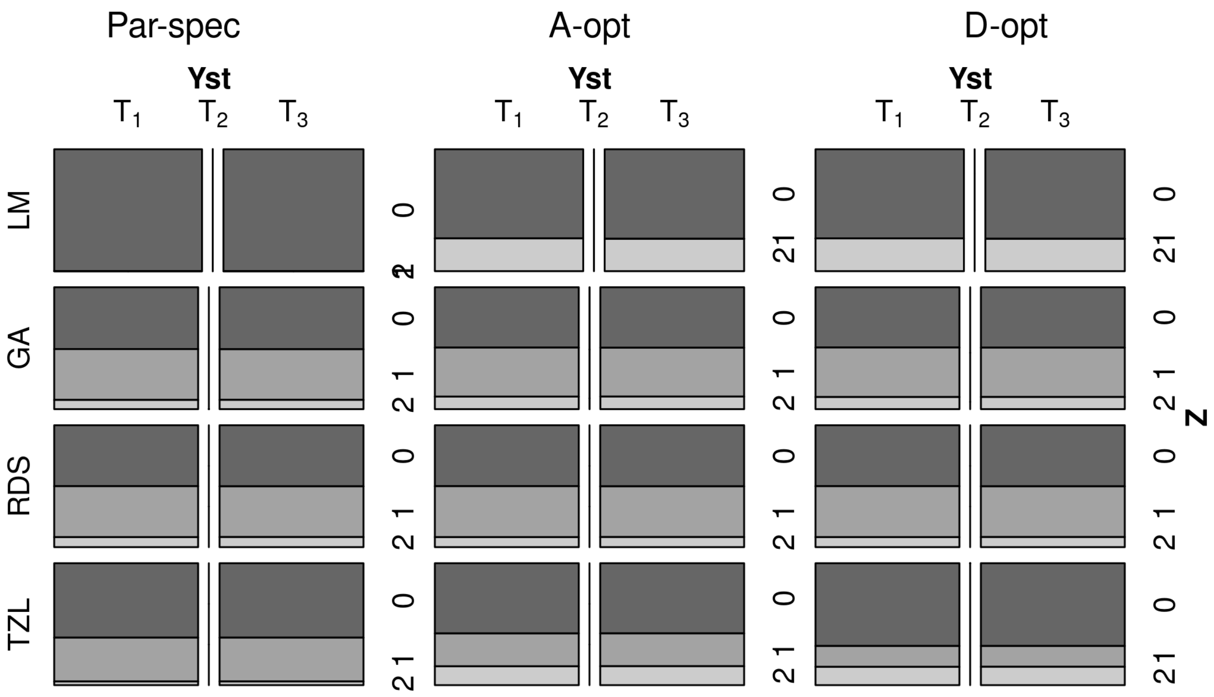

Figure S4: Mosaic plots with the average strata sizes across replicates of the resulting designs in the realistic simulation across optimality criteria (parameter-specific, A-/D-optimality) for $n = 1250$.

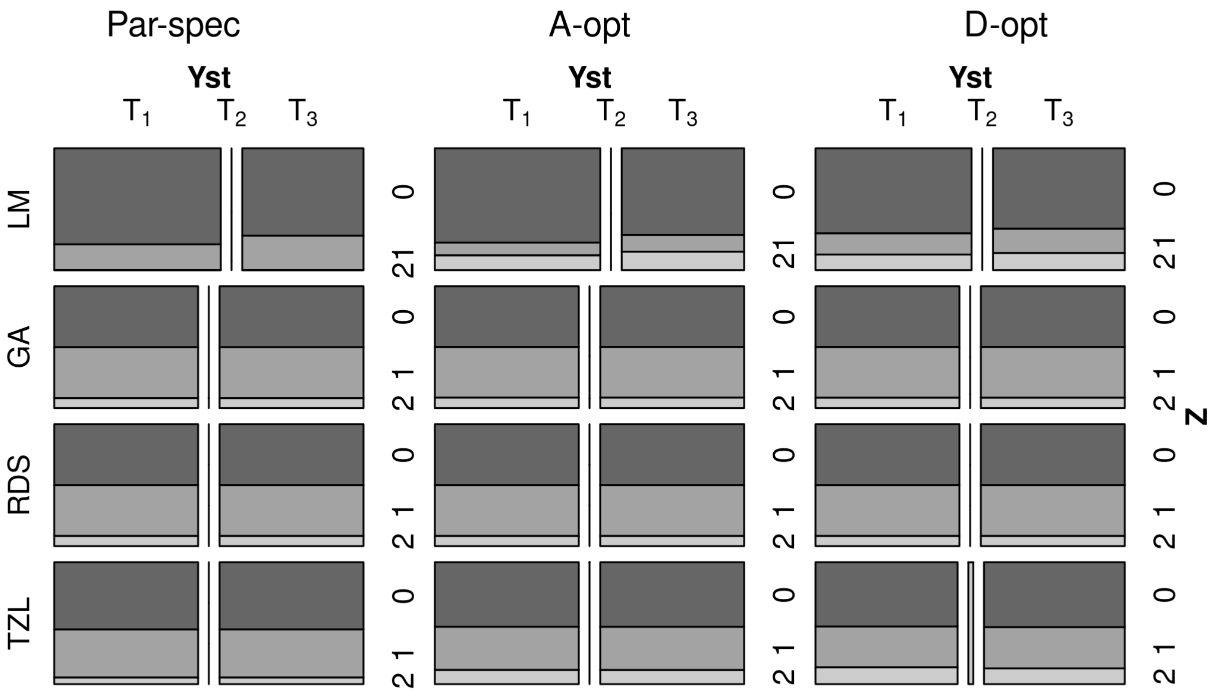

Figure S5: Mosaic plots with the average strata sizes across replicates of the resulting designs in the realistic simulation across optimality criteria (parameter-specific, A-/D-optimality) for $n = 2500$.

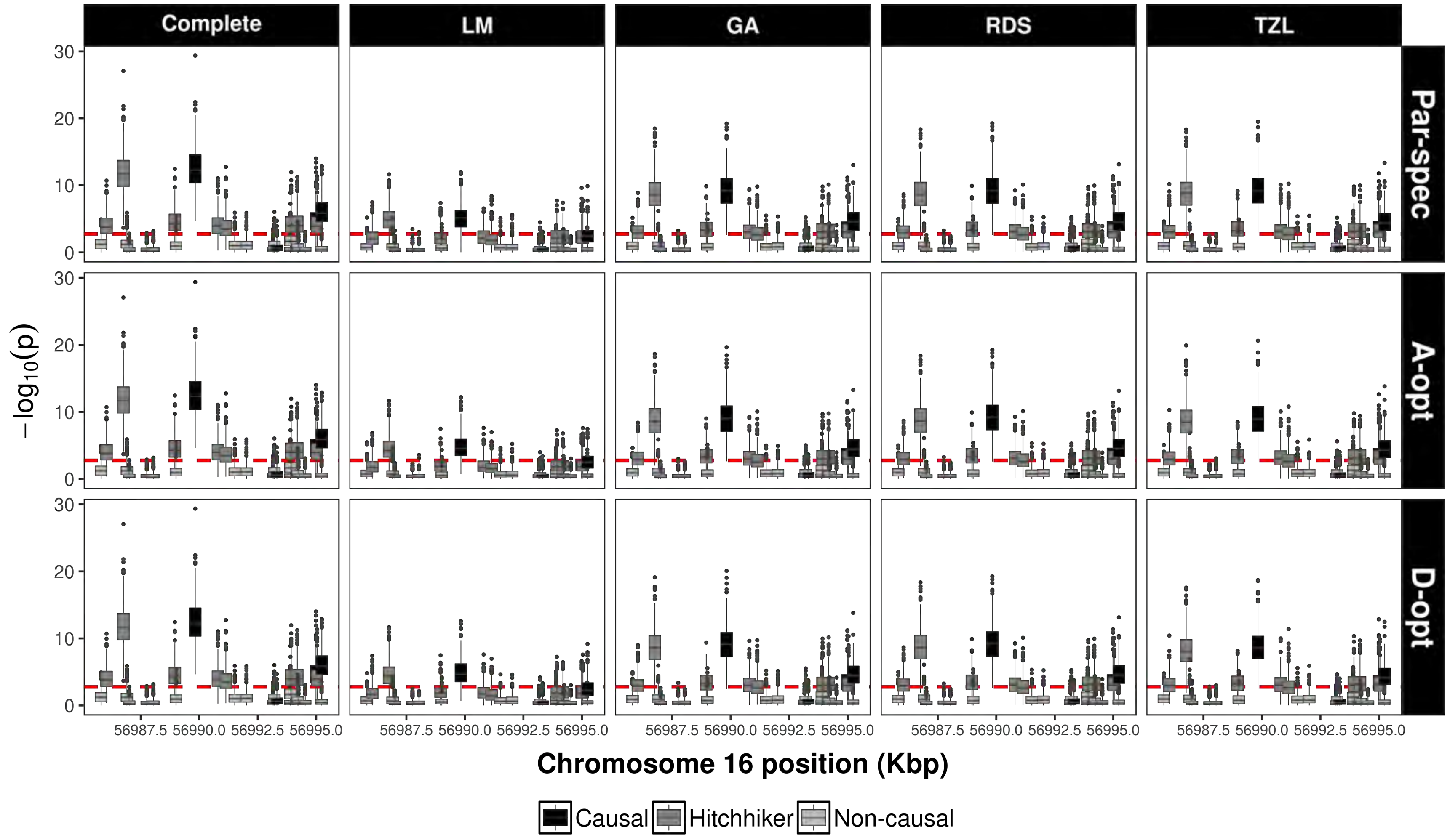

Figure S6: Boxplots of the $(-\log_{10})$ $p$-values across 500 replicates in the fine-mapping simulation single-variant analyses for a phase 2 sample size of $n = 1250$ across optimality criteria (for LM, GA, and TZL only): parameter-specific, A- and D- optimality in each row facet. Each column facet corresponds to the complete data analysis and studied designs respectively. The dashed line corresponds to a Bonferroni-corrected significance threshold of $\alpha = 0.05/29$.

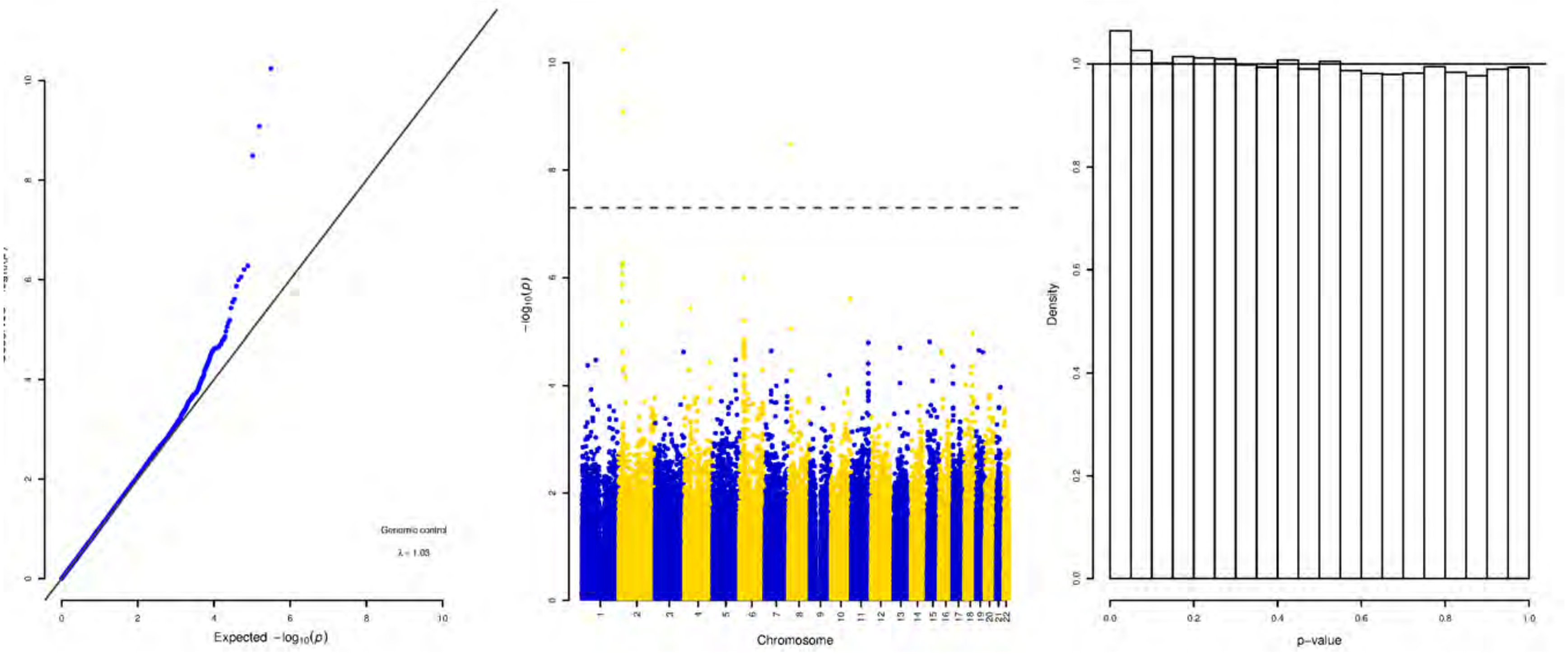

Figure S7: Genome-wide association results on the log-transformed triglyceride levels on 5300 subjects from the North Finland Birth Cohort of 1966. Left panel displays the quantile-quantile plot for the -$\log_{10}$($p$-values), the middle panel denotes the Manhattan plot across the autosomes, the right panel denotes the histogram of the $p$-values to assess the uniform distribution assumption. Results only include SNPs with MAF greater or equal than 5%

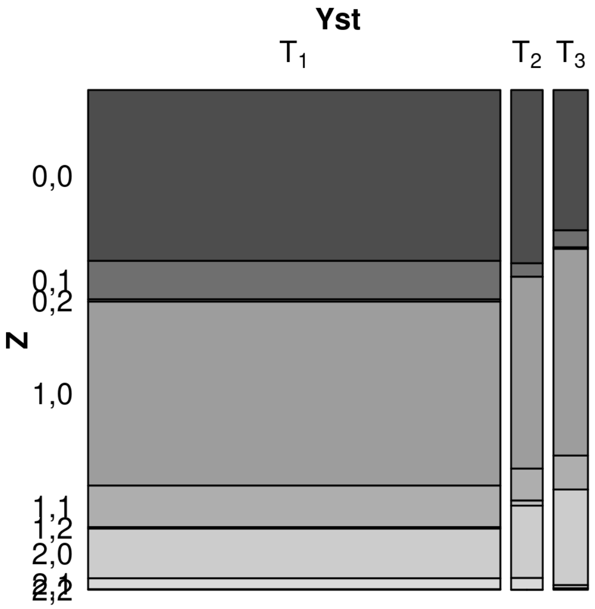

Figure S8: Mosaic plots for the strata distribution in NFBC66 phase 1 data.

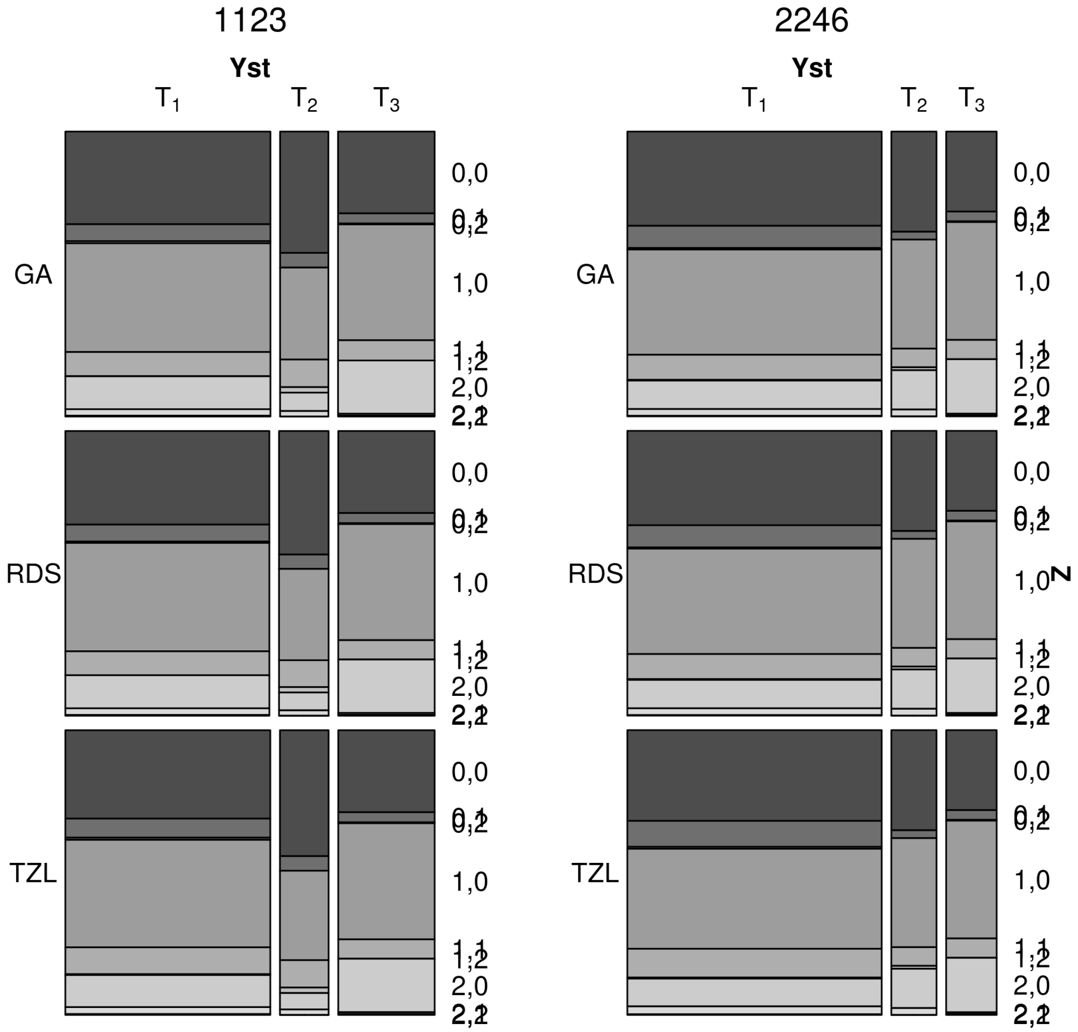

Figure S9: Mosaic plots for the strata distribution across studied designs GA, RDS, and TZL in NFBC66 data across different phase 2 sample sizes. Each column shows the corresponding designs for each phase 2 sample size.

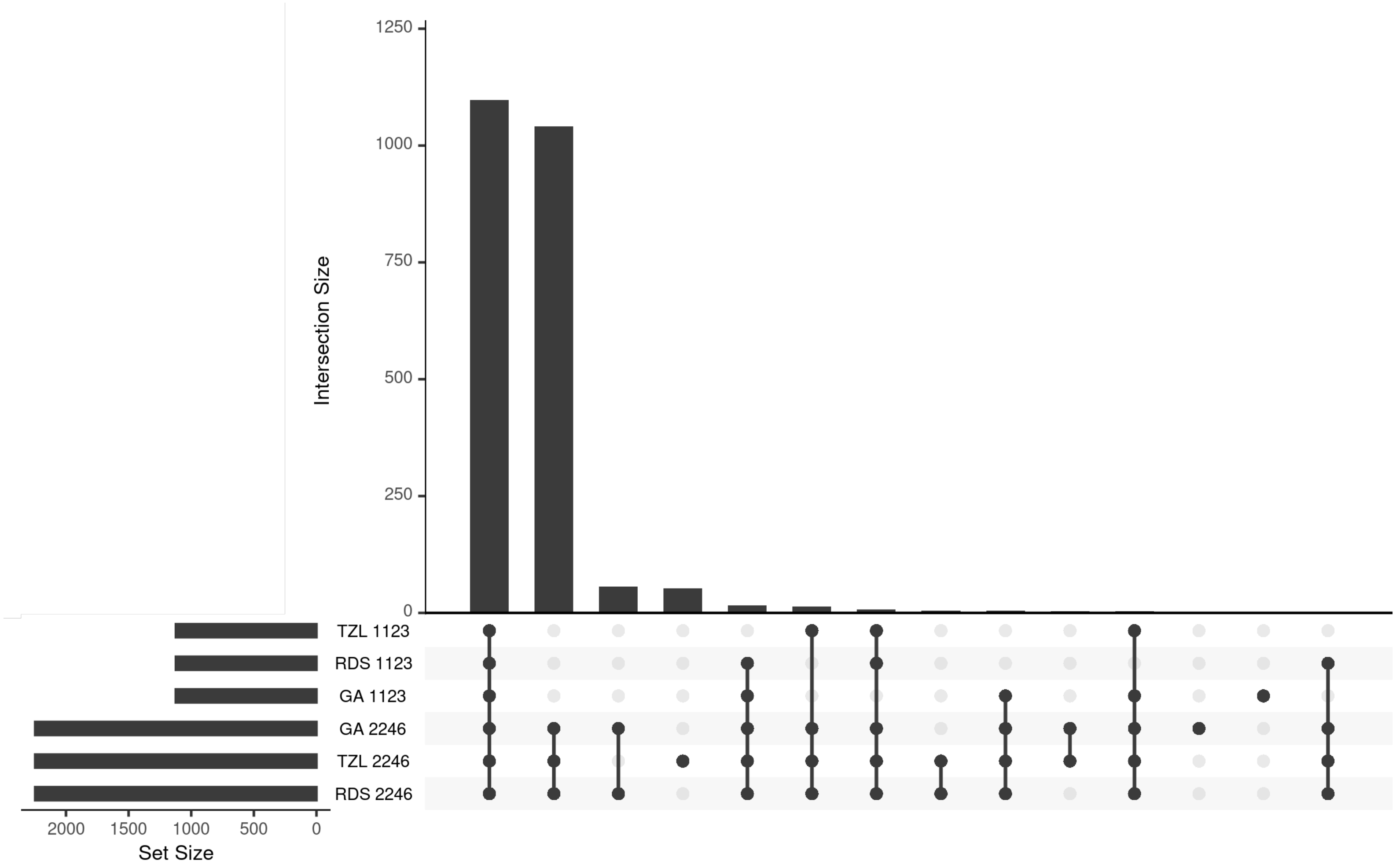

Figure S10: Upset plots for the NFCBB66 data to quantify the intersection sizes across studied designs and phase 2 sample sizes. Each bar denotes the size of a given intersection highlighted in the x-axis.

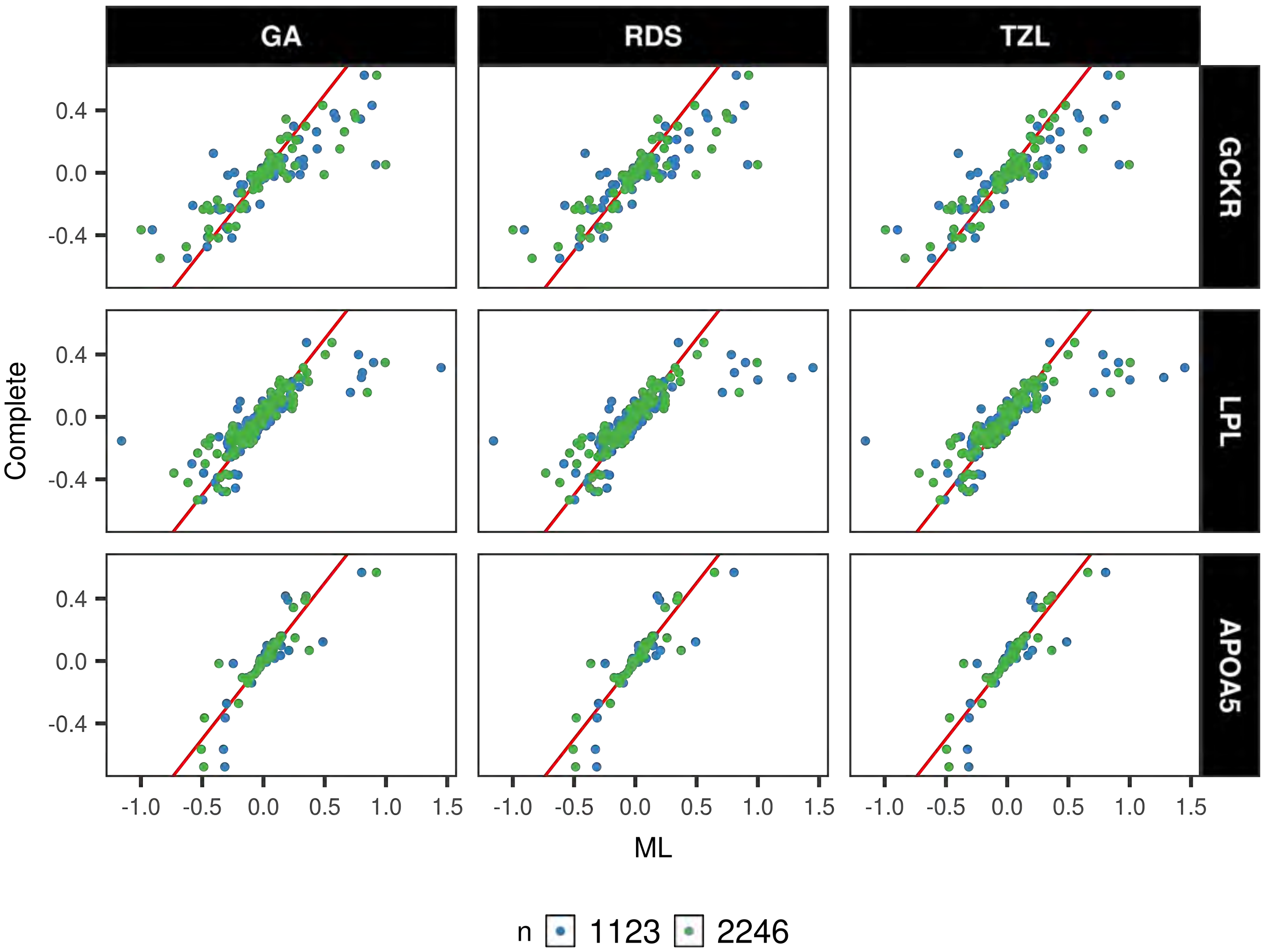

Figure S11: Beta-Beta plots for the ML analyses under the studied designs compared to the complete data analysis in the NFCBB66 data. Column facets shows one studied design whereas row facets denote each loci of interest.

### S2 Additional considerations in the optimization strategies

#### S2.1 Details on LM

Minimization of the LM equation (3) involves constrained optimization strategies; in particular, equation (3) can be solved using linear inequality constraints on the $\pi$'s. Given the complex nature of the functions involved in the optimality criteria, a derivative-free approach is more appealing than directly specifying the gradient. Although in principle the standard Nelder-Mead algorithm within function *constrOptim* in R language can be used, we found that the COBYLA routine in R package *nloptr* performed better.

#### S2.2 Details on GA

To determine the most appropriate tuning parameter configuration for simulations under GA, we evaluated various combinations of the control parameters: elitism rate, mutation rate, tournament rate, population size and number of generations (data not shown). Intuitively, the longer the number of generations the better the chances of reaching the optimum, however, evaluation across multiple replicates in simulations restricts the computing time. Since there is a trade-off between the elapsing time of the GA and the number of replications we aim to carry out, we only evaluated combinations that were able to be computed in acceptable amount of time (no longer than 3 minutes on a 3.1 GHz Intel Core i5 processor). For these combinations, we calculated the values of the optimality criterion for multiple initializations of the GA in a fixed test dataset. Based on our observations we set up the tuning parameters as population size, $\mathcal{M} = 60$; number of generations $\mathcal{H} = 300$; size of selection tournament, $\mathcal{T} = 0.90 \times \mathcal{M}$; number of elites, $\mathcal{E} = 0.90 \times \mathcal{M}$); and mutation rate, $r = 0.001$.

### S3 Simulations comparing heuristic designs

The purpose of this additional set of simulations is twofold: 1) compare the proposed designs against previously heuristically defined sampling designs, namely balanced and combined, and 2) assess the performance of the proposed designs when considering the variance-covariance matrix, $\mathbb{J}(\Phi)^{-1}$, instead of $\mathbb{V}_1^{-1}$ (Section 2.4.4). For comparability, the results from this set of simulations are obtained using the same simulated data from Section (3.1) of the main text.

As in Section 3, we evaluate the performance of the the proposed designs (LM and GA) against the two heuristic designs (balanced and combined) across three statistical tests (Wald, likelihood ratio (LR) and score) under the parameter-specific criterion. In addition, relative empirical power, asymptotic and empirical standard errors (rEP, rASE and rESE, respectively) of $\hat{\beta}_1$ for each design over that of the complete data are also computed.

#### S3.1 Heuristic designs

Given $L \times M$ strata defined by the auxiliary covariate $Z$ and a discrete version of the QT, $Y$, with $l = 1, \ldots, L = 3$ groups for $Z$ and $m = 1, \ldots, M = 3$ groups for $Y$, a balanced design aims for approximately equal sample sizes across strata in phase 2 data. On the other hand, a combined design considers a balanced allocation on the auxiliary covariate, $Z$, with samples only being allocated on the extremes of the QT, $Y$ [1]. In particular, even though the balanced design has been regarded as “near optimal” under binary outcomes, its properties are less understood for QTs. We also note that depending on $n$ and the phase 1 sample distribution among strata, it may be unfeasible to select the exact same phase 2 sample sizes for these designs as some strata may be completely exhausted. Consequently, the unallocated sample is selected at random into the remaining strata. Lastly, both the balanced and combined are sampling strategies, thus, are also subject to sampling variability. Hence, drawing a fixed number of subsamples and select the draw that minimizes $\Lambda(\cdot)$ can help in reducing the extra variation introduced by the sampling.

#### S3.2 Specifying design values

In this case, the specification of $\boldsymbol{\beta}_{des}$, the design parameters, corresponds to $(\beta_0, \beta_1, \beta_z = 0)$ where $\beta_0$ and $\beta_1$ are the values used for data generation. Similarly, we specify $\boldsymbol{p}_{des}$, the design haplotype distribution between $G$ and $Z$, under HWE by estimating $q_Z$ from the phase 1 sample and designating $q_G$ and $r$ to be the equal to the generating values. This specification for $\theta_{des} = (\boldsymbol{\beta}_{des}^t, \boldsymbol{p}_{des}^t)^t$ corresponds to the case where an investigator accurately specifies the parameter effect and haplotype distribution at design stage, which is of course idealistic. In the case of the balanced and combined designs, $\theta_{des}$ is used to select a drawn sample that minimizes $\Lambda(\cdot)$. Lastly, specifying $\beta_z = 0$ at design stage reflects the belief that the GWAS SNP is not truly causal but can detect an induced association with a causal variant in a marginal model.

#### S3.3 Results

The distribution of the average strata sizes in the LM, GA, balanced and combined designs aids in identifying relationships among the strata and genetic effect sizes, this can be visualized via mosaic plots (Figure S12). There, when focusing on the parameter-specific criterion comparing LM and GA against the balanced and combined designs for the smallest and largest studied genetic effects ($\beta_1 = 0.1$ and $\beta_1 = 0.7$), we observe: (1) LM varies considerably across $\beta_1$ values while GA appears to display more stable strata distribution, (2) LM and GA designs reach similar average distributions when $n = 2500$, and (3) the strata distribution of the heuristic suffer little change between genetic effect sizes (Figure S12).

At $\alpha$ =1%, type 1 error (T1E) rates for the proposed designs are anti-conservative under the Wald test and conservative under the score test while remaining at acceptable levels under the LR test, this is particularly true for LM; the T1E rates stabilize as $n$ increases, however (Tables S7). These results suggest that using $\mathbb{V}_1^{-1}$ instead of $\mathbb{J}^{-1}$ for LM and GA helps to better control T1E across all tests. Closer inspection of the $p$-value distribution under LR displays no gross departure from the expected uniform distribution (Figures S13).

Empirical bias $(\hat{\beta}_1 - \beta_1)$ is well centered around zero overall and decreases as $n$ increases, as expected (Figure S14). The exception of this is the LM design, which exhibits biased estimates when $\beta_1 = 0.7$. Notably, LM exhibit larger bias variability compared to the other designs, especially with smaller $n$ (Figure S14).

Power curves under the LR test for $\alpha = 1 \times 10^{-5}$ exhibit gains in power for GA compared to balanced and combined designs across values of $n$. LM only shows good performance for the largest phase 2 sample size ($n = 2500$) (FigureS15). Notably, only LM and GA reach similar power as the complete data analysis when $n = 2500$ (Figure S15). An overall comparison of the designs under shows that the power of LM and GA is larger for the comparison against ranked designs, i.e. when using $\mathbb{V}_1^{-1}$ and $\boldsymbol{\beta}_{des}$ is defined under the null are used in the optimization (Figure S15). The rEP is highest for GA followed by the combined, balanced designs across all values of $n$ whereas LM only achieves rEP comparable to GA when $n = 2500$ (Table S8).

The (r)ASE and (r)ESE for GA has better agreement and fall closer to 1 as $n$ increases when compared to the heuristic designs indicating higher relative efficiency (Tables S9-S10). The performance of LM substantially improves with $n$, with the ASE noticeably underestimating the ESE possibly due to sampling variability. Lastly, the 95% confidence intervals show adequate coverage across most designs, the exception is LM when $n = 540, 1250$ in comparisons against heuristic designs (Table S11).

###### S3.3.1 Tables

Table S7: Type 1 error (T1E) ($\alpha = 1\%$) across LM, GA, balanced, and combined designs, phase 2 sample sizes ($n = 540, 1250, 2500$) and statistical tests under a parameter-specific criterion. Each entry represents 11250 replicates. The rest of the simulation parameters correspond to $q_G = 0.2$, $q_Z = 0.3$, $r = 0.75$, $\beta_0 = 2$, $\sigma^2 = 1$, $N = 5000$. The complete data T1E is 1.16 for Wald/LR tests and 1.14 for the score test.

| $n$ | Test | LM | GA | Bal | Comb |
|---|---|---|---|---|---|
| 540 | Wald | 10.48 | 1.17 | 1.99 | 1.42 |
| | LR | 1.03 | 0.94 | 1.06 | 0.98 |
| | Score | 0.17 | 0.90 | 0.90 | 0.86 |
| 1250 | Wald | 4.91 | 1.12 | 1.14 | 1.13 |
| | LR | 1.01 | 1.05 | 0.96 | 1.03 |
| | Score | 0.22 | 1.02 | 0.91 | 0.99 |
| 2500 | Wald | 1.02 | 1.03 | 1.11 | 1.07 |
| | LR | 0.95 | 0.99 | 1.05 | 1.03 |
| | Score | 0.93 | 0.97 | 1.02 | 1.00 |

Table S8: Relative empirical power (rEP), calculated as the ratio of the empirical power of each studied design (LM, GA, balanced, combined) over that of the complete data, across phase 2 sample sizes, and effect sizes under the LR test ($\alpha = 1 \times 10^{-8}$). Phase 1 sample size is $N = 5000$ whereas phase 2 sample size is $n = 540, 1250, 2500$. These results exclude $\beta_1 > 0.5$ since power had already reached 100%. The rest of the simulation parameters correspond to $q_G = 0.2$, $q_Z = 0.3$, $r = 0.75$, $\beta_0 = 2$, and $\sigma^2 = 1$.

| $n$ | $\beta_1$ | LM | GA | Bal | Comb |
|---|---|---|---|---|---|
| 540 | 0.100 | 0.0 | 0.0 | 0.0 | 0.0 |
| | 0.125 | 0.0 | 12.5 | 0.0 | 0.0 |
| | 0.150 | 0.0 | 0.0 | 0.0 | 0.0 |
| | 0.175 | 0.0 | 1.1 | 0.0 | 0.0 |
| | 0.200 | 0.0 | 1.1 | 0.0 | 0.0 |
| | 0.225 | 0.0 | 2.5 | 0.3 | 0.8 |
| | 0.250 | 0.0 | 4.1 | 0.2 | 0.8 |
| | 0.300 | 0.0 | 14.0 | 1.1 | 3.1 |
| | 0.400 | 0.1 | 65.2 | 10.9 | 35.8 |
| | 0.500 | 0.3 | 97.6 | 51.8 | 84.4 |
| 1250 | 0.100 | 0.0 | 0.0 | 0.0 | 0.0 |
| | 0.125 | 0.0 | 25.0 | 0.0 | 12.5 |
| | 0.150 | 0.0 | 4.2 | 0.0 | 4.2 |
| | 0.175 | 0.6 | 4.5 | 1.7 | 2.8 |
| | 0.200 | 1.3 | 9.9 | 1.1 | 6.3 |
| | 0.225 | 3.9 | 17.4 | 3.7 | 10.9 |
| | 0.250 | 5.8 | 24.8 | 6.6 | 15.5 |
| | 0.300 | 5.2 | 54.2 | 19.8 | 39.4 |
| | 0.400 | 4.5 | 97.4 | 77.4 | 93.2 |
| | 0.500 | 20.4 | 100.0 | 99.3 | 100.0 |
| 2500 | 0.100 | 100.0 | 100.0 | 100.0 | 0.0 |
| | 0.125 | 100.0 | 112.5 | 12.5 | 50.0 |
| | 0.150 | 72.9 | 83.3 | 12.5 | 31.2 |
| | 0.175 | 84.8 | 87.6 | 6.2 | 30.3 |
| | 0.200 | 82.3 | 87.2 | 17.0 | 33.4 |
| | 0.225 | 92.4 | 94.5 | 26.0 | 52.2 |
| | 0.250 | 94.3 | 95.9 | 33.0 | 61.9 |
| | 0.300 | 98.7 | 99.1 | 65.9 | 88.9 |
| | 0.400 | 100.0 | 100.0 | 99.0 | 100.0 |
| | 0.500 | 100.0 | 100.0 | 100.0 | 100.0 |

Table S9: Asymptotic/Empirical standard errors (ASE and ESE, respectively) comparing against heuristic designs across effect sizes ($\beta_1$), and phase 2 sample sizes. Phase 1 sample size is $N = 5000$ whereas phase 2 sample size is $n = 540, 1250, 2500$.

| $n$ | $\beta_1$ | ASE | | | | | ESE | | | | |
|---|---|---|---|---|---|---|---|---|---|---|---|
| | | **Complete** | **LM** | **GA** | **Bal** | **Comb** | **Complete** | **LM** | **GA** | **Bal** | **Comb** |
| 540 | 0.0 | 0.025 | 0.077 | 0.058 | 0.076 | 0.065 | 0.025 | 0.113 | 0.058 | 0.079 | 0.066 |
| | 0.1 | 0.038 | 0.076 | 0.063 | 0.088 | 0.075 | 0.038 | 0.099 | 0.063 | 0.088 | 0.077 |
| | 0.3 | 0.038 | 0.068 | 0.063 | 0.080 | 0.070 | 0.038 | 0.125 | 0.066 | 0.086 | 0.073 |
| | 0.5 | 0.038 | 0.073 | 0.060 | 0.070 | 0.065 | 0.037 | 0.137 | 0.062 | 0.079 | 0.070 |
| | 0.7 | 0.038 | 0.055 | 0.057 | 0.062 | 0.060 | 0.037 | 0.119 | 0.062 | 0.072 | 0.068 |
| 1250 | 0.0 | 0.025 | 0.053 | 0.042 | 0.051 | 0.044 | 0.025 | 0.068 | 0.042 | 0.052 | 0.045 |
| | 0.1 | 0.038 | 0.051 | 0.050 | 0.061 | 0.054 | 0.038 | 0.060 | 0.050 | 0.060 | 0.053 |
| | 0.3 | 0.038 | 0.056 | 0.051 | 0.059 | 0.053 | 0.038 | 0.068 | 0.051 | 0.061 | 0.055 |
| | 0.5 | 0.038 | 0.052 | 0.050 | 0.057 | 0.052 | 0.037 | 0.080 | 0.050 | 0.059 | 0.054 |
| | 0.7 | 0.038 | 0.045 | 0.049 | 0.054 | 0.051 | 0.037 | 0.073 | 0.050 | 0.058 | 0.053 |
| 2500 | 0.0 | 0.025 | 0.035 | 0.035 | 0.037 | 0.032 | 0.025 | 0.035 | 0.035 | 0.038 | 0.032 |
| | 0.1 | 0.038 | 0.038 | 0.038 | 0.049 | 0.043 | 0.038 | 0.038 | 0.038 | 0.049 | 0.043 |
| | 0.3 | 0.038 | 0.037 | 0.038 | 0.048 | 0.043 | 0.038 | 0.038 | 0.039 | 0.048 | 0.044 |
| | 0.5 | 0.038 | 0.035 | 0.038 | 0.047 | 0.043 | 0.037 | 0.036 | 0.038 | 0.047 | 0.043 |
| | 0.7 | 0.038 | 0.033 | 0.038 | 0.046 | 0.043 | 0.037 | 0.036 | 0.038 | 0.046 | 0.043 |

Table S10: Relative asymptotic/empirical standard error (rASE/rESE, respectively) comparing heuristic designs across effect sizes ($\beta_1$) and phase 2 sample sizes. The ASE/ESE of each method (LM, GA, Balanced, Combined) was calculated against the ASE/ESE of the complete data. Phase 1 sample size is $N = 5000$ whereas phase 2 sample size is $n = 540, 1250, 2500$.

| $n$ | $\beta_1$ | rASE | | | | rESE | | | |
|---|---|---|---|---|---|---|---|---|---|
| | | **LM** | **GA** | **Bal** | **Comb** | **LM** | **GA** | **Bal** | **Comb** |
| 540 | 0.0 | 3.048 | 2.289 | 3.033 | 2.567 | 4.491 | 2.281 | 3.129 | 2.610 |
| | 0.1 | 2.000 | 1.662 | 2.333 | 1.985 | 2.647 | 1.679 | 2.353 | 2.049 |
| | 0.3 | 1.791 | 1.655 | 2.116 | 1.862 | 3.304 | 1.733 | 2.263 | 1.919 |
| | 0.5 | 1.938 | 1.593 | 1.849 | 1.706 | 3.710 | 1.668 | 2.138 | 1.893 |
| | 0.7 | 1.451 | 1.499 | 1.648 | 1.599 | 3.199 | 1.662 | 1.945 | 1.842 |
| 1250 | 0.0 | 2.094 | 1.669 | 2.043 | 1.747 | 2.712 | 1.683 | 2.060 | 1.767 |
| | 0.1 | 1.349 | 1.329 | 1.603 | 1.420 | 1.599 | 1.330 | 1.610 | 1.399 |
| | 0.3 | 1.472 | 1.337 | 1.563 | 1.411 | 1.793 | 1.342 | 1.613 | 1.444 |
| | 0.5 | 1.367 | 1.323 | 1.497 | 1.386 | 2.164 | 1.342 | 1.581 | 1.454 |
| | 0.7 | 1.182 | 1.286 | 1.423 | 1.346 | 1.965 | 1.338 | 1.561 | 1.424 |
| 2500 | 0.0 | 1.388 | 1.378 | 1.479 | 1.250 | 1.387 | 1.376 | 1.493 | 1.256 |
| | 0.1 | 1.012 | 1.012 | 1.285 | 1.147 | 1.003 | 1.004 | 1.296 | 1.149 |
| | 0.3 | 0.979 | 1.017 | 1.267 | 1.143 | 1.000 | 1.023 | 1.275 | 1.165 |
| | 0.5 | 0.927 | 1.017 | 1.237 | 1.136 | 0.976 | 1.025 | 1.261 | 1.171 |
| | 0.7 | 0.881 | 1.017 | 1.205 | 1.127 | 0.972 | 1.022 | 1.233 | 1.166 |

Table S11: 95% coverage probabilities in the simulation comparing against heuristic designs for $\beta_1$ across effect sizes and phase 2 sample sizes. Phase 1 sample size is $N = 5000$ whereas phase 2 sample size is $n = 540, 1250, 2500$.

| $n$ | $\beta_1$ | **LM** | **GA** | **Bal** | **Comb** |
|---|---|---|---|---|---|
| 540 | 0.0 | 80.6 | 94.9 | 93.4 | 94.2 |
| | 0.1 | 86.0 | 94.7 | 93.8 | 94.3 |
| | 0.3 | 71.0 | 94.0 | 93.6 | 94.1 |
| | 0.5 | 72.3 | 94.5 | 91.1 | 93.4 |
| | 0.7 | 57.5 | 93.0 | 91.2 | 92.0 |
| 1250 | 0.0 | 87.3 | 94.6 | 94.7 | 94.6 |
| | 0.1 | 90.8 | 95.0 | 95.3 | 95.4 |
| | 0.3 | 89.8 | 94.9 | 94.1 | 94.2 |
| | 0.5 | 77.1 | 94.7 | 93.9 | 94.7 |
| | 0.7 | 58.1 | 93.8 | 93.2 | 94.0 |
| 2500 | 0.0 | 95.1 | 95.0 | 94.9 | 94.7 |
| | 0.1 | 95.4 | 95.1 | 95.8 | 94.8 |
| | 0.3 | 93.0 | 94.2 | 94.2 | 94.3 |
| | 0.5 | 91.9 | 95.4 | 94.6 | 94.2 |
| | 0.7 | 70.7 | 95.0 | 95.5 | 94.7 |

##### S3.3.2 Figures

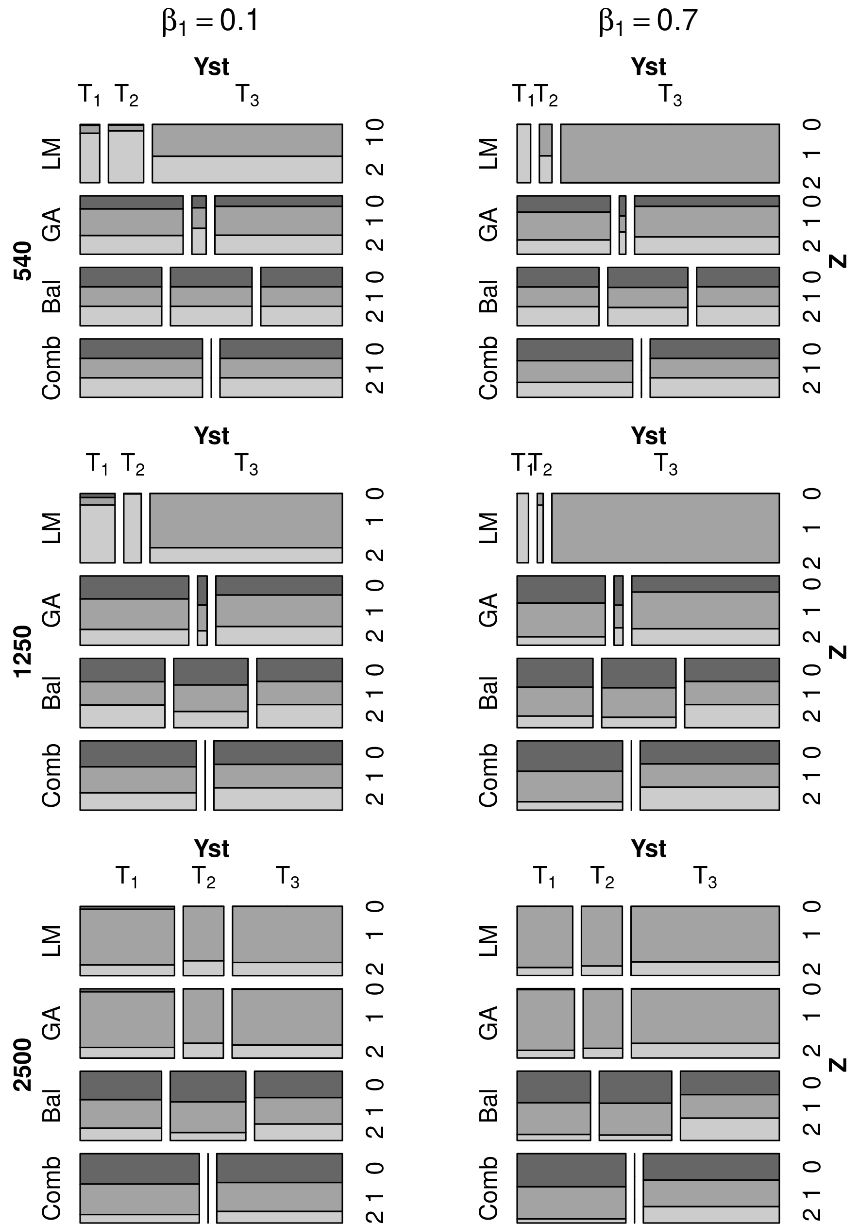

Figure S12: Mosaic plots with the average strata sizes across replicates for the proposed designs against heuristic designs across phase 2 sample sizes ($n = 540, 1250, 2500$) under the parameter-specific criterion. Averages were taken from the resulting designs in the main simulation study for the two most extreme values of $\beta_1$ (0.1,0.7). For LM and GA, $\mathbb{J}(\Phi)^{-1}$ was used in the optimization criterion.

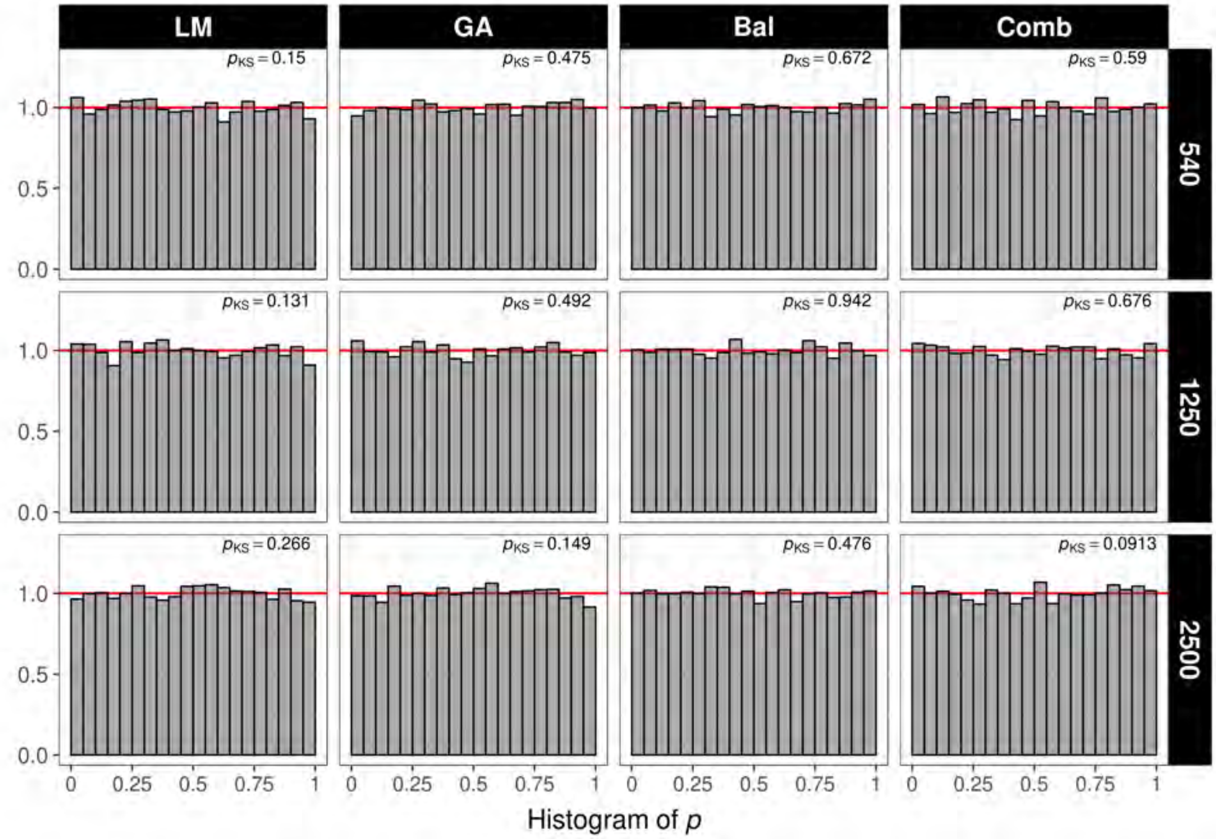

Figure S13: $P$-value histograms under the null hypothesis and likelihood ratio (LR) test in the comparison against heuristic designs across phase 2 sample sizes ($n = 540, 1250, 2500$). Column facets correspond to each of the studied designs. Row facets correspond to each phase 2 sample size. The top right values in each facet denote the $p$-value of the Kolmogorov-Sminrnov test that the observed $p$-values follow the expected uniform distribution. For LM and GA, $\mathbb{J}(\Phi)^{-1}$ was used in the optimization criterion.

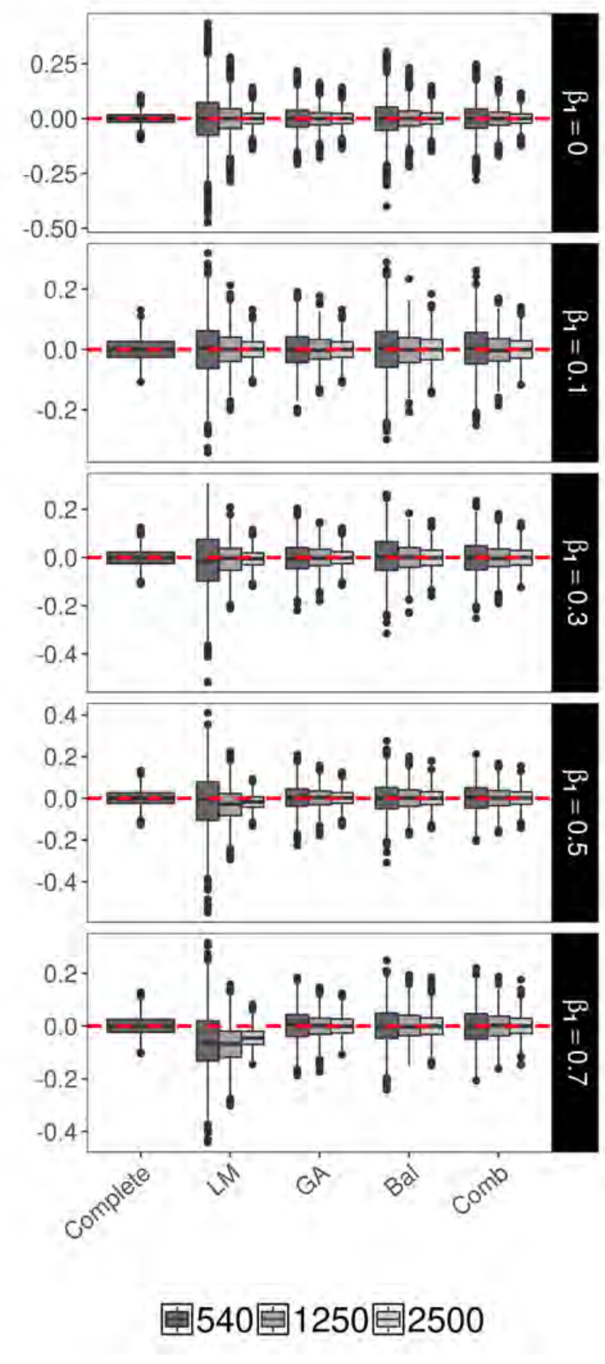

Figure S14: Boxplots for the distribution of the bias across genetic effect estimates ($\hat{\beta}_1 - \beta_1$) comparing the proposed designs against heuristic designs under a parameter-specific criterion. Row facets denote different true $\beta_1$ values $(0, 0.1, 0.3, 0.5, 0.7)$. For LM and GA, $\mathbb{J}(\Phi)^{-1}$ was used in the optimization criterion.

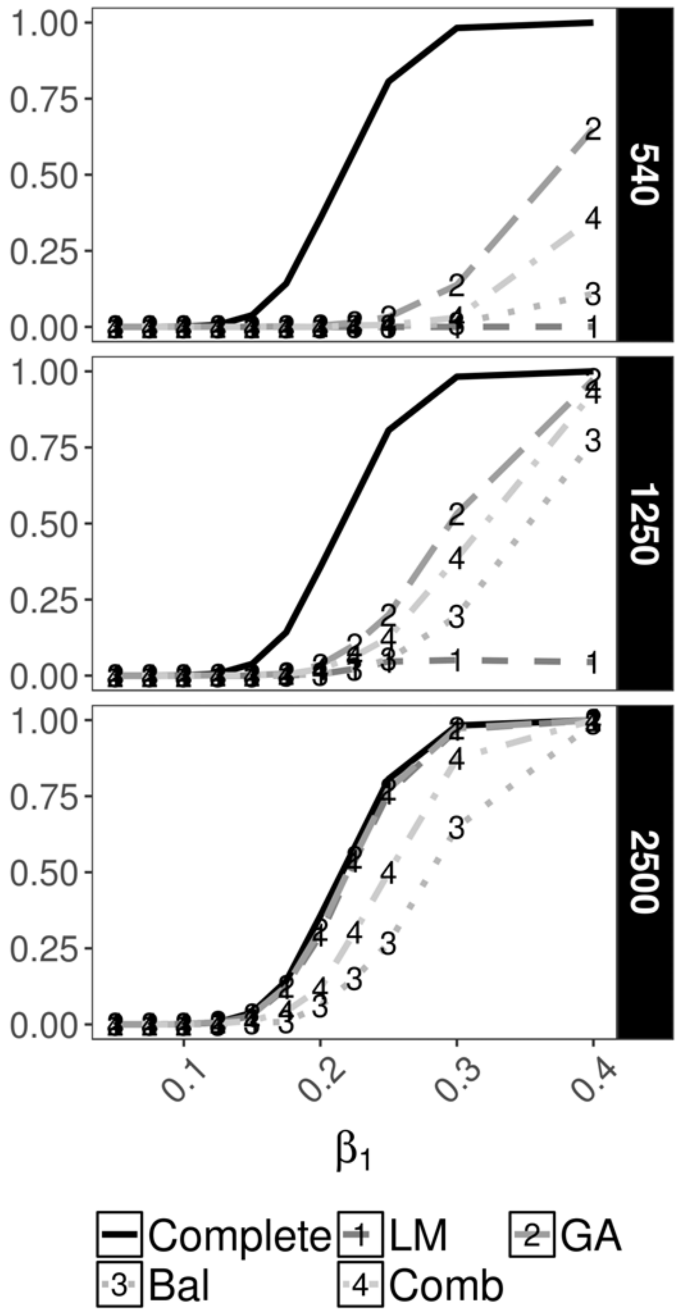

Figure S15: Power curves under the alternative $\beta_1 \neq 0$ at $\alpha = 1 \times 10^{-8}$ for testing $\mathrm{H}_0 : \beta_1 = 0$ under the LR test and parameter-specific criterion comparing designs (LM,GA,Bal,Comb) across phase 2 sample sizes. Row facets denote different phase 2 sample sizes ($n = 540, 1250, 2500$). For LM and GA, $\mathbb{J}(\Phi)^{-1}$ was used in the optimization criterion.

#### S4 Additional simulation studies

For additional simulations S4.1 and S4.2 below, we once gain focus on the parameter-specific optimality criterion as it may be preferred in the post-GWAS setting where single variant association is of main interest.

##### S4.1 Phase 1 & 2 sample sizes

We set effects sizes of $\beta_1 = \langle 0.150 + 0.025j | j = 0, \ldots, 4\rangle$, which led to empirical power in the range of 10-90% in the first set of simulations. Thus, for $N = 10000$, we consider values for $n$ that correspond to 0.10, 0.25 and 0.50 of the phase 1 data ($n/N$), i.e. $n = 1000, 2500, 5000$. The rest of the simulation parameters remain as in the main simulation study except for the number of replicates for which we draw 2000 this time. T1E rates at $\alpha$ =1% appear well controlled for the comparison against ranked designs and for the majority of designs in the comparison against heuristic designs (Table S12). The only exception in the comparison against heuristic designs is LM for which liberal T1E rates are observed under the Wald test and conservative especially when $n = 1000, 2500$; T1E rates under the LR test appear well controlled, however (Table S12). A closer inspection of the $p$-value distribution shows no gross departures for the expected uniform distribution and across values of $n$ under the LR test (Figures S16 and S17). As with the main simulation, GA achieves highest rEP when compared against heuristic designs across values of $n$; additionally, LM only demonstrates competitive power when $n = 5000$ ($n/N = .50$) (Table S13 and Figures S23-S24). Similarly, when comparing against ranked designs, TZL achieves the largest rEP across all values of $n$ with GA second and RDS, third. LM once again only shows comparable power when $n = 5000$ ($n/N = .50$) (Table S13 and Figures S23-S24).

##### S4.2 Joint distribution of $G$ and $Z$

We evaluate different combinations of MAFs ($q_G$ and $q_Z$) and $r$ values. LD, as quantified through $r$, corresponds to the correlation coefficient between $G$ and $Z$, which influences the informativeness of the auxiliary covariate $Z$. Intuitively, a poorly-correlated auxiliary covariate provides less information for $G$ than a highly correlated one. Consequently, for this study, we evaluate values of $r = -0.25, 0.25$ with combinations of $(q_G, q_Z)$ equal to $\{$ $(0.2, 0.2)$; $(0.2, 0.3)$; $(0.3, 0.2)$; $(0.3, 0.3)$ $\}$. The chosen values of $r$ represent a low correlation setting, especially compared to the main simulation setting ($r = 0.75$). For this set of simulations, we let $N = 5000$ and $n = 2500$, keeping the proportion of phase 1 data at 50%. Lastly, we focus on genetic effect sizes $\beta_1 = \langle 0.20 + 0.025j | j = 0, 1, 2\rangle$ and evaluate 1000 replicates. It is worth noting that these values for $r$ do not necessarily translate to lower LD between $G$ and $Z$. In fact, looking at $D'$, an alternative LD measure that takes into account the MAF in loci with domain in $[-1, 1]$ (although often denoted simply by its absolute value), the studied MAFs and $r$ combinations render $D'$ values ranging between $-1.00$ to $0.33$. Table S14 shows the exact correspondence between the studied combinations of MAFs and $r$ values with $D'$. When $r = -0.25$, $D'$ moves closer to 0 as the MAFs for $G$ and $Z$ increase. Likewise, when $r = 0.25$, $D'$ grows larger when MAFs are unequal. In absolute terms, LD (measured by $D'$) is stronger in all MAF instances when $r = -0.25$.

Type 1 error under the LR test remain at adequate levels across designs (Table S14). This is supported by the $p$-value distributions, which follow the expected uniform distribution (Figures S19-S22). Regarding rEP, similar patterns are observed with GA being most powerful when compared against heuristic designs with the combined designs closely following in second. On the other hand, GA, RDS, and TZL achieve similar rEP across correlation settings (Table S15 and Figures S23-S24). Interestingly, LM appears to be the design most affected by the different correlation patterns, with the lowest rEP for both comparisons against heuristic and ranked designs (Table S15 and Figures S23-S24).

#### S4.3 Results

##### S4.3.1 Tables

Table S12: Type 1 error rates at $\alpha = 1\%$ for simulation study S4.1 on phase 1 and 2 sample sizes ($N$ and $n$ respectively) across Wald, likelihood ratio (LR), and score tests under the parameter-specific optimality criterion. Each entry represents 10000 replicates. Phase 1 sample size is $N = 10$K whereas phase 2 sample size is $n = 1000, 2500, 5000$. The rest of the simulation parameters correspond to $q_G = 0.2$, $q_Z = 0.3$, $r = 0.75$, $\beta_0 = 2$, and $\sigma^2 = 1$. The type 1 error for the complete data is 1.11 for all tests.

| Test | $n$ | $n/N(\%)$ | vs. heuristic | | | | vs. ranked | | | |
|---|---|---|---|---|---|---|---|---|---|---|
| | | | LM | GA | Bal | Comb | LM | GA | RDS | TZL |
| Wald | 1000 | 10 | 16.64 | 1.09 | 1.89 | 1.19 | 1.17 | 0.95 | 0.99 | 1.05 |
| | 2500 | 25 | 4.08 | 1.06 | 1.06 | 0.91 | 0.94 | 0.96 | 0.90 | 1.10 |
| | 5000 | 50 | 1.04 | 0.98 | 1.19 | 1.08 | 0.95 | 0.95 | 1.10 | 0.98 |
| LR | 1000 | 10 | 1.07 | 0.93 | 1.26 | 0.90 | 0.89 | 0.90 | 0.98 | 1.03 |
| | 2500 | 25 | 0.83 | 0.99 | 0.90 | 0.87 | 0.91 | 0.96 | 0.90 | 1.09 |
| | 5000 | 50 | 1.04 | 0.96 | 1.15 | 1.08 | 0.94 | 0.95 | 1.10 | 0.97 |
| Score | 1000 | 10 | 0.01 | 0.93 | 1.19 | 0.87 | 0.86 | 0.84 | 0.94 | 1.03 |
| | 2500 | 25 | 0.17 | 0.98 | 0.87 | 0.84 | 0.91 | 0.96 | 0.90 | 1.09 |
| | 5000 | 50 | 1.02 | 0.97 | 1.10 | 1.05 | 0.94 | 0.95 | 1.10 | 0.97 |

Table S13: Relative empirical power (rEP), calculated as the ratio of the empirical power of each studied design over that of the complete data, across different genetic effects ($\beta_1$) and phase 2 sample sizes under the LR test ($\alpha = 5 \times 10^{-8}$) and parameter-specific optimality criterion in simulation study S4.1. Phase 1 sample size is 10K whereas phase 2 sample size is $n = 1000, 2500, 5000$ ($n/N = .10, .25, .50$). The rest of the simulation parameters correspond to $q_G = 0.2$, $q_Z = 0.3$, $r = 0.75$, $\beta_0 = 2$, and $\sigma^2 = 1$.

| $n$ | $\beta_1$ | vs. heuristic | | | | vs. ranked | | | |
|---|---|---|---|---|---|---|---|---|---|
| | | LM | GA | Bal | Comb | LM | GA | RDS | TZL |
| 1000 | 0.150 | 0.0 | 1.3 | 0.0 | 0.7 | 3.2 | 7.4 | 6.8 | 27.4 |
| | 0.175 | 0.0 | 2.4 | 0.3 | 0.6 | 6.3 | 16.6 | 15.5 | 46.2 |
| | 0.200 | 0.0 | 6.6 | 0.5 | 3.2 | 14.0 | 33.6 | 30.9 | 73.2 |
| | 0.225 | 0.1 | 15.4 | 1.4 | 7.4 | 28.6 | 56.5 | 53.9 | 89.8 |
| | 0.250 | 0.1 | 30.9 | 3.7 | 15.7 | 51.7 | 78.6 | 77.0 | 97.5 |
| 2500 | 0.150 | 3.8 | 14.0 | 3.8 | 12.3 | 32.2 | 44.7 | 44.5 | 80.8 |
| | 0.175 | 8.2 | 26.0 | 8.1 | 21.2 | 50.8 | 60.7 | 60.0 | 88.7 |
| | 0.200 | 18.9 | 46.5 | 18.1 | 40.3 | 74.5 | 84.9 | 84.5 | 97.3 |
| | 0.225 | 32.1 | 71.3 | 37.3 | 63.6 | 92.0 | 95.3 | 95.1 | 99.6 |
| | 0.250 | 29.0 | 88.9 | 59.6 | 84.4 | 98.0 | 99.1 | 99.1 | 100.0 |
| 5000 | 0.150 | 88.7 | 91.3 | 22.6 | 48.2 | 94.5 | 94.6 | 86.0 | 95.5 |
| | 0.175 | 95.5 | 95.2 | 40.8 | 67.2 | 96.5 | 96.4 | 91.8 | 98.0 |
| | 0.200 | 98.8 | 99.2 | 63.9 | 87.2 | 99.3 | 99.4 | 98.3 | 99.4 |
| | 0.225 | 99.8 | 99.9 | 85.6 | 96.9 | 99.9 | 100.0 | 99.8 | 100.0 |
| | 0.250 | 100.0 | 100.0 | 96.0 | 99.5 | 100.0 | 100.0 | 100.0 | 100.0 |

Table S14: Type 1 error rates under the LR test ($\alpha = 1\%$) for simulation study S4.2 on the joint distribution of $G$ and $Z$ under the parameter-specific optimality criterion. Each entry represents 3000 replicates. The rest of the simulation parameters correspond to $N = 5000$, $n = 2500$, $\beta_0 = 2$, and $\sigma^2 = 1$.

| $r$ | $q_G$ | $q_Z$ | D′ | Complete | vs. heuristic | | | | vs. ranked | | | |
|---|---|---|---|---|---|---|---|---|---|---|---|---|
| | | | | | LM | GA | Bal | Comb | LM | GA | RDS | TZL |
| -0.25 | 0.2 | 0.2 | -1.00 | 0.93 | 1.17 | 0.73 | 0.67 | 1.03 | 0.87 | 0.80 | 0.83 | 1.03 |
| | 0.2 | 0.3 | -0.76 | 0.93 | 1.07 | 1.13 | 1.10 | 1.10 | 1.27 | 1.03 | 1.07 | 1.07 |
| | 0.3 | 0.2 | -0.76 | 0.93 | 1.30 | 0.83 | 0.83 | 1.23 | 0.83 | 1.03 | 1.07 | 1.07 |
| | 0.3 | 0.3 | -0.58 | 0.87 | 1.40 | 0.97 | 0.83 | 0.93 | 0.67 | 0.77 | 0.80 | 0.80 |
| 0.25 | 0.2 | 0.2 | 0.25 | 1.03 | 0.97 | 0.97 | 1.23 | 0.83 | 1.20 | 1.37 | 1.37 | 1.20 |
| | 0.2 | 0.3 | 0.33 | 0.83 | 0.80 | 0.93 | 1.03 | 0.67 | 0.70 | 0.87 | 0.93 | 0.97 |
| | 0.3 | 0.2 | 0.33 | 0.90 | 1.50 | 1.00 | 1.23 | 1.13 | 1.23 | 1.27 | 1.30 | 1.27 |
| | 0.3 | 0.3 | 0.25 | 0.80 | 0.90 | 1.17 | 0.77 | 0.93 | 0.90 | 0.87 | 0.90 | 0.93 |

Table S15: Relative empirical power (rEP), calculated as the ratio of the empirical power of each studied design over that of the complete data, across different effect sizes under the LR test ($\alpha = 5 \times 10^{-8}$) and parameter-specific optimality criterion on the joint distribution of $G$ and $Z$ in simulation study S4.2. The rest of the simulation parameters correspond to $N = 5000$, $n = 2500$, $\beta_0 = 2$, and $\sigma^2 = 1$.

| $r$ | $q_G$ | $q_Z$ | $\beta_1$ | vs. heuristic | | | | vs. ranked | | | |
|---|---|---|---|---|---|---|---|---|---|---|---|
| | | | | LM | GA | Bal | Comb | LM | GA | RDS | TZL |
| -0.25 | 0.2 | 0.2 | 0.200 | 16.9 | 88.4 | 29.6 | 65.7 | 91.9 | 98.7 | 98.7 | 99.3 |
| | | | 0.225 | 25.6 | 96.7 | 51.8 | 87.8 | 98.6 | 99.9 | 99.9 | 99.9 |
| | | | 0.250 | 36.5 | 99.8 | 71.1 | 96.1 | 100.0 | 100.0 | 100.0 | 100.0 |
| | 0.2 | 0.3 | 0.200 | 32.5 | 85.5 | 27.7 | 68.6 | 82.6 | 98.8 | 98.9 | 98.7 |
| | | | 0.225 | 53.6 | 98.5 | 55.7 | 92.2 | 96.5 | 100.1 | 100.1 | 100.1 |
| | | | 0.250 | 63.3 | 99.6 | 74.1 | 97.9 | 99.3 | 100.0 | 100.0 | 100.0 |
| | 0.3 | 0.2 | 0.200 | 19.8 | 97.2 | 61.9 | 91.0 | 97.6 | 100.0 | 100.0 | 100.0 |
| | | | 0.225 | 37.0 | 99.6 | 83.9 | 98.7 | 99.9 | 100.0 | 100.0 | 100.0 |
| | | | 0.250 | 60.1 | 100.0 | 95.2 | 99.8 | 100.0 | 100.0 | 100.0 | 100.0 |
| | 0.3 | 0.3 | 0.200 | 15.8 | 96.9 | 64.8 | 93.2 | 94.1 | 99.8 | 99.8 | 99.8 |
| | | | 0.225 | 32.3 | 100.0 | 85.4 | 99.2 | 99.6 | 100.0 | 100.0 | 100.0 |
| | | | 0.250 | 55.6 | 100.0 | 96.2 | 99.9 | 99.9 | 100.0 | 100.0 | 100.0 |
| 0.25 | 0.2 | 0.2 | 0.200 | 17.7 | 87.9 | 50.8 | 83.6 | 85.0 | 99.1 | 99.1 | 98.7 |
| | | | 0.225 | 40.0 | 97.8 | 74.9 | 95.8 | 96.4 | 99.6 | 99.6 | 99.7 |
| | | | 0.250 | 69.2 | 99.6 | 91.6 | 99.4 | 99.4 | 100.0 | 100.0 | 100.0 |
| | 0.2 | 0.3 | 0.200 | 14.4 | 87.8 | 49.1 | 81.6 | 86.2 | 98.9 | 98.6 | 98.9 |
| | | | 0.225 | 39.9 | 96.3 | 73.2 | 94.4 | 96.4 | 100.0 | 100.0 | 99.9 |
| | | | 0.250 | 71.8 | 99.8 | 90.5 | 99.6 | 99.7 | 100.0 | 100.0 | 100.0 |
| | 0.3 | 0.2 | 0.200 | 12.7 | 97.9 | 71.9 | 95.1 | 95.0 | 100.0 | 100.0 | 100.1 |
| | | | 0.225 | 30.9 | 99.8 | 92.1 | 99.7 | 99.9 | 100.0 | 100.0 | 100.0 |
| | | | 0.250 | 54.4 | 100.0 | 98.8 | 100.0 | 100.0 | 100.0 | 100.0 | 100.0 |
| | 0.3 | 0.3 | 0.200 | 19.0 | 97.4 | 73.0 | 95.5 | 96.8 | 99.8 | 99.8 | 99.8 |
| | | | 0.225 | 43.8 | 99.9 | 91.4 | 99.4 | 99.6 | 100.0 | 100.0 | 100.0 |
| | | | 0.250 | 70.4 | 100.0 | 98.2 | 100.0 | 100.0 | 100.0 | 100.0 | 100.0 |

##### S4.3.2 Figures

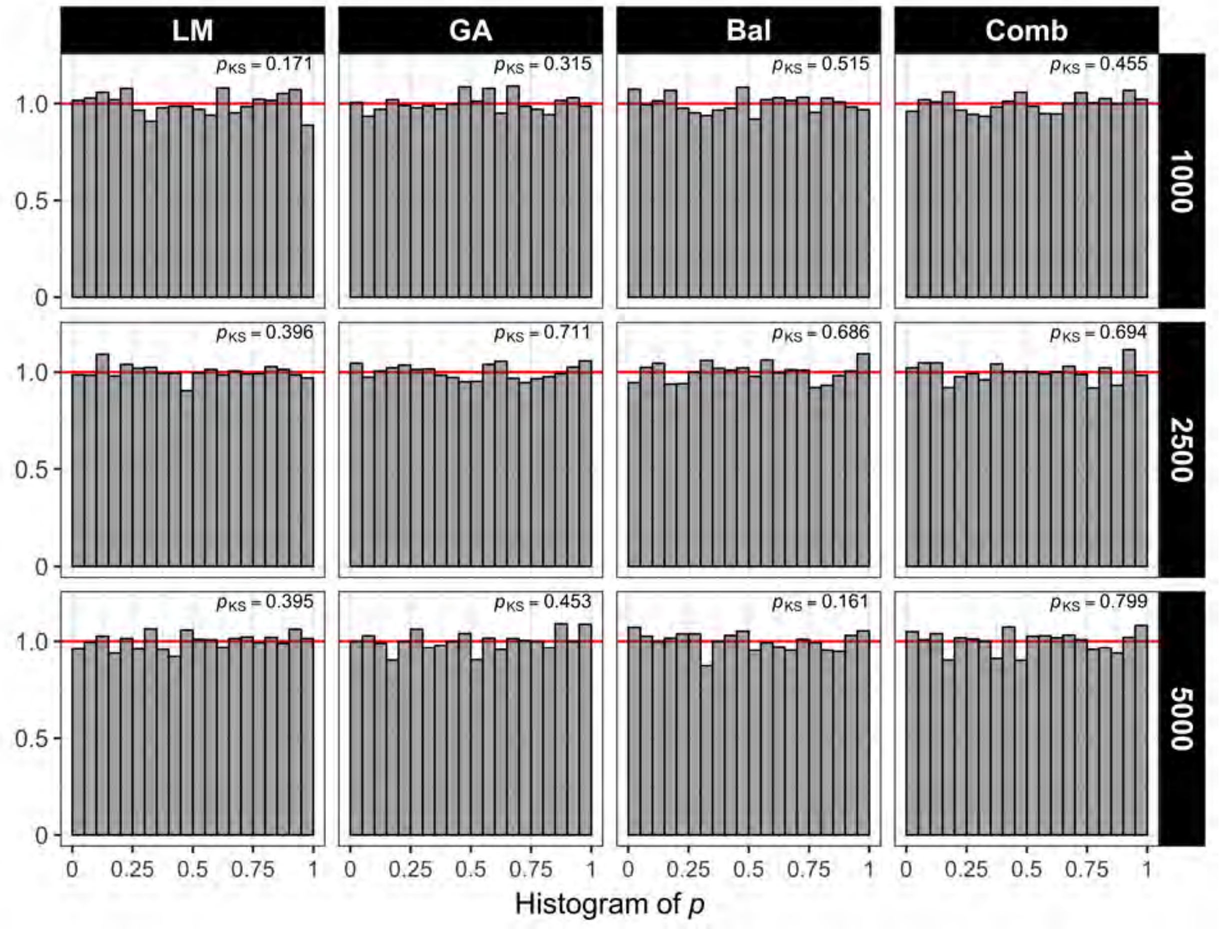

Figure S16: $P$-value histograms under the null hypothesis and LR test across studied designs in simulation study S4.1 for sample sizes in phases 1 & 2 when comparing against heurstic designs. Column facets correspond to each of the studied designs. Row facets correspond to different phase 2 sample sizes $n = 1000, 2500, 5000$ ($n/N = 0.10, 0.25, 0.75$). The top right values in each facet denote the $p$-value of the Kolmogorov-Smirnov test that the observed $p$-values follow the expected uniform distribution.

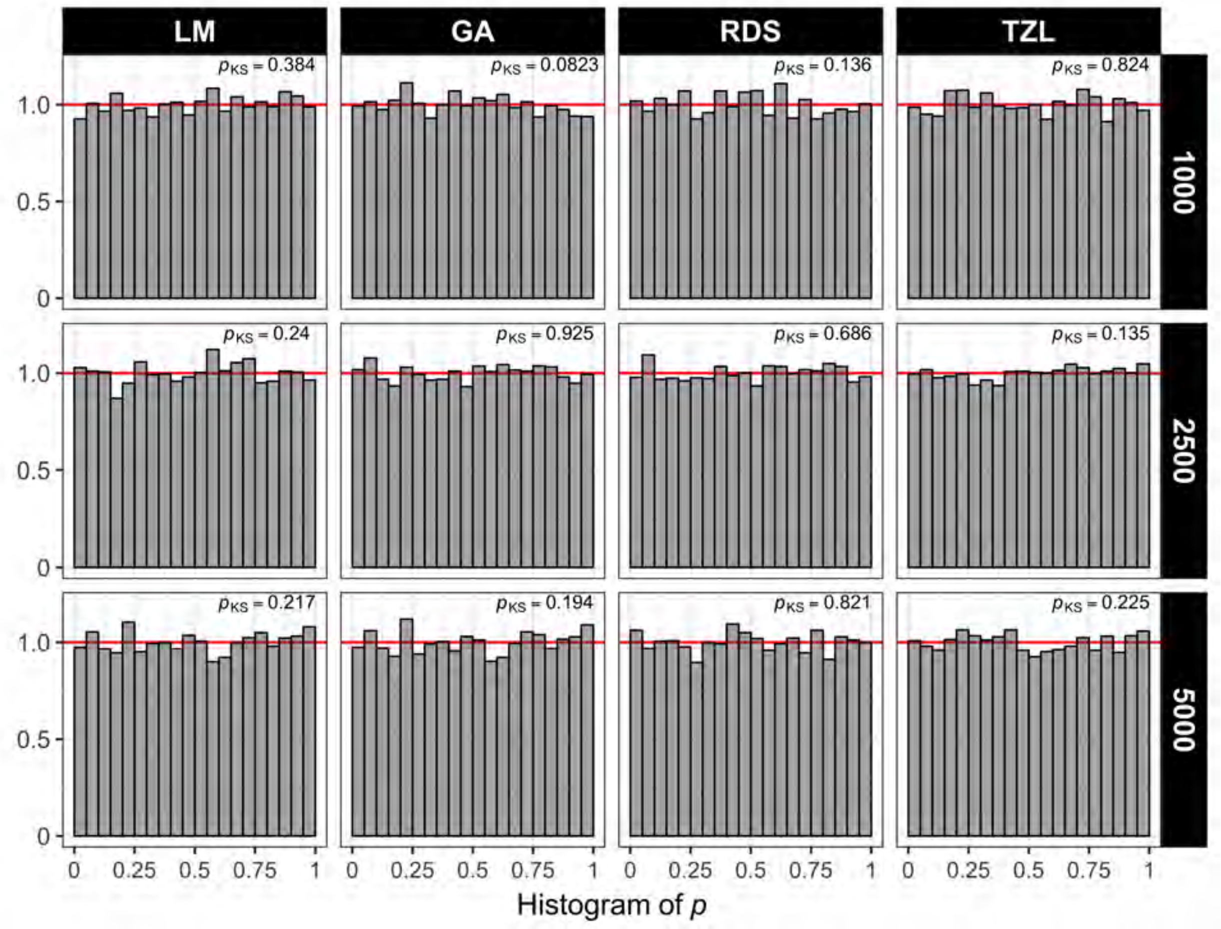

Figure S17: $P$-value histograms under the null hypothesis and LR test across studied designs in simulation study S4.1 for sample sizes in phases 1 & 2, $N = 10000$, when comparing against ranked designs. Column facets correspond to each of the studied designs. Row facets correspond to different phase 2 sample sizes $n = 1000, 2500, 5000$ ($n/N = 0.10, 0.25, 0.75$). The top right values in each facet denote the $p$-value of the Kolmogorov-Smirnov test that the observed $p$-values follow the expected uniform distribution.

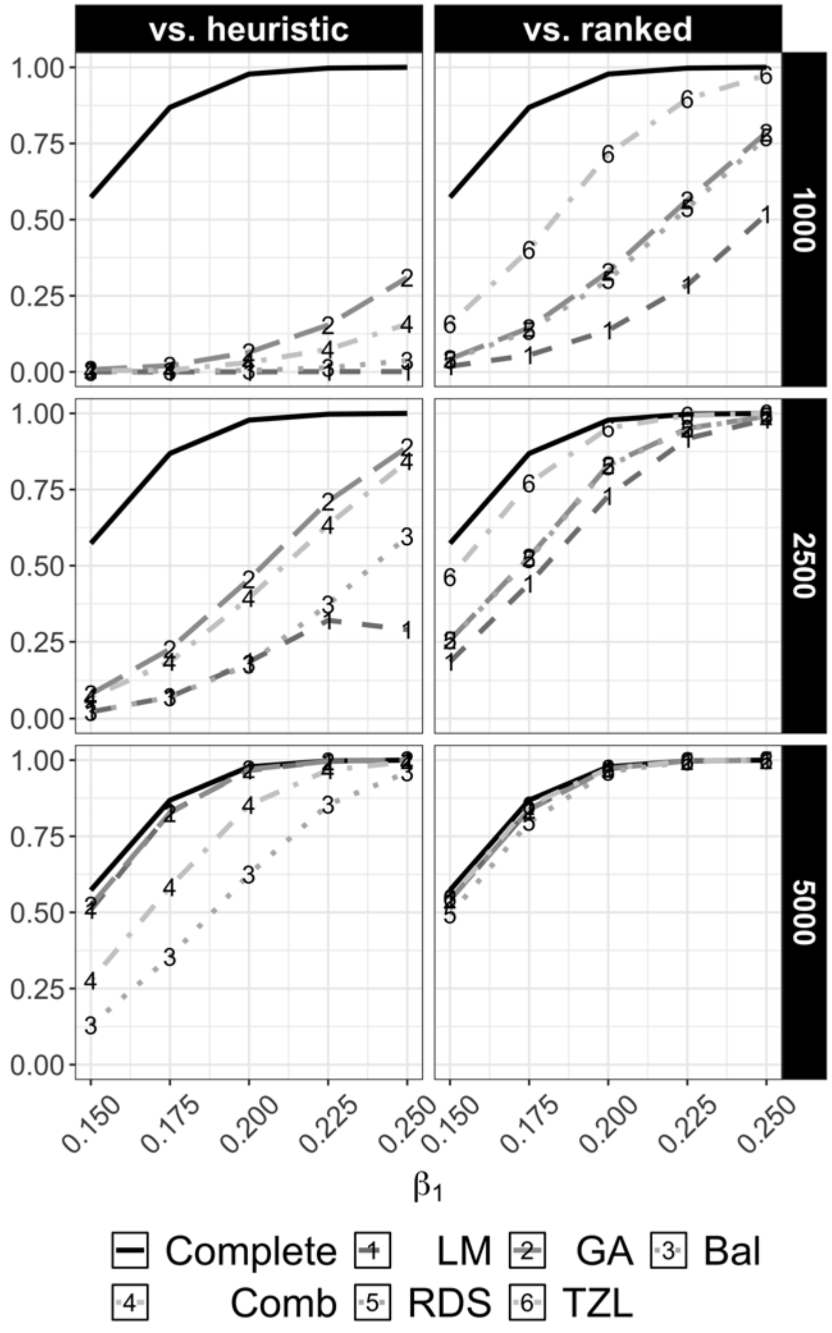

Figure S18: Power curves under the alternative $\beta_1 \neq 0$ at $\alpha = 5 \times 10^{-8}$ for testing $\mathrm{H}_0 : \beta_1 = 0$ under the LR test across studied designs in simulation study S4.1 for sample sizes in phases 1 & 2, $N = 10000$. Row facets denote phase 2 samples sizes ($n = 1000, 2500, 5000$) whereas column facets denote the design comparison (vs heuristic or vs. ranked, respectively).

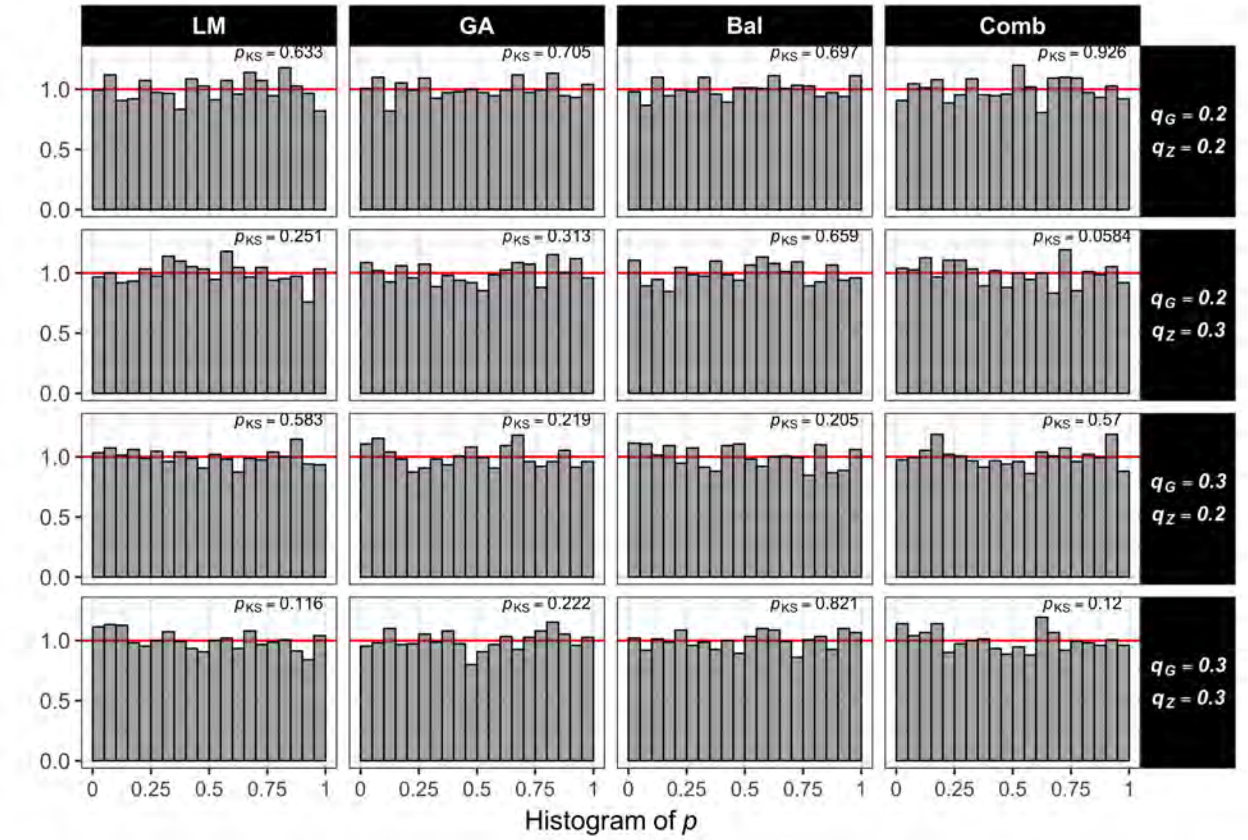

Figure S19: $P$-value histograms under the null hypothesis and LR test across studied designs in simulation study S4.2 on the joint distribution of $G$ and $Z$ when $r = -0.25$ and comparing against heuristic designs. Column facets correspond to each of the studied designs. Row facets correspond to four different minor allele frequency combinations for $G$ and $Z$. The top right values in each facet denote the $p$-value of the Kolmogorov-Smirnov test that the observed $p$-values follow the expected uniform distribution.

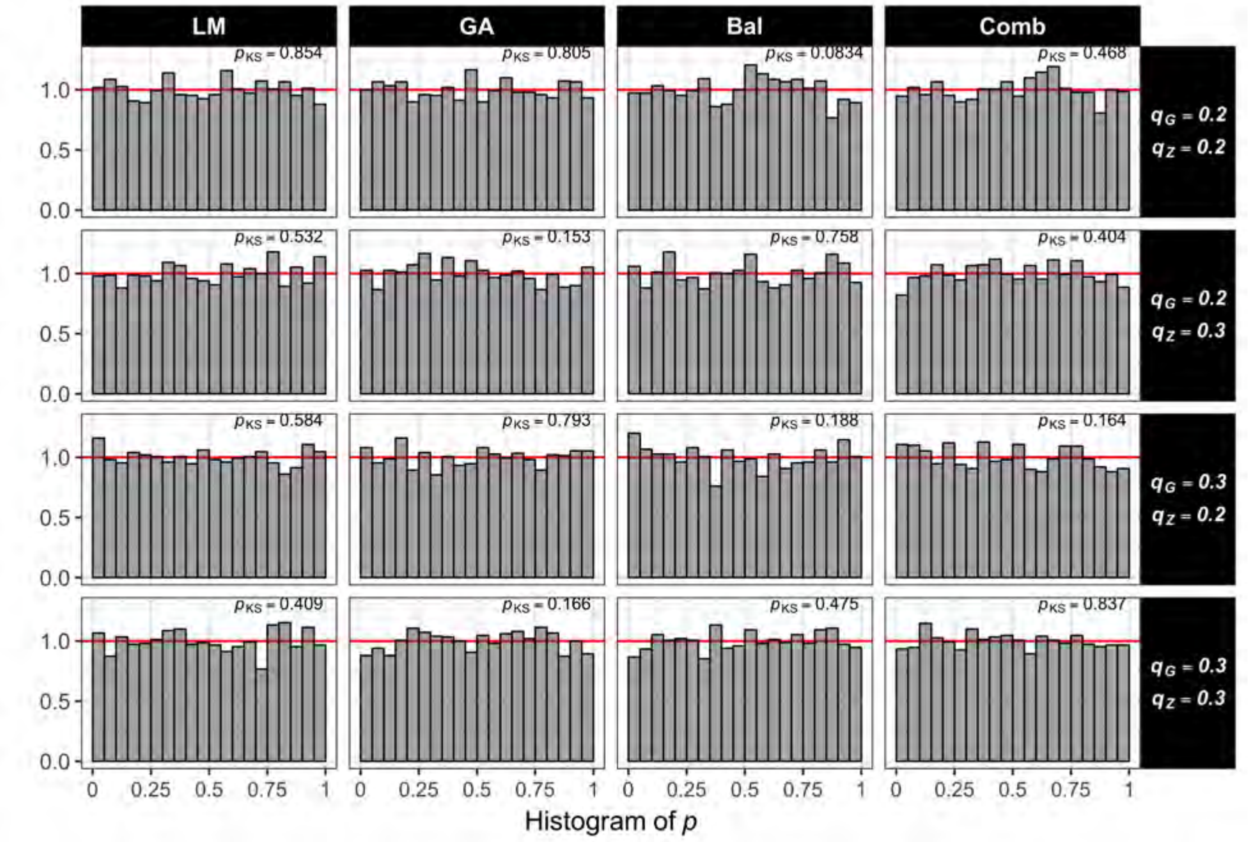

Figure S20: $P$-value histograms under the null hypothesis and LR test across studied designs in simulation study S4.2 on the joint distribution of $G$ and $Z$ when $r = 0.25$ and comparing against heuristic designs. Column facets correspond to each of the studied designs. Row facets correspond to four different minor allele frequency combinations for $G$ and $Z$. The top right values in each facet denote the $p$-value of the Kolmogorov-Smirnov test that the observed $p$-values follow the expected uniform distribution.

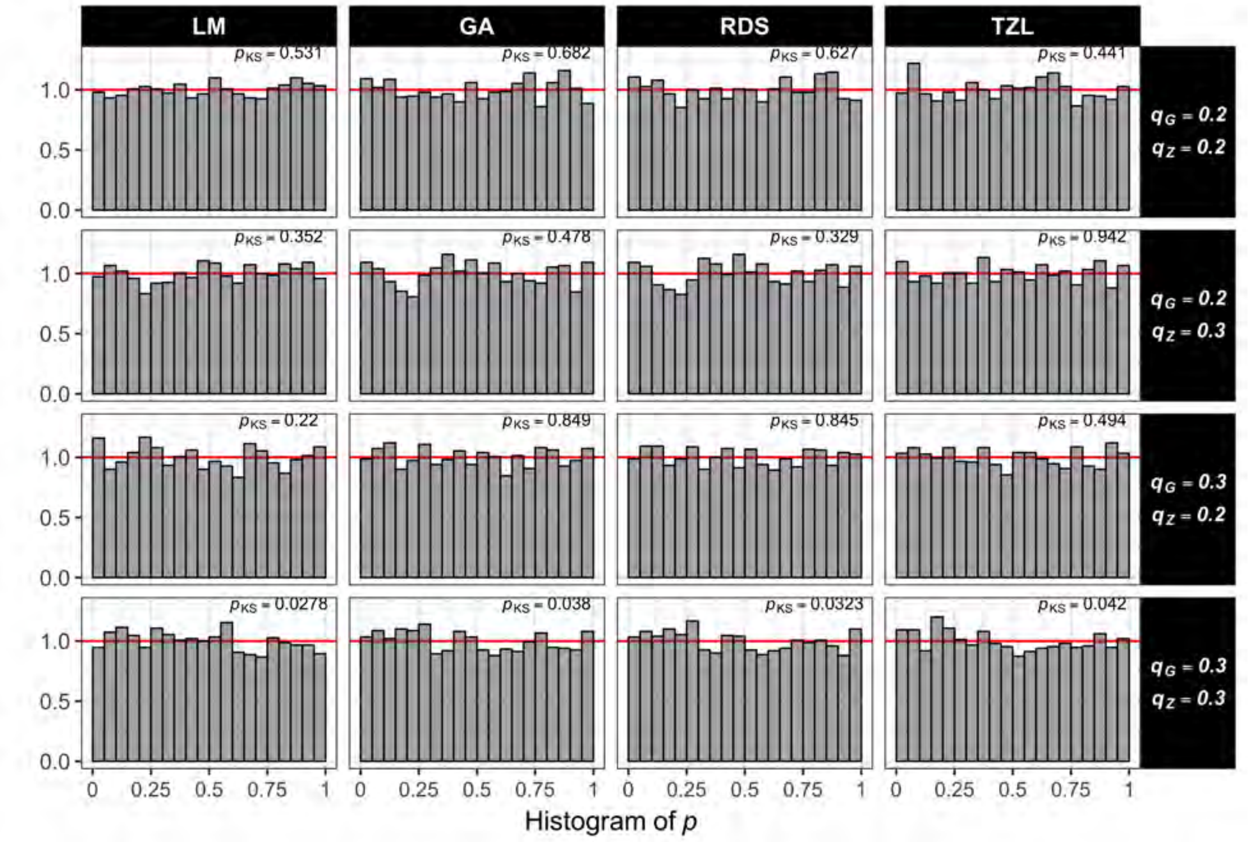

Figure S21: $P$-value histograms under the null hypothesis and LR test across studied designs in simulation study S4.2 on the joint distribution of $G$ and $Z$ when $r = -0.25$ and comparing against ranked designs. Column facets correspond to each of the studied designs. Row facets correspond to four different minor allele frequency combinations for $G$ and $Z$. The top right values in each facet denote the $p$-value of the Kolmogorov-Smirnov test that the observed $p$-values follow the expected uniform distribution.

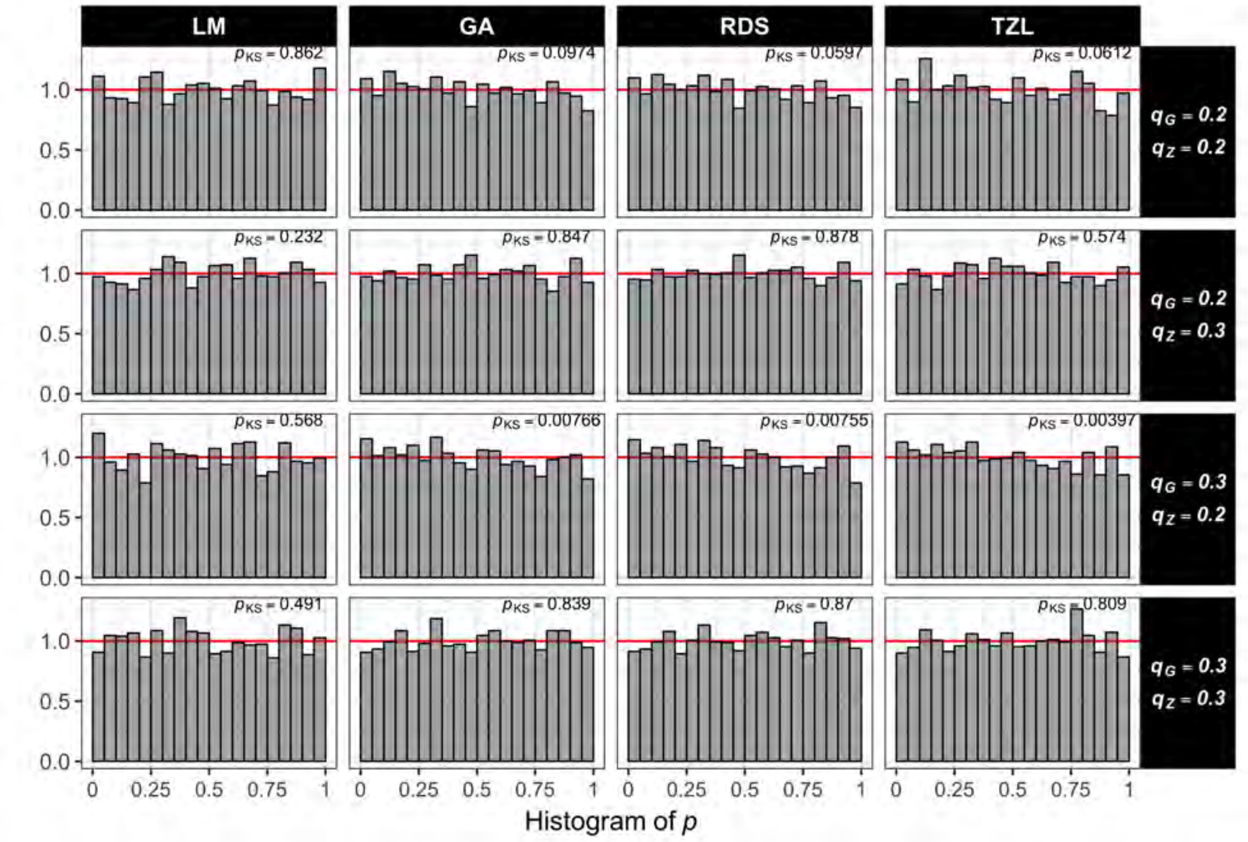

Figure S22: $P$-value histograms under the null hypothesis and LR test across studied designs in simulation study S4.2 on the joint distribution of $G$ and $Z$ when $r = 0.25$ and comparing against ranked designs. Column facets correspond to each of the studied designs. Row facets correspond to four different minor allele frequency combinations for $G$ and $Z$. The top right values in each facet denote the $p$-value of the Kolmogorov-Smirnov test that the observed $p$-values follow the expected uniform distribution.

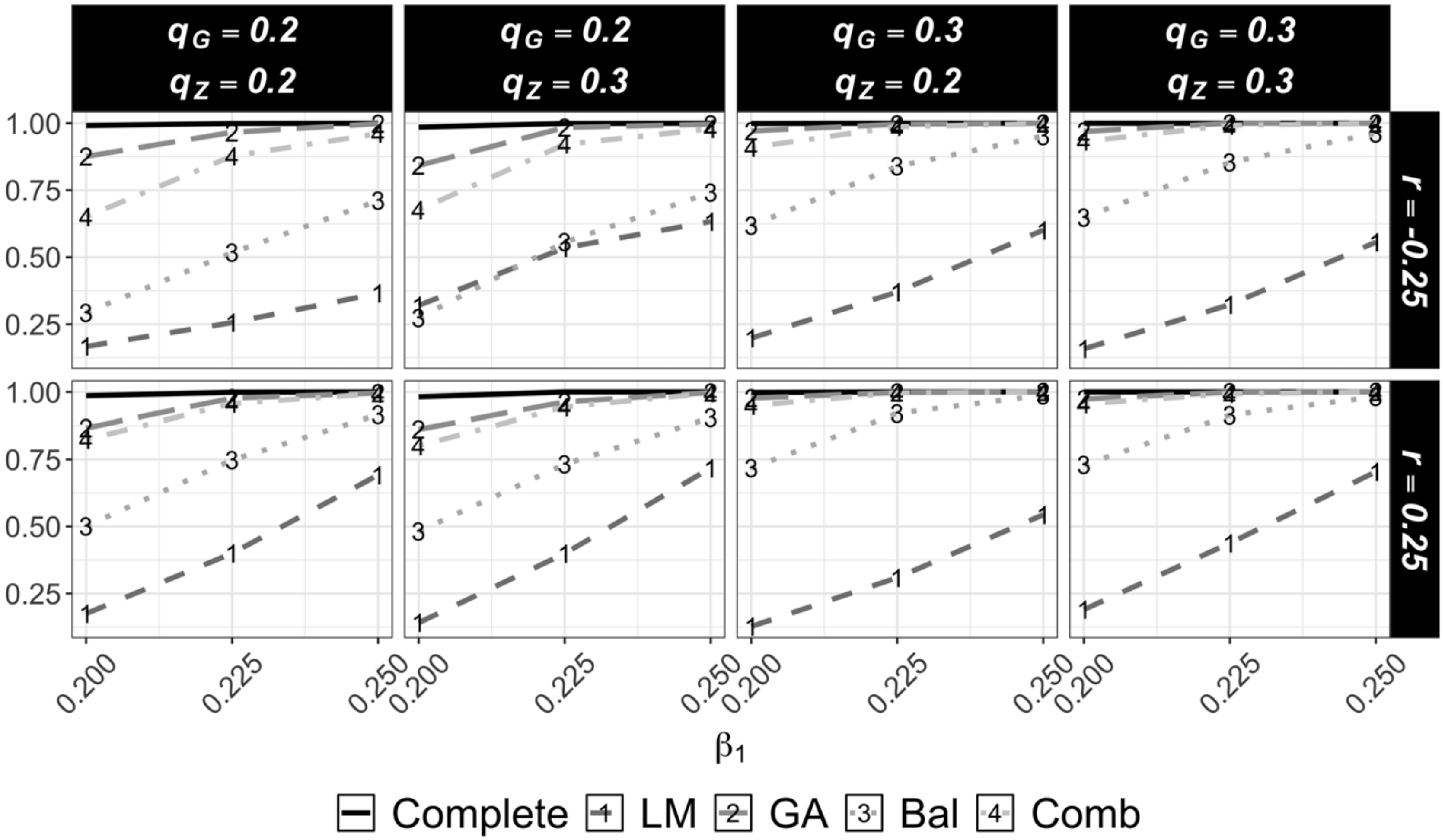

Figure S23: Power curves under the alternative $\beta_1 \neq 0$ at $\alpha = 5 \times 10^{-8}$ for testing $\mathrm{H}_0 : \beta_1 = 0$ under the LR test across studied designs in simulation study S4.2 for the joint distribution of $G$ and $Z$ when comparing against heuristic designs. Row facets denote the level of linkage correlation between $G$ and $Z$ measured through $r$ $(-0.25, 0.25)$. Column facets denote four different minor allele frequency combinations for $G$ and $Z$.

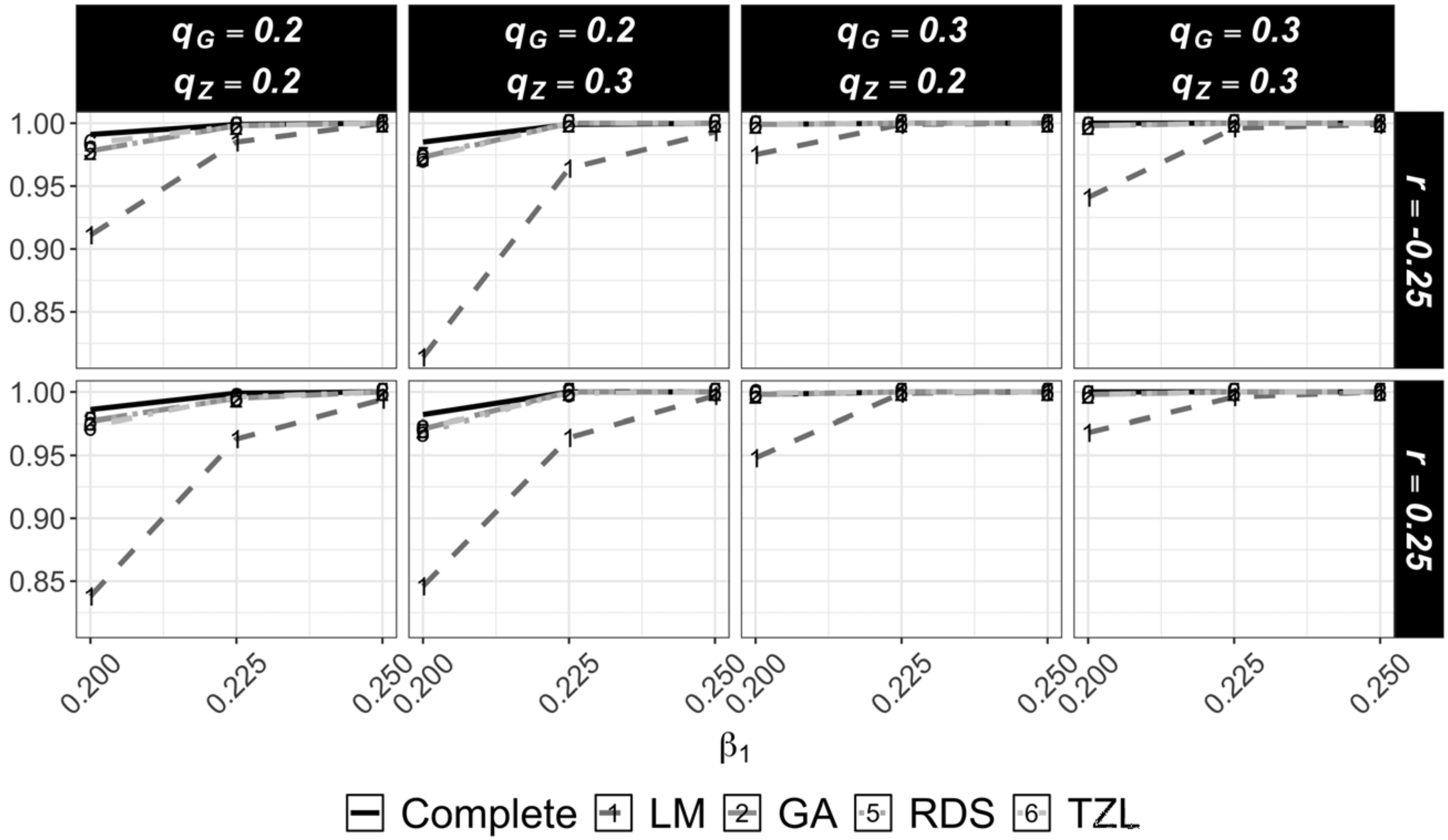

Figure S24: Power curves under the alternative $\beta_1 \neq 0$ at $\alpha = 5 \times 10^{-8}$ for testing $\mathrm{H}_0 : \beta_1 = 0$ under the score test across studied designs in simulation study S4.2 for the joint distribution of $G$ and $Z$ when comparing against ranked designs. Row facets denote the level of linkage correlation between $G$ and $Z$ measured through $r$ $(-0.25, 0.25)$. Column facets denote four different minor allele frequency combinations for $G$ and $Z$.

### S5 Data generation details under realistic LD patterns

Here, we provide details on the data generation outlined in Section 4.2. As mentioned, we simulate genetic variants from a pre-specified region for the European population in the 1000 Genomes Project using the R package *sim1000G* [2]. Here, we describe the data generation procedure

1. Designate rs247617 (position 56990716, $q_Z = 0.293$) as the GWAS SNP, which has been associated with several cardiovascular traits in the GWAS catalog [3].

2. Fix genetic variants across simulation replicates from a region comprising genes HERPUD1 and CETP in chromosome 16 (29 sequence variants (seq-SNPs) within ±5000bp of rs247617 with a minimum MAF of 5% in the reference population).

3. Draw a dataset of 5000 independent subjects' genotypes. All variants in the selected region are coded additively with minor allele designated as the risk allele. The LD structure in terms of $\mathrm{r}^2$ and $\mathrm{D}'$ in the simulated region is depicted in Figure S25.

4. Select four loci as "causal" SNPs, $\boldsymbol{G} = (G_1, \ldots, G_4)$, with corresponding effect sizes $\boldsymbol{\beta}_1 = (\beta_{1G_1}, \ldots, \beta_{1G_4})'$ in positions 56989830, 56993324, 56994990, 56995236 (hereafter all positions are truncated to the last 5 digits). These variants are chosen to have different MAF and LD (with $Z$) combinations (Table S16).

5. In each of 500 replicates, generate a QT following $Y = \beta_0 + \boldsymbol{\beta}_1' \boldsymbol{G} + \varepsilon$, where $\beta_0 = 2$, $\boldsymbol{\beta}_1 = (-0.200, 0.125, 0.250, -0.150)'$, and $\varepsilon \sim \mathcal{N}(0, \sigma^2 = 1)$. This data generation introduces multiple causal variants. GWAS-SNP-association is induced by only keeping replicates that show association between $Z$ and $Y$ ($p < 1 \times 10^{-5}$) as in Section 3.1.

#### S5.1 Tables

Table S16: Characteristics of the four causal variants in practical simulation, the base pair positions (pos.) are truncated to the last 5 digits. Minor allele frequencies are denoted by $q_G$. LD measures are calculated with respect to the GWAS SNP, $Z$, rs247617 in pos. 56990716 (hg19).

| $\boldsymbol{G}$ | **pos.** | $\boldsymbol{q_G}$ | $\boldsymbol{r_{G,Z}}$ | $\boldsymbol{r^2_{G,Z}}$ | $\mathbf{D}'_{G,Z}$ |
|---|---|---|---|---|---|
| $G_1$ | 89830 | 0.478 | -0.414 | 0.172 | -0.673 |
| $G_2$ | 93324 | 0.294 | 0.942 | 0.888 | 0.945 |
| $G_3$ | 94990 | 0.126 | -0.147 | 0.021 | -0.600 |
| $G_4$ | 95236 | 0.486 | 0.540 | 0.292 | 0.816 |

#### S5.2 Figures

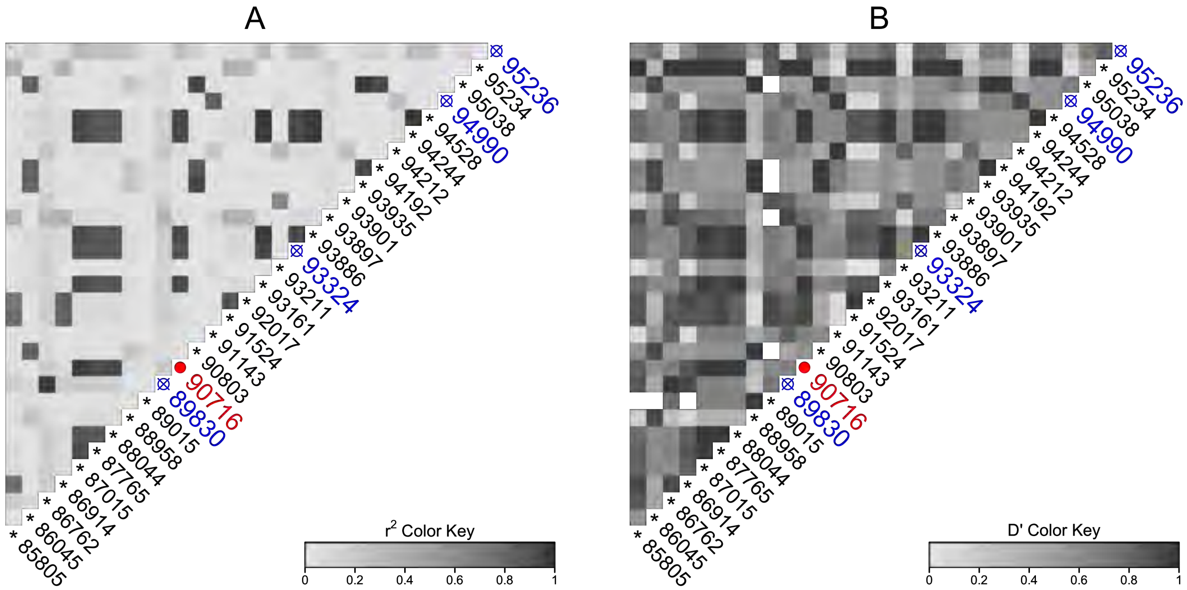

Figure S25: LD heatmap plots ($r^2$ and —D′— in panels A and B respectively) of the designated region in chromosome 16 between genes HERPUD1 and CETP for the fine-mapping simulation. Each square corresponds to the correlation ($r^2$ or —D′—) between any pair of variants in the region. Darker shades denote higher correlation. The variant marked with a filled circle denotes the GWAS SNP, $Z$ (rs247617, pos. 56990716) while the variants marked with the crossed circle denote the designated four causal variant.

#### S6 Conditional fine-mapping analysis

From the single-variant analysis, causal variant 89830 is the top SNP in 76% of the replicates for the complete data analysis and ranges between 50-72% for the studied designs across different phase 2 sample sizes (Table S17). Variant 86762, which is in high LD with 89830, is the next most frequent top SNP in the complete data (17% of the replicates) with frequencies ranging between 19-26% across designs (Table S17). Two other causal variants (94990 and 95236) were next to be ranked as top SNPs. Interestingly, hitchhikers and other non-causal variants were ranked as top SNPs in decreasing numbers as the phase 2 sample size increases (Table S17). To make results comparable, we run the conditional region scan by setting variant 89830 as $G_{top}$ for all replicates.

##### S6.1 Results

In the conditional analysis, we observe that the ranking in terms of the studied designs for $\beta_1$ remains as in the single-variant analysis with GA, RDS and TZL designs showing, on average, smaller $p$-values followed by LM. Additionally, conditional analysis shows that after adjusting for the top SNP, the effect for the majority of the hitchhiker variants is more accurately estimated except for variant 91143, which is in high LD with causal variant 94990 ($r = 0.913$, $D' = 0.987$) (Tables S18-S19). Similarly, the empirical power ($\alpha = 0.5/29$) and $p$-value distribution for the conditional analysis better reflects the true generating mechanism by boosting the signal in the two causal SNPs while decreasing it in the non-causal variants (Tables S20-S21 and bottom of Figures S26-S27) Conditional analysis results for $\beta_2$ (top SNP 89830) suggest that parameter estimation and power substantially improve compared to the single-variant analysis (Tables S22 and S23). Once again, we observe no major differences between GA and TZL across optimality criteria.

###### S6.1.1 Tables

Table S17: Top SNPs in complete data analysis and studied designs across 500 replicates in fine-mapping simulation single-variant analysis. Even though variant 86762$^\dagger$ is also a "hitchhiker", we separate its results given its high LD with causal variant 89830 ($r = 0.975$, $D' = 0.976$).

| $n$ | Criterion | Design | 86762$^\dagger$ | 89830$^*$ | 93324$^*$ | 94990$^*$ | 95236$^*$ | Other Hitchhikers | Other non-causal |
|---|---|---|---|---|---|---|---|---|---|
| | Complete | | 87 | 381 | 0 | 16 | 14 | 2 | 0 |
| 1250 | RDS | | 113 | 328 | 0 | 21 | 29 | 9 | 0 |
| | Par-spec | LM | 129 | 252 | 0 | 49 | 43 | 25 | 2 |
| | | GA | 112 | 330 | 0 | 23 | 27 | 8 | 0 |
| | | TZL | 119 | 321 | 0 | 22 | 31 | 7 | 0 |
| | A-opt | LM | 95 | 263 | 0 | 33 | 57 | 30 | 22 |
| | | GA | 112 | 323 | 0 | 23 | 33 | 9 | 0 |
| | | TZL | 115 | 325 | 0 | 23 | 29 | 8 | 0 |
| | D-opt | LM | 101 | 262 | 2 | 30 | 53 | 37 | 15 |
| | | GA | 113 | 327 | 0 | 21 | 30 | 9 | 0 |
| | | TZL | 113 | 319 | 0 | 27 | 33 | 8 | 0 |
| 2500 | RDS | | 99 | 359 | 0 | 18 | 22 | 2 | 0 |
| | Par-spec | LM | 113 | 333 | 0 | 27 | 20 | 7 | 0 |
| | | GA | 98 | 360 | 0 | 18 | 22 | 2 | 0 |
| | | TZL | 99 | 360 | 0 | 19 | 19 | 3 | 0 |
| | A-opt | LM | 109 | 324 | 0 | 27 | 30 | 10 | 0 |
| | | GA | 98 | 359 | 0 | 18 | 23 | 2 | 0 |
| | | TZL | 95 | 365 | 0 | 15 | 23 | 2 | 0 |
| | D-opt | LM | 103 | 335 | 0 | 23 | 32 | 7 | 0 |
| | | GA | 101 | 355 | 0 | 19 | 22 | 3 | 0 |
| | | TZL | 97 | 360 | 0 | 14 | 25 | 4 | 0 |

Table S18: Estimates and asymptotic standard errors for $\beta_1$ in fine-mapping simulation conditional analysis for $n = 1250$. This table removes results from variant 56989830 as it appeared as the top SNP in the majority of replicates, thus, it has too few replicates to be included. Positions marked with $*$ denote causal variants whereas, the ones marked with † denote hitchhikers. The remaining ones are non-causal. Positions are truncated to the last five digits.

| **$G$ pos.** | **$\beta_{1G}$** | **Complete** | **RDS** | **Par-spec** | | | **A-opt** | | | **D-opt** | | |
|---|---|---|---|---|---|---|---|---|---|---|---|---|
| | | | | **LM** | **GA** | **TZL** | **LM** | **GA** | **TZL** | **LM** | **GA** | **TZL** |
| 85805 | 0.00 | 0.021 (0.03) | 0.022 (0.04) | 0.010 (0.05) | 0.022 (0.04) | 0.021 (0.04) | 0.012 (0.05) | 0.022 (0.04) | 0.020 (0.04) | 0.016 (0.05) | 0.022 (0.04) | 0.023 (0.04) |
| 86045† | 0.00 | 0.034 (0.03) | 0.035 (0.03) | 0.039 (0.05) | 0.035 (0.03) | 0.035 (0.03) | 0.037 (0.05) | 0.035 (0.03) | 0.037 (0.03) | 0.036 (0.04) | 0.035 (0.03) | 0.034 (0.03) |
| 86762† | 0.00 | 0.000 (0.09) | 0.000 (0.11) | 0.014 (0.15) | 0.000 (0.11) | 0.002 (0.11) | 0.011 (0.15) | -0.001 (0.11) | 0.003 (0.11) | -0.008 (0.15) | 0.000 (0.11) | -0.003 (0.11) |
| 86914 | 0.00 | 0.023 (0.03) | 0.025 (0.04) | 0.015 (0.05) | 0.025 (0.04) | 0.024 (0.04) | 0.015 (0.05) | 0.025 (0.04) | 0.022 (0.04) | 0.018 (0.04) | 0.025 (0.04) | 0.025 (0.03) |
| 87015 | 0.00 | -0.015 (0.11) | -0.015 (0.14) | -0.055 (0.15) | -0.017 (0.14) | -0.021 (0.14) | -0.054 (0.15) | -0.016 (0.14) | -0.026 (0.15) | -0.044 (0.23) | -0.016 (0.14) | -0.018 (0.14) |
| 87765 | 0.00 | -0.017 (0.09) | -0.017 (0.10) | -0.050 (0.13) | -0.018 (0.10) | -0.022 (0.11) | -0.049 (0.13) | -0.018 (0.10) | -0.028 (0.11) | -0.071 (0.24) | -0.017 (0.10) | -0.017 (0.11) |
| 88044 | 0.00 | 0.008 (0.07) | 0.008 (0.08) | 0.002 (0.11) | 0.009 (0.08) | 0.009 (0.08) | 0.004 (0.11) | 0.009 (0.08) | 0.006 (0.09) | -0.006 (0.13) | 0.008 (0.08) | 0.007 (0.09) |
| 88958† | 0.00 | -0.034 (0.02) | -0.034 (0.03) | -0.036 (0.04) | -0.034 (0.03) | -0.034 (0.03) | -0.035 (0.04) | -0.033 (0.03) | -0.035 (0.03) | -0.038 (0.04) | -0.034 (0.03) | -0.034 (0.03) |
| 89015 | 0.00 | 0.036 (0.07) | 0.037 (0.08) | 0.054 (0.12) | 0.036 (0.08) | 0.038 (0.08) | 0.048 (0.12) | 0.036 (0.08) | 0.041 (0.08) | 0.047 (0.11) | 0.036 (0.08) | 0.036 (0.08) |
| 90803† | 0.00 | 0.032 (0.03) | 0.033 (0.03) | 0.037 (0.05) | 0.033 (0.03) | 0.034 (0.03) | 0.035 (0.05) | 0.033 (0.03) | 0.035 (0.03) | 0.036 (0.04) | 0.033 (0.03) | 0.033 (0.03) |
| 91143† | 0.00 | 0.196 (0.03) | 0.196 (0.04) | 0.196 (0.05) | 0.196 (0.04) | 0.196 (0.04) | 0.192 (0.05) | 0.197 (0.04) | 0.195 (0.04) | 0.193 (0.05) | 0.196 (0.04) | 0.195 (0.04) |
| 91524 | 0.00 | 0.024 (0.03) | 0.025 (0.04) | 0.016 (0.05) | 0.025 (0.04) | 0.024 (0.04) | 0.016 (0.05) | 0.025 (0.04) | 0.022 (0.04) | 0.018 (0.04) | 0.025 (0.04) | 0.025 (0.03) |
| 92017 | 0.00 | 0.022 (0.03) | 0.023 (0.04) | 0.014 (0.05) | 0.023 (0.04) | 0.022 (0.04) | 0.013 (0.05) | 0.023 (0.04) | 0.020 (0.04) | 0.016 (0.04) | 0.023 (0.04) | 0.023 (0.03) |
| 93161 | 0.00 | 0.045 (0.07) | 0.048 (0.09) | 0.015 (0.12) | 0.048 (0.09) | 0.045 (0.09) | 0.018 (0.12) | 0.048 (0.09) | 0.040 (0.10) | 0.013 (0.18) | 0.048 (0.09) | 0.047 (0.09) |
| 93211 | 0.00 | 0.016 (0.03) | 0.017 (0.03) | 0.020 (0.05) | 0.017 (0.03) | 0.017 (0.03) | 0.015 (0.05) | 0.017 (0.03) | 0.018 (0.03) | 0.021 (0.04) | 0.017 (0.03) | 0.018 (0.03) |
| 93324* | 0.12 | 0.067 (0.07) | 0.069 (0.08) | 0.041 (0.10) | 0.068 (0.08) | 0.067 (0.08) | 0.043 (0.10) | 0.069 (0.08) | 0.063 (0.08) | 0.048 (0.13) | 0.069 (0.08) | 0.067 (0.08) |
| 93886 | 0.00 | 0.038 (0.07) | 0.038 (0.08) | 0.016 (0.11) | 0.038 (0.08) | 0.038 (0.09) | 0.018 (0.11) | 0.039 (0.08) | 0.033 (0.09) | 0.010 (0.15) | 0.039 (0.08) | 0.036 (0.09) |
| 93897 | 0.00 | 0.051 (0.03) | 0.052 (0.03) | 0.047 (0.05) | 0.051 (0.03) | 0.051 (0.03) | 0.046 (0.05) | 0.052 (0.03) | 0.051 (0.03) | 0.050 (0.04) | 0.052 (0.03) | 0.052 (0.03) |
| 93901 | 0.00 | 0.015 (0.03) | 0.015 (0.03) | 0.015 (0.05) | 0.015 (0.03) | 0.015 (0.03) | 0.012 (0.05) | 0.015 (0.03) | 0.015 (0.03) | 0.017 (0.04) | 0.015 (0.03) | 0.017 (0.03) |
| 93935† | 0.00 | 0.032 (0.03) | 0.033 (0.03) | 0.037 (0.05) | 0.033 (0.03) | 0.033 (0.03) | 0.035 (0.05) | 0.033 (0.03) | 0.034 (0.03) | 0.035 (0.04) | 0.033 (0.03) | 0.032 (0.03) |
| 94192† | 0.00 | 0.031 (0.03) | 0.032 (0.03) | 0.035 (0.05) | 0.032 (0.03) | 0.032 (0.03) | 0.034 (0.05) | 0.032 (0.03) | 0.033 (0.03) | 0.034 (0.04) | 0.032 (0.03) | 0.031 (0.03) |
| 94212 | 0.00 | 0.047 (0.02) | 0.046 (0.03) | 0.050 (0.04) | 0.046 (0.03) | 0.048 (0.03) | 0.049 (0.04) | 0.047 (0.03) | 0.048 (0.03) | 0.049 (0.04) | 0.047 (0.03) | 0.047 (0.03) |
| 94244 | 0.00 | 0.038 (0.07) | 0.038 (0.08) | 0.016 (0.11) | 0.038 (0.08) | 0.038 (0.09) | 0.018 (0.11) | 0.039 (0.08) | 0.033 (0.09) | 0.010 (0.15) | 0.039 (0.08) | 0.036 (0.09) |
| 94528 | 0.00 | 0.038 (0.07) | 0.038 (0.08) | 0.016 (0.11) | 0.038 (0.08) | 0.038 (0.09) | 0.018 (0.11) | 0.039 (0.08) | 0.033 (0.09) | 0.010 (0.15) | 0.039 (0.08) | 0.036 (0.09) |
| 94990* | 0.25 | 0.206 (0.03) | 0.206 (0.04) | 0.207 (0.05) | 0.206 (0.04) | 0.206 (0.04) | 0.203 (0.05) | 0.207 (0.04) | 0.207 (0.04) | 0.203 (0.05) | 0.206 (0.04) | 0.206 (0.04) |
| 95038† | 0.00 | 0.032 (0.03) | 0.033 (0.03) | 0.037 (0.05) | 0.033 (0.03) | 0.034 (0.03) | 0.036 (0.05) | 0.033 (0.03) | 0.035 (0.03) | 0.035 (0.04) | 0.033 (0.03) | 0.032 (0.03) |
| 95234 | 0.00 | 0.034 (0.04) | 0.035 (0.05) | 0.025 (0.07) | 0.036 (0.05) | 0.033 (0.05) | 0.026 (0.07) | 0.035 (0.05) | 0.032 (0.05) | 0.025 (0.06) | 0.035 (0.05) | 0.034 (0.05) |
| 95236* | -0.15 | -0.106 (0.02) | -0.108 (0.03) | -0.108 (0.04) | -0.109 (0.03) | -0.108 (0.03) | -0.106 (0.04) | -0.108 (0.03) | -0.108 (0.03) | -0.101 (0.04) | -0.108 (0.03) | -0.107 (0.03) |

Table S19: Estimates and asymptotic standard errors for $\beta_1$ in fine-mapping simulation conditional analysis for $n = 2500$. This table removes results from variant 56989830 as it appeared as the top SNP in the majority of replicates, thus, it has too few replicates to be included. Positions are truncated to the last five digits.

| **$G$ pos.** | $\beta_{1G}$ | **Complete** | **RDS** | **Par-spec** | | | **A-opt** | | | **D-opt** | | |
|---|---|---|---|---|---|---|---|---|---|---|---|---|
| | | | | **LM** | **GA** | **TZL** | **LM** | **GA** | **TZL** | **LM** | **GA** | **TZL** |
| 85805 | 0.00 | 0.021 (0.03) | 0.021 (0.03) | 0.018 (0.04) | 0.021 (0.03) | 0.021 (0.03) | 0.017 (0.04) | 0.021 (0.03) | 0.021 (0.03) | 0.018 (0.03) | 0.021 (0.03) | 0.021 (0.03) |
| 86045$^\dagger$ | 0.00 | 0.034 (0.03) | 0.035 (0.03) | 0.037 (0.03) | 0.035 (0.03) | 0.035 (0.03) | 0.036 (0.03) | 0.035 (0.03) | 0.035 (0.03) | 0.036 (0.03) | 0.035 (0.03) | 0.035 (0.03) |
| 86762$^\dagger$ | 0.00 | 0.000 (0.09) | 0.001 (0.09) | 0.004 (0.11) | 0.001 (0.09) | 0.001 (0.09) | 0.001 (0.11) | 0.001 (0.09) | 0.001 (0.09) | -0.009 (0.11) | 0.001 (0.09) | -0.001 (0.09) |
| 86914 | 0.00 | 0.023 (0.03) | 0.024 (0.03) | 0.020 (0.04) | 0.024 (0.03) | 0.024 (0.03) | 0.020 (0.04) | 0.024 (0.03) | 0.024 (0.03) | 0.020 (0.03) | 0.024 (0.03) | 0.024 (0.03) |
| 87015 | 0.00 | -0.015 (0.11) | -0.016 (0.12) | -0.035 (0.15) | -0.016 (0.12) | -0.017 (0.12) | -0.028 (0.15) | -0.015 (0.12) | -0.015 (0.12) | -0.029 (0.15) | -0.016 (0.12) | -0.015 (0.12) |
| 87765 | 0.00 | -0.017 (0.09) | -0.018 (0.09) | -0.039 (0.12) | -0.018 (0.09) | -0.019 (0.09) | -0.033 (0.11) | -0.018 (0.09) | -0.018 (0.09) | -0.038 (0.13) | -0.018 (0.09) | -0.018 (0.09) |
| 88044 | 0.00 | 0.008 (0.07) | 0.009 (0.07) | 0.004 (0.09) | 0.009 (0.07) | 0.009 (0.07) | 0.005 (0.09) | 0.009 (0.07) | 0.010 (0.07) | -0.005 (0.09) | 0.009 (0.07) | 0.009 (0.07) |
| 88958$^\dagger$ | 0.00 | -0.034 (0.02) | -0.034 (0.02) | -0.036 (0.03) | -0.034 (0.02) | -0.034 (0.02) | -0.036 (0.03) | -0.034 (0.02) | -0.034 (0.02) | -0.037 (0.03) | -0.034 (0.02) | -0.034 (0.02) |
| 89015 | 0.00 | 0.036 (0.07) | 0.036 (0.07) | 0.044 (0.08) | 0.036 (0.07) | 0.039 (0.07) | 0.042 (0.08) | 0.036 (0.07) | 0.039 (0.07) | 0.040 (0.08) | 0.036 (0.07) | 0.038 (0.07) |
| 90803$^\dagger$ | 0.00 | 0.032 (0.03) | 0.033 (0.03) | 0.035 (0.03) | 0.033 (0.03) | 0.034 (0.03) | 0.035 (0.03) | 0.033 (0.03) | 0.034 (0.03) | 0.034 (0.03) | 0.033 (0.03) | 0.033 (0.03) |
| 91143$^\dagger$ | 0.00 | 0.196 (0.03) | 0.196 (0.04) | 0.196 (0.04) | 0.196 (0.04) | 0.196 (0.04) | 0.195 (0.04) | 0.196 (0.04) | 0.196 (0.04) | 0.191 (0.04) | 0.196 (0.04) | 0.196 (0.03) |
| 91524 | 0.00 | 0.024 (0.03) | 0.024 (0.03) | 0.020 (0.04) | 0.024 (0.03) | 0.024 (0.03) | 0.020 (0.04) | 0.024 (0.03) | 0.024 (0.03) | 0.021 (0.03) | 0.024 (0.03) | 0.024 (0.03) |
| 92017 | 0.00 | 0.022 (0.03) | 0.022 (0.03) | 0.019 (0.04) | 0.022 (0.03) | 0.022 (0.03) | 0.018 (0.04) | 0.022 (0.03) | 0.022 (0.03) | 0.018 (0.03) | 0.022 (0.03) | 0.022 (0.03) |
| 93161 | 0.00 | 0.045 (0.07) | 0.045 (0.08) | 0.027 (0.10) | 0.045 (0.08) | 0.045 (0.08) | 0.035 (0.10) | 0.045 (0.08) | 0.045 (0.08) | 0.030 (0.11) | 0.045 (0.08) | 0.045 (0.08) |
| 93211 | 0.00 | 0.016 (0.03) | 0.016 (0.03) | 0.019 (0.03) | 0.016 (0.03) | 0.017 (0.03) | 0.017 (0.03) | 0.016 (0.03) | 0.017 (0.03) | 0.019 (0.03) | 0.016 (0.03) | 0.017 (0.03) |
| 93324$^*$ | 0.12 | 0.067 (0.07) | 0.067 (0.07) | 0.054 (0.09) | 0.067 (0.07) | 0.066 (0.07) | 0.060 (0.09) | 0.067 (0.07) | 0.067 (0.07) | 0.056 (0.09) | 0.067 (0.07) | 0.066 (0.07) |
| 93886 | 0.00 | 0.038 (0.07) | 0.037 (0.07) | 0.024 (0.10) | 0.037 (0.07) | 0.037 (0.07) | 0.031 (0.09) | 0.037 (0.07) | 0.038 (0.07) | 0.020 (0.10) | 0.037 (0.07) | 0.037 (0.07) |
| 93897 | 0.00 | 0.051 (0.03) | 0.051 (0.03) | 0.050 (0.03) | 0.051 (0.03) | 0.051 (0.03) | 0.050 (0.03) | 0.051 (0.03) | 0.051 (0.03) | 0.051 (0.03) | 0.051 (0.03) | 0.051 (0.03) |
| 93901 | 0.00 | 0.015 (0.03) | 0.015 (0.03) | 0.016 (0.03) | 0.015 (0.03) | 0.015 (0.03) | 0.014 (0.03) | 0.015 (0.03) | 0.015 (0.03) | 0.017 (0.03) | 0.015 (0.03) | 0.015 (0.03) |
| 93935$^\dagger$ | 0.00 | 0.032 (0.03) | 0.033 (0.03) | 0.035 (0.03) | 0.033 (0.03) | 0.033 (0.03) | 0.035 (0.03) | 0.033 (0.03) | 0.033 (0.03) | 0.034 (0.03) | 0.033 (0.03) | 0.033 (0.03) |
| 94192$^\dagger$ | 0.00 | 0.031 (0.03) | 0.032 (0.03) | 0.034 (0.03) | 0.032 (0.03) | 0.032 (0.03) | 0.034 (0.03) | 0.032 (0.03) | 0.032 (0.03) | 0.033 (0.03) | 0.032 (0.03) | 0.032 (0.03) |
| 94212 | 0.00 | 0.047 (0.02) | 0.047 (0.03) | 0.048 (0.03) | 0.046 (0.03) | 0.047 (0.03) | 0.047 (0.03) | 0.047 (0.03) | 0.047 (0.03) | 0.046 (0.03) | 0.046 (0.03) | 0.047 (0.03) |
| 94244 | 0.00 | 0.038 (0.07) | 0.037 (0.07) | 0.024 (0.10) | 0.037 (0.07) | 0.037 (0.07) | 0.031 (0.09) | 0.037 (0.07) | 0.038 (0.07) | 0.020 (0.10) | 0.037 (0.07) | 0.037 (0.07) |
| 94528 | 0.00 | 0.038 (0.07) | 0.037 (0.07) | 0.024 (0.10) | 0.037 (0.07) | 0.037 (0.07) | 0.031 (0.09) | 0.037 (0.07) | 0.038 (0.07) | 0.020 (0.10) | 0.037 (0.07) | 0.037 (0.07) |
| 94990$^*$ | 0.25 | 0.206 (0.03) | 0.206 (0.03) | 0.207 (0.04) | 0.206 (0.03) | 0.207 (0.03) | 0.206 (0.04) | 0.206 (0.03) | 0.207 (0.03) | 0.201 (0.04) | 0.206 (0.03) | 0.206 (0.03) |
| 95038$^\dagger$ | 0.00 | 0.032 (0.03) | 0.033 (0.03) | 0.035 (0.03) | 0.033 (0.03) | 0.033 (0.03) | 0.035 (0.03) | 0.033 (0.03) | 0.033 (0.03) | 0.034 (0.03) | 0.033 (0.03) | 0.033 (0.03) |
| 95234 | 0.00 | 0.034 (0.04) | 0.035 (0.04) | 0.029 (0.05) | 0.035 (0.04) | 0.034 (0.04) | 0.029 (0.05) | 0.035 (0.04) | 0.034 (0.04) | 0.030 (0.05) | 0.035 (0.04) | 0.034 (0.04) |
| 95236$^*$ | -0.15 | -0.106 (0.02) | -0.107 (0.03) | -0.106 (0.03) | -0.107 (0.03) | -0.107 (0.03) | -0.106 (0.03) | -0.107 (0.03) | -0.107 (0.03) | -0.104 (0.03) | -0.107 (0.03) | -0.107 (0.03) |

Table S20: Empirical power rates at significance level $\alpha = 0.05/29$ and $n = 1250$ for the test $H_0 : \beta_1 = 0$ across 500 replicates in fine-mapping simulation conditional analysis. This table removes results from variant 56989830 as it appeared as the top SNP in the majority of replicates, thus, it has too few replicates to be included. Positions marked with $*$ denote causal variants whereas the ones marked with † denote hitchhikers. The remaining ones are non-causal. Positions are truncated to the last 5 digits.

| ***G* pos.** | $\beta_{1G}$ | **Complete** | **RDS** | **Par-spec** | | | **A-opt** | | | **D-opt** | | |
|---|---|---|---|---|---|---|---|---|---|---|---|---|
| | | | | **LM** | **GA** | **TZL** | **LM** | **GA** | **TZL** | **LM** | **GA** | **TZL** |
| 85805 | 0.00 | 0.6 | 0.4 | 0.2 | 0.2 | 0.6 | 0.0 | 0.4 | 0.2 | 0.0 | 0.4 | 0.2 |
| 86045† | 0.00 | 2.8 | 1.6 | 1.6 | 1.2 | 1.4 | 1.0 | 1.4 | 1.8 | 1.0 | 1.6 | 2.2 |
| 86762† | 0.00 | 0.0 | 0.0 | 0.4 | 0.2 | 0.2 | 0.2 | 0.2 | 0.4 | 0.0 | 0.0 | 0.4 |
| 86914 | 0.00 | 0.6 | 0.6 | 0.2 | 0.2 | 0.8 | 0.2 | 0.4 | 0.2 | 0.8 | 0.4 | 0.4 |
| 87015 | 0.00 | 0.0 | 0.2 | 0.0 | 0.2 | 0.0 | 0.0 | 0.2 | 0.4 | 0.2 | 0.0 | 0.2 |
| 87765 | 0.00 | 0.2 | 0.2 | 0.0 | 0.2 | 0.0 | 0.0 | 0.4 | 0.4 | 0.4 | 0.2 | 0.0 |
| 88044 | 0.00 | 0.0 | 0.0 | 0.4 | 0.2 | 0.2 | 0.0 | 0.0 | 0.0 | 0.0 | 0.0 | 0.0 |
| 88958† | 0.00 | 6.6 | 3.0 | 0.4 | 2.4 | 2.0 | 1.6 | 2.4 | 2.4 | 0.4 | 3.2 | 3.0 |
| 89015 | 0.00 | 0.4 | 0.4 | 0.2 | 0.4 | 0.4 | 0.4 | 0.4 | 0.4 | 0.2 | 0.4 | 0.2 |
| 90803† | 0.00 | 2.0 | 1.8 | 1.6 | 1.2 | 2.2 | 1.0 | 1.6 | 1.8 | 0.8 | 1.8 | 1.8 |
| 91143† | 0.00 | 99.6 | 96.2 | 67.0 | 96.4 | 95.6 | 62.2 | 96.2 | 95.8 | 73.8 | 96.6 | 97.0 |
| 91524 | 0.00 | 0.6 | 0.8 | 0.4 | 0.8 | 0.6 | 0.4 | 0.6 | 0.8 | 0.0 | 0.8 | 0.6 |
| 92017 | 0.00 | 0.2 | 0.8 | 0.8 | 0.8 | 0.8 | 0.4 | 0.8 | 0.4 | 0.2 | 0.8 | 0.8 |
| 93161 | 0.00 | 0.6 | 0.6 | 0.0 | 0.6 | 0.4 | 0.0 | 0.8 | 0.6 | 0.0 | 0.6 | 0.8 |
| 93211 | 0.00 | 1.4 | 1.4 | 0.0 | 1.2 | 1.2 | 0.0 | 1.2 | 0.8 | 0.0 | 1.2 | 1.2 |
| 93324* | 0.12 | 2.4 | 1.4 | 0.4 | 1.4 | 1.0 | 1.0 | 1.6 | 0.8 | 0.2 | 1.4 | 1.2 |
| 93886 | 0.00 | 0.2 | 0.4 | 0.0 | 0.4 | 0.2 | 0.2 | 0.2 | 0.0 | 0.0 | 0.4 | 0.2 |
| 93897 | 0.00 | 8.8 | 4.0 | 2.2 | 3.6 | 4.0 | 1.6 | 4.4 | 4.2 | 1.8 | 3.8 | 3.2 |
| 93901 | 0.00 | 1.2 | 1.2 | 0.4 | 1.0 | 1.0 | 0.2 | 1.0 | 1.0 | 0.4 | 1.2 | 1.4 |
| 93935† | 0.00 | 2.2 | 2.2 | 1.4 | 1.8 | 2.4 | 1.0 | 1.8 | 2.2 | 0.8 | 2.2 | 2.2 |
| 94192† | 0.00 | 1.8 | 1.8 | 1.4 | 1.8 | 2.2 | 0.8 | 1.6 | 1.8 | 0.6 | 1.8 | 2.0 |
| 94212 | 0.00 | 13.4 | 7.4 | 1.2 | 6.8 | 7.8 | 2.2 | 6.8 | 6.6 | 2.6 | 7.6 | 8.2 |
| 94244 | 0.00 | 0.2 | 0.4 | 0.0 | 0.4 | 0.2 | 0.2 | 0.2 | 0.0 | 0.0 | 0.4 | 0.2 |
| 94528 | 0.00 | 0.2 | 0.4 | 0.0 | 0.4 | 0.2 | 0.2 | 0.2 | 0.0 | 0.0 | 0.4 | 0.2 |
| 94990* | 0.25 | 100.0 | 98.8 | 78.2 | 98.8 | 99.2 | 75.2 | 99.2 | 99.0 | 83.4 | 99.2 | 99.2 |
| 95038† | 0.00 | 1.2 | 1.8 | 1.4 | 1.4 | 1.8 | 1.2 | 2.0 | 2.0 | 0.8 | 2.4 | 1.8 |
| 95234 | 0.00 | 1.2 | 1.2 | 0.8 | 1.2 | 0.6 | 0.4 | 1.2 | 0.8 | 0.0 | 1.2 | 0.8 |
| 95236* | -0.15 | 89.4 | 73.2 | 31.4 | 72.2 | 71.0 | 29.4 | 72.2 | 70.6 | 31.2 | 73.6 | 72.0 |

Table S21: Empirical power rates at significance level $\alpha = 0.05/29$ for causal variants only and $n = 2500$ for the test $H_0 : \beta_1 = 0$ across 500 replicates in fine-mapping simulation conditional analysis. This table removes results from variant 56989830 as it appeared as the top SNP in the majority of replicates, thus, it has too few replicates to be included. Positions are truncated to the last 5 digits.

| ***G* pos.** | $\beta_{1G}$ | **Complete** | **RDS** | **Par-spec** | | | **A-opt** | | | **D-opt** | | |
|---|---|---|---|---|---|---|---|---|---|---|---|---|
| | | | | **LM** | **GA** | **TZL** | **LM** | **GA** | **TZL** | **LM** | **GA** | **TZL** |
| 85805 | 0.00 | 0.6 | 0.8 | 0.2 | 0.8 | 0.6 | 0.2 | 0.8 | 0.8 | 0.4 | 0.8 | 0.6 |
| 86045$^{\dagger}$ | 0.00 | 2.8 | 3.2 | 2.6 | 3.2 | 3.2 | 1.4 | 3.4 | 3.0 | 1.8 | 3.6 | 2.8 |
| 86762$^{\dagger}$ | 0.00 | 0.0 | 0.0 | 0.6 | 0.0 | 0.0 | 0.4 | 0.0 | 0.0 | 0.2 | 0.0 | 0.0 |
| 86914 | 0.00 | 0.6 | 0.4 | 0.4 | 0.4 | 0.4 | 0.2 | 0.4 | 0.2 | 0.2 | 0.6 | 0.6 |
| 87015 | 0.00 | 0.0 | 0.0 | 0.4 | 0.0 | 0.0 | 0.4 | 0.0 | 0.0 | 0.2 | 0.0 | 0.2 |
| 87765 | 0.00 | 0.2 | 0.2 | 0.0 | 0.2 | 0.2 | 0.2 | 0.2 | 0.2 | 0.0 | 0.2 | 0.4 |
| 88044 | 0.00 | 0.0 | 0.0 | 0.0 | 0.0 | 0.0 | 0.0 | 0.2 | 0.2 | 0.0 | 0.0 | 0.0 |
| 88958$^{\dagger}$ | 0.00 | 6.6 | 5.4 | 2.6 | 5.2 | 4.8 | 2.0 | 5.2 | 4.6 | 4.2 | 5.2 | 4.4 |
| 89015 | 0.00 | 0.4 | 0.4 | 0.0 | 0.4 | 0.6 | 0.0 | 0.4 | 0.6 | 0.2 | 0.4 | 0.6 |
| 90803$^{\dagger}$ | 0.00 | 2.0 | 2.4 | 2.0 | 2.4 | 2.8 | 1.8 | 2.4 | 2.8 | 1.2 | 2.4 | 2.2 |
| 91143$^{\dagger}$ | 0.00 | 99.6 | 99.6 | 95.8 | 99.6 | 99.4 | 95.6 | 99.6 | 99.6 | 96.0 | 99.6 | 99.6 |
| 91524 | 0.00 | 0.6 | 0.6 | 0.0 | 0.6 | 0.8 | 0.0 | 0.6 | 0.8 | 0.4 | 0.6 | 0.6 |
| 92017 | 0.00 | 0.2 | 0.6 | 0.8 | 0.8 | 0.6 | 0.4 | 0.6 | 0.8 | 0.4 | 0.6 | 0.6 |
| 93161 | 0.00 | 0.6 | 0.4 | 0.0 | 0.4 | 0.4 | 0.8 | 0.4 | 0.2 | 0.2 | 0.4 | 0.8 |
| 93211 | 0.00 | 1.4 | 0.8 | 0.4 | 0.8 | 1.2 | 0.2 | 1.0 | 1.2 | 0.8 | 0.8 | 0.8 |
| 93324* | 0.12 | 2.4 | 2.2 | 1.2 | 2.2 | 1.6 | 1.4 | 2.0 | 2.0 | 0.4 | 2.4 | 1.8 |
| 93886 | 0.00 | 0.2 | 0.2 | 0.0 | 0.2 | 0.2 | 0.2 | 0.2 | 0.4 | 0.0 | 0.2 | 0.0 |
| 93897 | 0.00 | 8.8 | 7.0 | 4.2 | 7.4 | 6.4 | 3.0 | 8.0 | 6.8 | 4.8 | 6.8 | 7.4 |
| 93901 | 0.00 | 1.2 | 1.2 | 1.0 | 1.2 | 1.2 | 0.6 | 1.2 | 1.2 | 1.6 | 1.2 | 1.4 |
| 93935$^{\dagger}$ | 0.00 | 2.2 | 2.4 | 2.0 | 2.8 | 2.8 | 1.8 | 2.6 | 2.4 | 1.0 | 2.4 | 3.0 |
| 94192$^{\dagger}$ | 0.00 | 1.8 | 2.4 | 1.6 | 2.4 | 2.0 | 1.8 | 2.4 | 2.2 | 1.6 | 2.4 | 2.2 |
| 94212 | 0.00 | 13.4 | 11.6 | 8.4 | 11.0 | 11.8 | 7.6 | 11.6 | 10.2 | 8.0 | 11.8 | 11.4 |
| 94244 | 0.00 | 0.2 | 0.2 | 0.0 | 0.2 | 0.2 | 0.2 | 0.2 | 0.4 | 0.0 | 0.2 | 0.0 |
| 94528 | 0.00 | 0.2 | 0.2 | 0.0 | 0.2 | 0.2 | 0.2 | 0.2 | 0.4 | 0.0 | 0.2 | 0.0 |
| 94990* | 0.25 | 100.0 | 100.0 | 99.0 | 100.0 | 100.0 | 99.0 | 100.0 | 100.0 | 99.2 | 100.0 | 100.0 |
| 95038$^{\dagger}$ | 0.00 | 1.2 | 2.0 | 2.0 | 2.2 | 2.2 | 2.2 | 2.0 | 2.0 | 0.8 | 2.0 | 1.8 |
| 95234 | 0.00 | 1.2 | 1.4 | 0.6 | 1.2 | 1.6 | 0.8 | 1.4 | 1.4 | 0.8 | 1.4 | 1.2 |
| 95236* | -0.15 | 89.4 | 86.6 | 68.8 | 86.6 | 85.4 | 70.8 | 86.4 | 84.4 | 68.0 | 86.6 | 86.8 |

Table S22: Estimates and asymptotic standard errors for $\beta_2$ in fine-mapping simulation conditional analysis for $n = 1250, 2500$ for the most common top SNP, 89830.

| $n$ | $G_{top}$ pos. | $\beta_2$ | Complete | RDS | Par-spec | | | A-opt | | | D-opt | | |
|---|---|---|---|---|---|---|---|---|---|---|---|---|---|
| | | | | | LM | GA | TZL | LM | GA | TZL | LM | GA | TZL |
| 1250 | 89830* | -0.20 | -0.162 (0.03) | -0.162 (0.03) | -0.158 (0.04) | -0.162 (0.03) | -0.162 (0.03) | -0.159 (0.04) | -0.162 (0.03) | -0.161 (0.03) | -0.158 (0.04) | -0.162 (0.03) | -0.162 (0.03) |
| 2500 | | | | -0.161 (0.03) | -0.160 (0.03) | -0.161 (0.03) | -0.161 (0.03) | -0.159 (0.03) | -0.161 (0.03) | -0.161 (0.03) | -0.159 (0.03) | -0.161 (0.03) | -0.161 (0.03) |

Table S23: Empirical power rates at significance level $\alpha = 0.05/29$ for the test $H_0 : \beta_2 = 0$ across 500 replicates in fine-mapping simulation conditional analysis for $n = 1250, 2500$ for the most common top SNP, 89830.

| $n$ | $G_{top}$ pos. | $\beta_2$ | Complete | RDS | Par-spec | | | A-opt | | | D-opt | | |
|---|---|---|---|---|---|---|---|---|---|---|---|---|---|
| | | | | | LM | GA | TZL | LM | GA | TZL | LM | GA | TZL |
| 1250 | 89830* | -0.20 | 96.7 | 96.3 | 79.3 | 96.2 | 96.2 | 80.0 | 96.3 | 96.0 | 83.3 | 96.3 | 96.3 |
| 2500 | | | | 96.7 | 95.6 | 96.7 | 96.6 | 95.5 | 96.7 | 96.7 | 95.7 | 96.7 | 96.7 |

##### S6.1.2 Figures

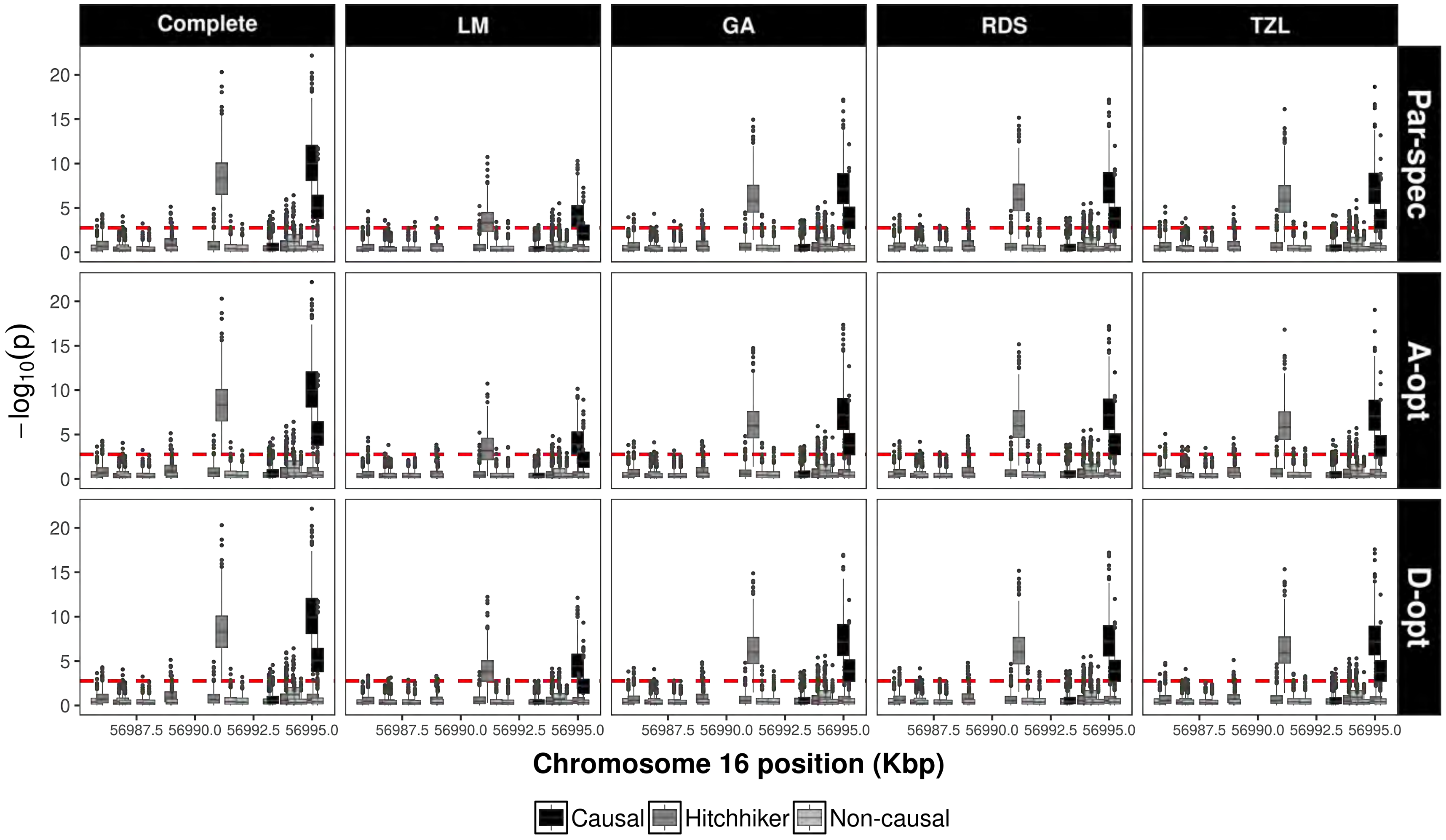

Figure S26: Boxplots of the $(-\log_{10})$ $p$-values across 500 replicates in the fine-mapping simulation conditional analyses for a phase 2 sample size of $n = 1250$ across optimality criteria (for LM, GA, and TZL only): parameter-specific, A- and D- optimality in each row facet. Each column facet corresponds to the complete data analysis and studied designs respectively. The dashed line corresponds to a Bonferroni-corrected significance threshold of $\alpha = 0.05/29$.

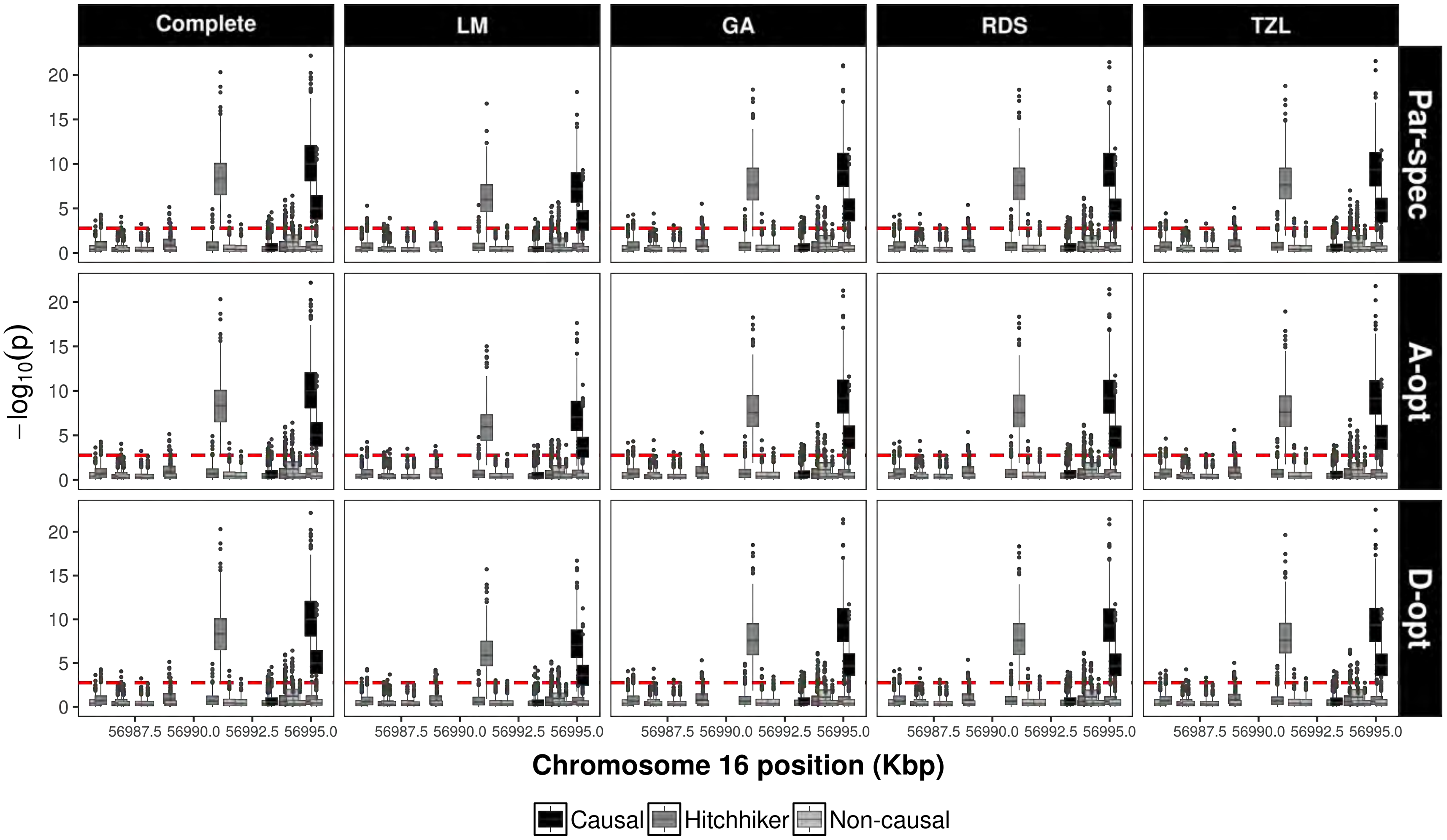

Figure S27: Boxplots of the $(-\log_{10})$ $p$-values across 500 replicates in the fine-mapping simulation conditional analyses for a phase 2 sample size of $n = 2500$ across optimality criteria (for LM, GA, and TZL only): parameter-specific, A- and D- optimality in each row facet. Each column facet corresponds to the complete data analysis and studied designs respectively. The dashed line corresponds to a Bonferroni-corrected significance threshold of $\alpha = 0.05/29$.

### S7 Rare-variant analysis

Beyond common variants, the appeal of fine-mapping targeted regions also lies in identifying rare variants (RVs) that may play a causal role. The main challenge of rare-variant analysis, however, is the very large sample sizes required to pinpoint them. A number of statistical approaches have been proposed to study RVs.[4] Here, we describe how to use the ML framework described in the main text to construct a genetic score to implement burden tests for association analysis of RVs. We first define the genetic score, $GS$, for an $i$th subject across $m$ RVs as follows

$$GS_i = \sum_{j=1}^{m} w_j G_{ij},$$

where $w_j$ is a weight for variant $j$ and $G_{ij}$ is the $j$th RV for the $i$th subject denoting the allele count (additive model), a risk allele indicator (dominant model), or some alternative genetic model specification.

We then carry out ML analyses described in Section 2 by replacing $G$ in equation 1 with $GS$. The main challenge in this setting is that $GS$ may no longer be discrete. However, the described ML approach can still be applied by rounding the $GS$ values to the nearest decimal during optimization via the EM algorithm, which reduces the complexity in the data augmentation procedure. We analyze each fine-mapped gene under two different assumptions to upweigth rare variants: 1) $w_j = [p_j(1 - p_j)]^{-1/2}$ as proposed by Madsen and Browning[5] and 2) $w_j = \text{beta}(p_j, 1, 25)$ as descibed by Wu *et al.*[6]. In addition, we determine RVs by specifying two different thresholds with respect to the maximum observed MAF in the complete data: 0.1% and 1%, respectively.

#### S7.1 Results

We found no evidence against the null hypothesis, $\text{H}_0\text{:}\beta_{GS} = 0$, for neither of the studied genes regardless of the RV threshold or weighting scheme, which could be partially attributed to limited power. Overall, the estimates from the GA, RDS and TZL designs do not follow those of the complete data analysis as closely as in the common variant analyses; although estimation improves with $n$, as expected (Table S24). The lack of agreement between complete data analysis and ML of the studied designs can be due to low observed correlation between the genetic score and the GWAS SNPs (rs1260326 and rs10096633), making $Z$ an imperfect auxiliary variable.

##### S7.1.1 Tables

Table S24: Estimation and testing results for (log-transformed) triglyceride levels across rare-variants (RVs) via burden tests in the NFBC66. The weights $w_j$ used to construct the genetic score ($GS$) correspond to $[p_j(1-p_j)]^{-1/2}$ (**A**) and beta$(p_j, 1, 25)$ (**B**). The last two columns denote the Pearson correlation between the genetic score ($GS$) and each of the GWAS SNPs, rs1260326 ($Z_1$) and rs10096633($Z_2$).

| $w_j$ | $n$ | max. MAF | # of RVs | Gene | $\hat{\beta}_{GS}$ (s.e.[$\hat{\beta}_{GS}$]) $p-$value | | | | $r_{GS,Z_1}$ | $r_{GS,Z_2}$ |
|---|---|---|---|---|---|---|---|---|---|---|
| | | | | | **Complete** | **GA** | **RDS** | **TZL** | | |
| **A** | 1123 | 0.001 | 15 | APOA5 | 0.0013 (0.001) p=0.309 | 0.0000 (0.001) p=0.963 | 0.0000 (0.001) p=1.000 | 0.0001 (0.001) p=0.894 | -0.014 | -0.018 |
| | | | 43 | GCKR | -0.0010 (0.001) p=0.174 | -0.0006 (0.001) p=0.494 | -0.0007 (0.001) p=0.417 | -0.0007 (0.001) p=0.425 | 0.013 | -0.011 |
| | | | 57 | LPL | -0.0007 (0.000) p=0.134 | -0.0014 (0.001) p=0.079 | -0.0015 (0.001) p=0.078 | -0.0012 (0.001) p=0.154 | 0.014 | -0.009 |
| | | 0.010 | 28 | APOA5 | 0.0001 (0.001) p=0.951 | -0.0004 (0.001) p=0.613 | -0.0004 (0.001) p=0.651 | -0.0004 (0.001) p=0.650 | -0.029 | -0.009 |
| | | | 75 | GCKR | -0.0004 (0.001) p=0.515 | 0.0000 (0.001) p=0.958 | 0.0000 (0.001) p=0.989 | 0.0001 (0.001) p=0.927 | 0.093 | -0.024 |
| | | | 120 | LPL | -0.0002 (0.000) p=0.587 | -0.0004 (0.000) p=0.383 | -0.0005 (0.000) p=0.342 | -0.0003 (0.000) p=0.520 | -0.012 | 0.061 |
| | 2246 | 0.001 | 15 | APOA5 | 0.0013 (0.001) p=0.309 | 0.0010 (0.001) p=0.415 | 0.0011 (0.001) p=0.390 | 0.0010 (0.001) p=0.427 | -0.014 | -0.018 |
| | | | 43 | GCKR | -0.0010 (0.001) p=0.174 | -0.0008 (0.001) p=0.339 | -0.0008 (0.001) p=0.334 | -0.0008 (0.001) p=0.324 | 0.013 | -0.011 |
| | | | 57 | LPL | -0.0007 (0.000) p=0.134 | -0.0010 (0.001) p=0.080 | -0.0010 (0.001) p=0.076 | -0.0010 (0.001) p=0.098 | 0.014 | -0.009 |
| | | 0.010 | 28 | APOA5 | 0.0001 (0.001) p=0.951 | -0.0002 (0.001) p=0.855 | -0.0001 (0.001) p=0.878 | -0.0001 (0.001) p=0.889 | -0.029 | -0.009 |
| | | | 75 | GCKR | -0.0004 (0.001) p=0.515 | -0.0002 (0.001) p=0.788 | -0.0002 (0.001) p=0.757 | -0.0002 (0.001) p=0.741 | 0.093 | -0.024 |
| | | | 120 | LPL | -0.0002 (0.000) p=0.587 | -0.0003 (0.000) p=0.417 | -0.0003 (0.000) p=0.411 | -0.0003 (0.000) p=0.475 | -0.012 | 0.061 |
| **B** | 1123 | 0.001 | 15 | APOA5 | 0.0030 (0.002) p=0.182 | 0.0006 (0.002) p=0.781 | 0.0005 (0.002) p=0.812 | 0.0007 (0.002) p=0.713 | -0.013 | -0.017 |
| | | | 43 | GCKR | -0.0019 (0.001) p=0.125 | -0.0012 (0.002) p=0.445 | -0.0014 (0.001) p=0.368 | -0.0013 (0.001) p=0.376 | 0.016 | -0.009 |
| | | | 57 | LPL | -0.0012 (0.001) p=0.177 | -0.0023 (0.001) p=0.114 | -0.0023 (0.001) p=0.114 | -0.0018 (0.001) p=0.217 | 0.014 | -0.007 |
| | | 0.010 | 28 | APOA5 | -0.0001 (0.001) p=0.919 | -0.0002 (0.001) p=0.766 | -0.0001 (0.001) p=0.864 | -0.0002 (0.001) p=0.781 | -0.027 | -0.003 |
| | | | 75 | GCKR | -0.0002 (0.001) p=0.682 | 0.0000 (0.001) p=0.954 | 0.0000 (0.001) p=0.954 | 0.0001 (0.001) p=0.867 | 0.130 | -0.027 |
| | | | 120 | LPL | 0.0001 (0.000) p=0.811 | -0.0001 (0.000) p=0.739 | -0.0002 (0.000) p=0.664 | -0.0001 (0.000) p=0.839 | -0.026 | 0.105 |
| | 2246 | 0.001 | 15 | APOA5 | 0.0030 (0.002) p=0.182 | 0.0024 (0.002) p=0.286 | 0.0025 (0.002) p=0.269 | 0.0024 (0.002) p=0.296 | -0.013 | -0.017 |
| | | | 43 | GCKR | -0.0019 (0.001) p=0.125 | -0.0015 (0.001) p=0.283 | -0.0015 (0.001) p=0.278 | -0.0015 (0.001) p=0.273 | 0.016 | -0.009 |
| | | | 57 | LPL | -0.0012 (0.001) p=0.177 | -0.0016 (0.001) p=0.120 | -0.0016 (0.001) p=0.114 | -0.0015 (0.001) p=0.144 | 0.014 | -0.007 |
| | | 0.010 | 28 | APOA5 | -0.0001 (0.001) p=0.919 | -0.0002 (0.001) p=0.726 | -0.0002 (0.001) p=0.732 | -0.0002 (0.001) p=0.767 | -0.027 | -0.003 |
| | | | 75 | GCKR | -0.0002 (0.001) p=0.682 | -0.0001 (0.001) p=0.891 | -0.0001 (0.001) p=0.851 | -0.0001 (0.001) p=0.871 | 0.130 | -0.027 |
| | | | 120 | LPL | 0.0001 (0.000) p=0.811 | 0.0000 (0.000) p=0.909 | 0.0000 (0.000) p=0.904 | 0.0000 (0.000) p=0.944 | -0.026 | 0.105 |
